# Assessing equity in effects of nutritional supplementation for child growth, development, and anemia

**DOI:** 10.1101/2025.06.05.25329064

**Authors:** Pearl Anne Ante-Testard, Charles D. Arnold, K. Ryan Wessells, Seth Adu-Afarwuah, Per Ashorn, Elodie Becquey, Kenneth H. Brown, Parul Christian, John M. Colford, Lia C.H. Fernald, Emanuela Galasso, Sonja Y. Hess, Jean H. Humphrey, Lieven Huybregts, Lora L. Iannotti, Stephen P. Luby, Kenneth Maleta, Clair Null, Andrew J. Prendergast, Ann M. Weber, Hasmot Ali, Shahjahan Ali, Ulla Ashorn, Jaden Bendabenda, Bernard Chasekwa, Loty Diop, Sherlie Jean-Louis Dulience, Kaniz Jannat, Chiza Kumwenda, Anna Lartey, Agnes Le Port, Jef L Leroy, Charles Mangani, Susana Matias, Malay Kanti Mridha, Robert Ntozini, Harriet Okronipa, Jean-Bosco Ouedraogo, John Phuka, Mahbubur Rahman, Lisy Ratsifandrihamanana, Saijuddin Shaikh, Abu Ahmed Shamim, Naume Tavengwa, Mariama Touré, Patricia Wolff, Tarik Benmarhnia, Christine P. Stewart, Kathryn G. Dewey, Benjamin F. Arnold

## Abstract

Undernutrition in early childhood causes stunted growth, cognitive delays, and anemia, with effects often magnified among children from the poorest households. Small-quantity lipid-based nutrient supplements (SQ-LNS) are effective in addressing undernutrition and improving child development. As momentum builds to scale up SQ-LNS for children aged 6-24 months in the Global South, a key concern is achieving equity in its distribution and outcomes. We performed equity analysis of individual participant data from 14 randomized controlled trials in nine countries (N=37,707 children) to assess SQ-LNS effects on child growth, development, and anemia across levels of an international wealth index. Benefits of SQ-LNS were consistent across the wealth spectrum, leading to similar improvements in child growth, development, and anemia regardless of wealth. However, such equal benefits of SQ-LNS did not erase large inequities in child growth and development between the poorest and wealthier households.

---

Undernutrition remains a global health crisis affecting children worldwide^1^. According to the United Nations International Children’s Fund (UNICEF), 440 million children, two-thirds of the world’s children under the age of five, do not consume diverse diets that provide sufficient nutrients necessary for healthy growth and development^2^. Nutrition in the first two years of life is especially critical, yet only one in three children aged 6-23 months receives the minimum required dietary diversity^3^. In 2022, the World Health Organization (WHO) reported that approximately 148 million children suffered from stunting, while 45 million experienced wasting at any given time; both conditions can be caused by undernutrition^4,5^. Undernutrition hinders children from reaching their full potential and, in extreme cases, can be life-threatening. Nearly half of all deaths among children under five are linked to undernutrition^6^.

Inadequate intake of essential nutrients during pregnancy^7–9^ and early childhood can have severe consequences, posing risks to child survival, physical growth^10^, and neurobehavioral development^2^. The impact of undernutrition on these acute and long-term outcomes may be worsened by poverty, as individuals in the poorest households are consistently at higher risk of infectious diseases, antimicrobial resistance, and delayed vaccination^11^. Wealthier households tend to have more resources and time for activities that improve health such as access to healthy food^12,13^ and access to activities that improve health^12^, while poorer households often face poor diets^13^.

Sustainable Development Goal 2.2 aims to address undernutrition^14^. One potential preventive intervention is the provision of small-quantity lipid-based nutrient supplements (SQ-LNS) to complement the diets of children aged 6-23 months^15^. SQ-LNS is a paste that provides about 120 kcal (about four teaspoons) per day, and is designed to fortify children’s diets to prevent undernutrition by supplementing, not replacing, human milk and locally available nutrient-rich foods^15^. Its base typically includes vegetable oil rich in omega-3 fatty acids, legumes (e.g., peanuts, chickpeas, lentils, or soybeans), milk powder, and a small amount of sugar. The formulation is fortified with 22 vitamins and minerals at levels close to the daily recommended intake of these nutrients for children 6-24 months of age. SQ-LNS provision leads to improved linear growth, ponderal growth, cognitive development, and reduced anemia prevalence among children 6-24 months old without displacing breast milk ^16–19^. The strength of evidence based on a meta-analysis of randomized controlled trials (RCTs) has led to a recommendation by the World Bank to scale up SQ-LNS for children aged 6-24 months^20^ in the Global South. Since poorer and wealthier households may differ in how they access and engage with interventions^21^, it is critical to assess these disparities as global initiatives consider scaling up SQ-LNS to ensure that children in need are effectively reached. Delivering blanket supplementation to entire populations is often expensive and logistically challenging, making it essential to explore equitable distribution strategies that result in greatest benefit — recognizing that the potential to benefit may not always align with the level of need^22^.

Here, we conducted an individual participant data analysis of 14 RCTs that studied the effects of SQ-LNS on child growth and development outcomes^23–36^. Previous analyses of the impact of SQ-LNS investigated several dimensions of effect modification, including socioeconomic status, but for that factor only considered above- and below-within-study median household wealth based on individual study asset indices^16–18^. Our objectives were to compute more granular, internationally standardized measures of wealth across the trials to: (i) assess inequalities in child achieved growth and development and anemia by wealth standard, and (ii) estimate the effect of SQ-LNS on those outcomes across varying levels of wealth standards.

Throughout this study, we use the term *inequality* to refer to systematic differences in outcomes between individuals or groups, without implying a value judgment regarding fairness^37^. In contrast, *inequity* refers to differences that are considered avoidable and unjust. As defined by WHO, *equity* is the absence of avoidable disparities among socially, geographically, economically, or demographically defined groups^38^. In our analysis, *inequality* is used when reporting objectively measured differences in benefits and outcomes, whereas *equity* and *inequity* were used to describe the implications of these differences.

## Results

### Characteristics of the RCTs included in the study

We built on a previous meta-analyses of 14 prospective RCTs of SQ-LNS administered to infants and young children aged 6-24 months^23–36^ (**Table 1**). The trials were conducted in sub-Saharan Africa (Burkina Faso, Ghana, Kenya, Madagascar, Malawi, Mali, and Zimbabwe; 10 trials), Bangladesh (three trials), and Haiti (one trial). Of the 14 studies included, eight were efficacy trials, and six were effectiveness trials. Most studies used a cluster-randomized design except for Ghana, Haiti, DYAD-Ghana (DYAD-G), DYAD-Malawi (DYAD-M), and DOSE, which were individually randomized. Overall, this analysis included 37,707 infants and young children aged 6-24 months.

**Table 1.** Randomized controlled trials that were included in the individual participant data meta-analysis.

| Study Name | Study | Country | Type of study | Cluster-randomized | Child SQ-LNS Age at start in months (duration) | Maternal SQ-LNS | Children<sup>a</sup> |
|---|---|---|---|---|---|---|---|
| **JiVitA-4** | Christian 2015 | Bangladesh | Efficacy | Y | 6 (12) | N | 4568 |
| **RDNS** | Dewey 2017 | Bangladesh | Effectiveness | Y | 6 (18) | Y<sup>c</sup> | 2567 |
| **WASH-B Bangladesh** | Luby 2018 | Bangladesh | Efficacy | Y<br>(block) | 6 (18) | N | 4792 |
| **PROMIS-BF** | Becquey 2019 | Burkina Faso | Effectiveness | Y | 6 (12) | N | 1782 |
| **PROMIS-BF CS** |  |  | Effectiveness<br>Repeated cross-sectional |  |  |  | 1157 |
| **iLiNS-Zinc** | Hess 2015 | Burkina Faso | Efficacy | Y | 9 (9) | N | 2647 |
| **Ghana** | Adu Afarwuah 2007 | Ghana | Effectiveness | N<br>(individually randomized) | 6 (6) | N | 194 |
| **iLiNS-DYAD-G** | Adu Afarwuah 2016 | Ghana | Efficacy | N<br>(individually randomized) | 6 (12) | Y | 1113 |
| **Haiti** | Iannotti 2014 | Haiti | Efficacy | N<br>(individually randomized) | 6-11 (6-12) | N | 322 |
| **WASH-B Kenya** | Null 2018 | Kenya | Efficacy | Y | 6 (18) | N | 6815 |
| **MAHAY** | Galasso 2019 | Madagascar | Effectiveness | Y<br>(community) | 6-11 (6-12) | Y<sup>c</sup> | 3438 |
| **iLiNS-DYAD-M** | Ashorn 2015 | Malawi | Efficacy | N<br>(individually randomized) | 6 (12) | Y | 675 |
| **iLiNS-DOSE** | Maleta 2015 | Malawi | Efficacy | N<sup>b</sup> | 6 (12) | N | 1018 |
| **PROMIS-M** | Huybregts 2019 | Mali | Effectiveness | Y | 6 (18) | N | 1013 |
| **PROMIS-M CS** |  |  | Effectiveness<br>Repeated cross-sectional |  |  |  | 1927 |
| **SHINE HIV-** | Humphrey 2019 | Zimbabwe | Effectiveness | Y | 6 (12) | N | 3679 |

<sup>a</sup> Including all variables of interest and excluding missing values for the intervention variable. SQ-LNS: Small quantity lipid-based nutrient supplement; N: no; <sup>b</sup> indicates that the study was individually randomized using one set of randomization blocks; <sup>c</sup> indicates that the trial included at least one arm with maternal SQ-LNS combined with child SQ-LNS and at least one arm with child SQ-LNS alone.

The primary analyses compared outcomes between children who received SQ-LNS (defined as LNS providing ∼120 kcal per day, hereafter SQ-LNS) versus no intervention or an intervention that did not include any form of LNS or child supplementation (hereafter, control). Consistent with prior analyses^16^, study arms that included provision of maternal SQ-LNS, in addition to child SQ-LNS (**Table 1**), were excluded from the primary analysis but were included in sensitivity analyses, with results that were consistent with the primary analysis (**Text S1**), except for socioemotional scores.

### Harmonized measures of wealth across trials

We used the International Wealth Index (IWI) as our main indicator of wealth. The IWI is a harmonized composite measure of the level of material wellbeing or standard of living of households using 12 asset-based variables with 20 indicators in total^39^ reflecting ownership of durables, housing quality, and access to public services (**Text S2**). Details on how the IWI was constructed are in the **Methods**. The distribution of asset variables and indicators was balanced between the control and SQ-LNS groups (**Tables S2A and S2B,** respectively). The IWI was harmonized and successfully constructed across all studies, allowing for consistent comparison of socioeconomic status despite differences in study settings. In most studies, the ownership of asset variables (e.g., TV, phone and car) increased with higher IWI levels (**Tables S3A and S3B**). The wealthier households were more likely to own high-quality assets (e.g., high-quality water source), whereas the poorest households predominantly owned lower-quality assets (e.g., low-quality water source). The aggregate distribution of the IWI across studies was unimodal, with overlap across studies for most of the distributions (**Fig. 1**). While models were fit using the full range of the IWI (0–100), predictions were restricted to IWI values between 0 and 70, as relatively few children had IWI scores ≥70. We focused on this range of common support — the area where the IWI distributions across studies overlap for subsequent effect measure modification analyses. All studies were conducted in deprived settings. Thus, *wealthier* children signify relatively less poor than the poorest.

**Fig. 1.**
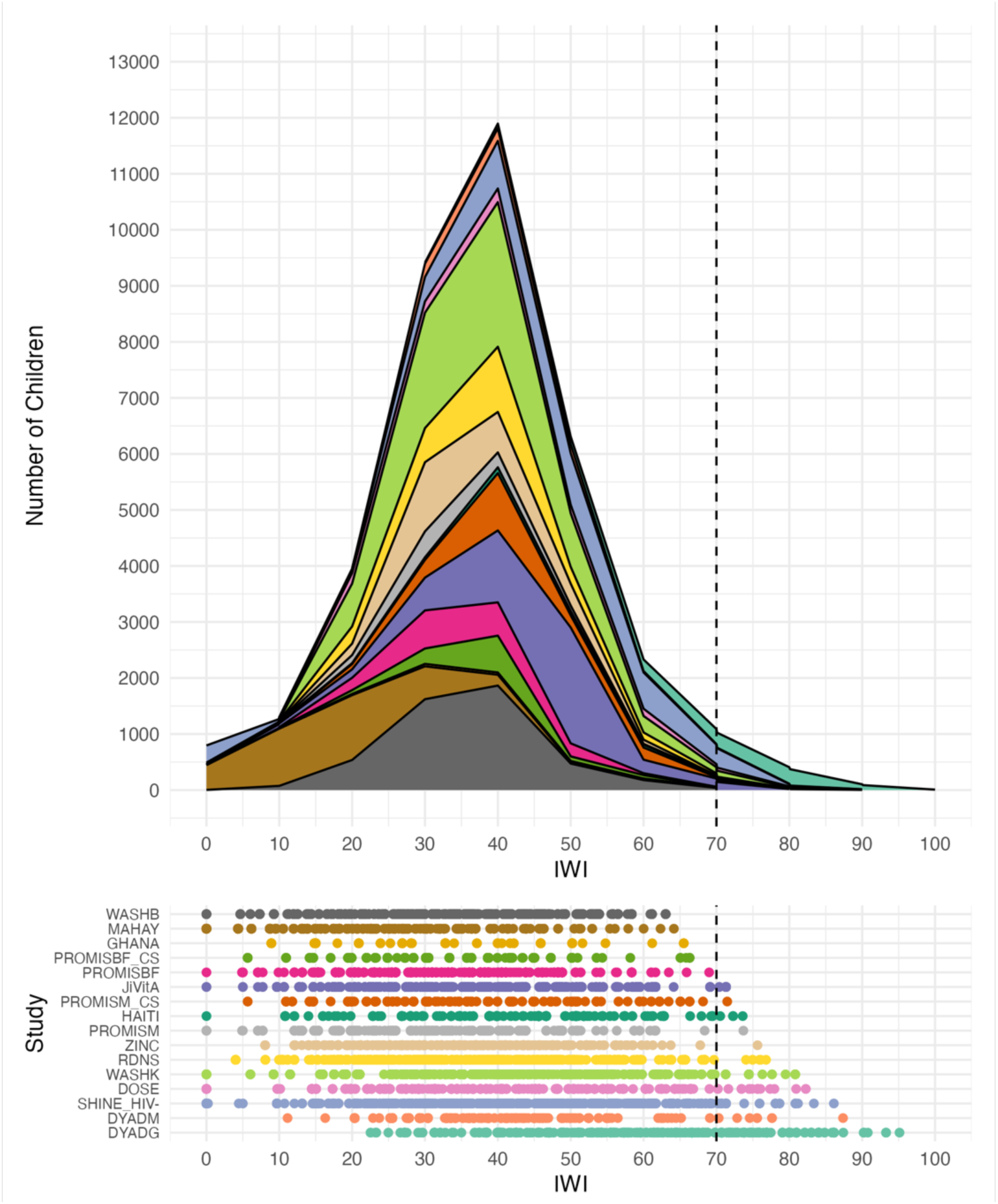
Distribution of the International Wealth Index by study. The International Wealth Index (IWI) is based on 12 household assets (i.e., 20 indicators in total) measured and weighted to provide a score between 0 (poorest) and 100 (wealthier). Overall, there were 37,605 children that contributed to the IWI distribution (excluding the missing values). The dashed line indicates an IWI value of 70. When fitting models to estimate effects, we included the whole range of the International Wealth Index (0-100), but predictions were made conditional on IWI values between 0 and 70, as few children had an IWI ≥70 (519 children from 11 studies, 1.4% of all measurements). For the stacked area plot, we binned the IWI values into intervals (bins) of 10 units, and calculated the number of children in each bin within each study.

### Measurement of child growth, development, and anemia

Growth outcomes were assessed as continuous measures, including the length-for-age z-score (LAZ) and weight-for-length z-score (WLZ), and as binary measures, such as stunting (LAZ < -2 standard deviation), wasting (WLZ < -2 SD), and severe stunting (LAZ < -3 SD). We excluded severe wasting (WLZ < -3 SD) as an outcome because of the very small number of events across the studies (**Fig. S11 and S12**). Development outcomes were evaluated using continuous scores for language, gross and fine motor skills, executive function, and socioemotional skills. The z-scores were standardized within each study by regressing the raw score on child age and sex, and calculating the standardized residuals with further details published previously^17^. Anemia outcomes were measured as a continuous outcome through blood hemoglobin concentration (g/L), and as a binary outcome defined as blood hemoglobin concentration < 110 g/L to define anemia.

Overall, children who received SQ-LNS showed better outcomes than those who did not in most studies, but substantial growth deficits remained based on WHO growth standards. Across the 14 studies, child growth, development, and anemia outcomes at endline varied substantially, with SQ-LNS groups generally showing modest improvements over control groups. Further details are provided in **Text S3**.

### Wealth inequalities in adverse child outcomes across studies

We hypothesized that children from wealthier households would have better growth, development, and anemia status compared to children from poorer households. To evaluate relative and absolute inequalities in adverse child outcomes (stunting, wasting, severe stunting, and anemia), we calculated the relative index of inequality (RII) and slope index of inequality (SII) for each binary outcome in each study, stratified by study arm. An RII > 1 (and 95% CI does not include 1) and an SII > 0 (and 95% CI does not include 0) indicate relative and absolute inequalities that favor wealthier households (i.e., adverse child outcomes were more prevalent among the poorer children). Conversely, an RII < 1 (and 95% CI does not include 1) and an SII < 0 (and 95% CI does not include 0) signify that these adverse outcomes are more prevalent among wealthier children. An RII with a 95% CI that includes 1 and an SII with a 95% CI that includes 0 reflect non-significant relative and absolute inequalities.

Overall, on a relative scale (**Fig. 2A**), we observed IWI-related inequalities in both study arms, with stunting, wasting, and severe stunting more prevalent among children born in the poorer households (RII and 95% CI > 1). In contrast, no significant inequalities were observed for anemia in either of the study arms (RII and 95% CI: 1). On the absolute scale (**Fig. 2B**), a similar pattern emerged, with absolute wealth inequalities in stunting, wasting, and severe stunting disproportionately affecting poorer participants (SII and 95% CI, > 0). Study-specific inequalities are detailed in **Text S4.**

**Fig. 2.**
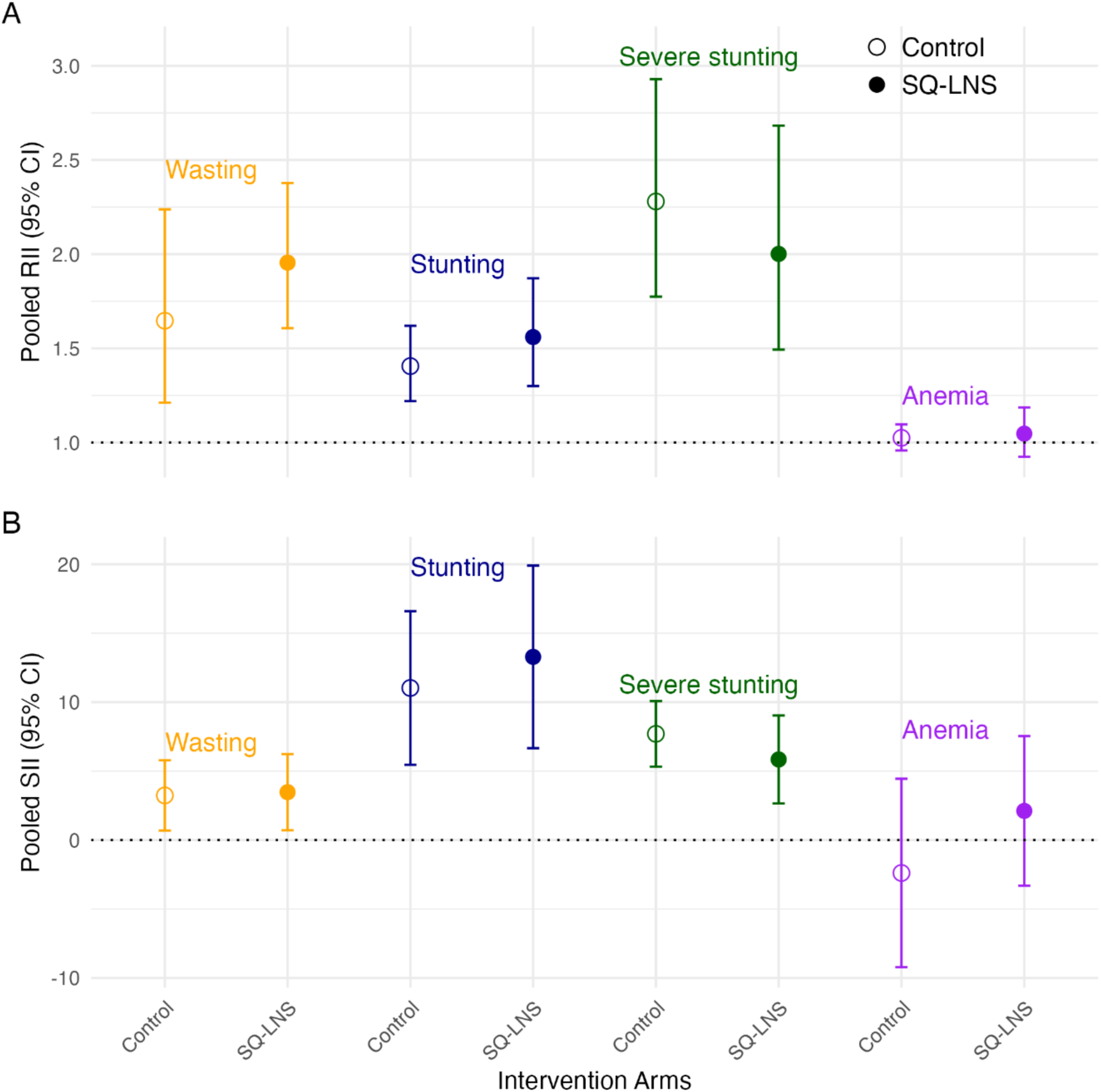
Pooled Relative Index of Inequality and Slope Index of Inequality across studies for stunting, wasting, severe stunting at endline and anemia between intervention arms. We restricted our analysis to binary outcomes for which the RII and SII are most clearly defined. The RII and SII quantify inequalities in relative and absolute scales, respectively. These are regression-based indicators which use all subgroups compared to a pairwise comparison that ignores other groups. To estimate these indices, we first ranked the individuals from the poorest to the wealthier in the cumulative distribution of the International Wealth Index deciles. A. The RII represents the ratio of the value at the bottom of the social hierarchy (poorest; intercept) to the value at the top (wealthier; slope + intercept). **B.** The SII represents the difference between these values. The SII point estimates and their 95% confidence intervals were multiplied by 100 to express them as percentage points. We used log-binomial regression to model the association between participants’ relative rank in the cumulative distribution of the IWI deciles and the child outcome, and modified Poisson if log-binomial models failed to converge (anemia). The RIIs and SIIs and their standard errors were pooled across studies using random-effects meta-analysis. An RII > 1 (and 95% CI does not include 1) and an SII > 0 (and 95% CI does not include 0) indicate relative and absolute inequalities in which adverse child outcomes were more prevalent among the poorest households. Conversely, an RII < 1 (and 95% CI does not include 1) and an SII < 0 (and 95% CI does not include 0) signify that these adverse outcomes were more common among the wealthier households. An RII (and 95% CI) including 1 and an SII (and 95% CI) including 0 reflect no significant relative and absolute inequalities. Intervention included maternal supplementation. Error bars represent 95% confidence intervals.

### Effect of SQ-LNS interventions across the wealth gradient

We hypothesized that the benefits due to SQ-LNS would be larger among children in the poorer households (lower levels of the IWI). SQ-LNS provides essential micronutrients that may be particularly beneficial for children in the poorer households, where growth and development deficiencies are more prevalent, offering greater potential to benefit. We estimated the mean outcomes by intervention group and continuous IWI in each study using splines in generalized additive models (**Fig. 3A**), and then pooled group-specific means (**Fig. 3B**) and differences (**Fig. 3C)** across studies using pointwise random-effects meta-analyses^40^. Spline fits for each study and child outcome are provided in the Supplementary Materials (**Text S1**).

**Fig. 3.**
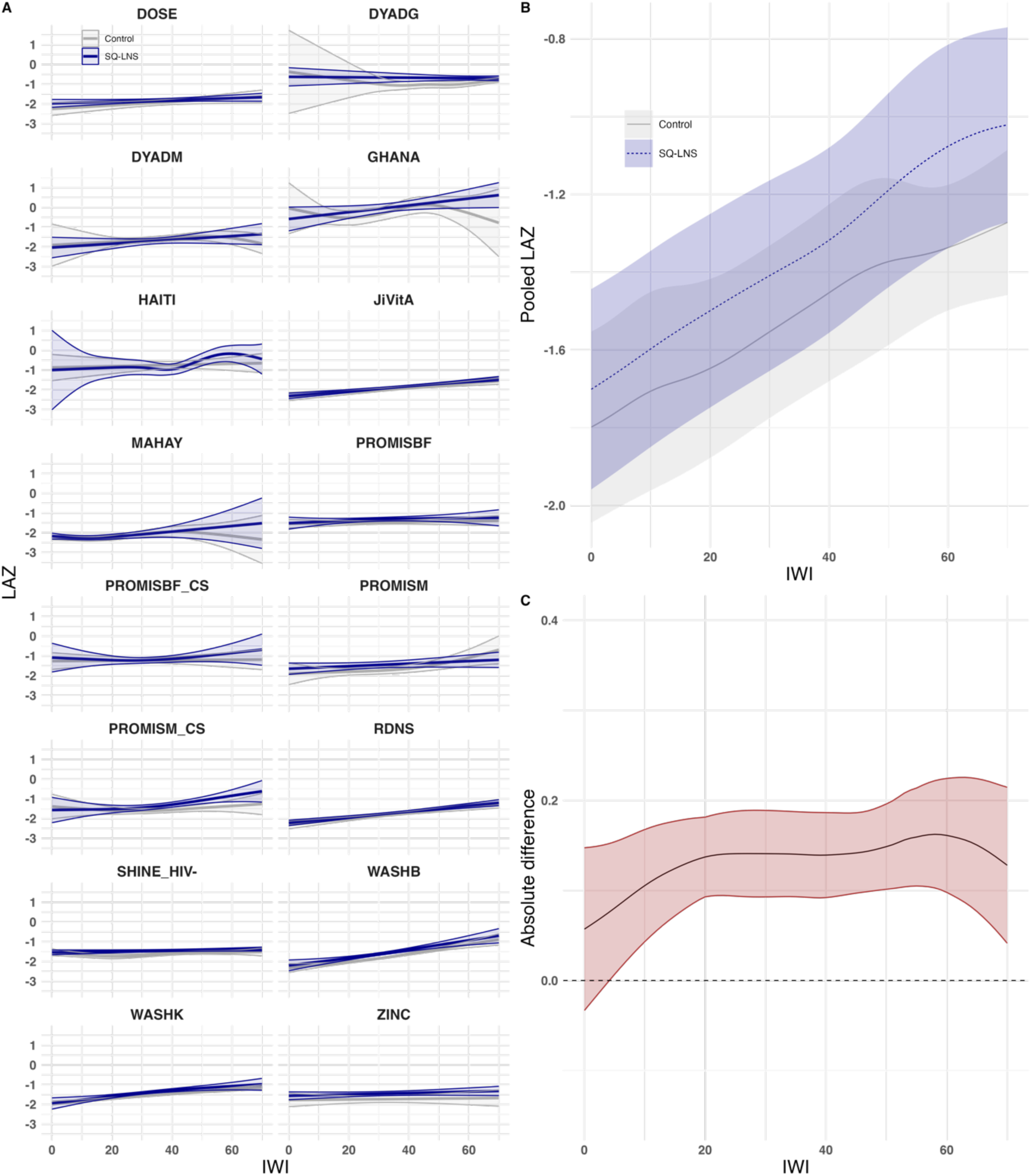
The analysis estimated group specific means and pooled differences by wealth index using point-wise meta-analyses. Example of the analysis process using length-for-age Z-score by International Wealth Index (IWI) and intervention group. **A.** Study-specific relationships between the IWI and length-for-age Z-scores (LAZ) by intervention group. **B.** Pooled estimates from a two-stage random effects meta-analysis of LAZ by IWI and intervention group across all studies in panel A. **C.** Pooled difference in LAZ between intervention groups over levels of IWI. Intervention effects conditional on IWI were estimated by subtracting the spline fits for the intervention group from the control group within each study. These effects were then pooled using pointwise random-effects meta-analysis with restricted maximum likelihood. In all panels, shaded bands represent 95% confidence intervals (CIs), while panels B and C specifically represent 95% pointwise CIs.

Overall, the SQ-LNS intervention significantly improved child growth outcomes, and the pooled mean LAZ and WLZ in children who received SQ-LNS were higher than those in the control group across all values of IWI (the estimates for stunting, wasting and severe stunting in SQ-LNS were generally below those of the control) (**Fig. 4**). The pooled differences between the SQ-LNS and control groups were generally constant across the IWI range (pooled interaction p-values >0.05). When accounting for the nonlinearity of the effects using splines, we observed point estimates suggesting greater SQ-LNS benefits on WLZ among children with higher IWI, with a similar but less pronounced pattern for LAZ and stunting, although the interaction p-values were not significant. Meanwhile, the effects of SQ-LNS on wasting and severe stunting showed no clear pattern across levels of IWI.

**Fig. 4.**
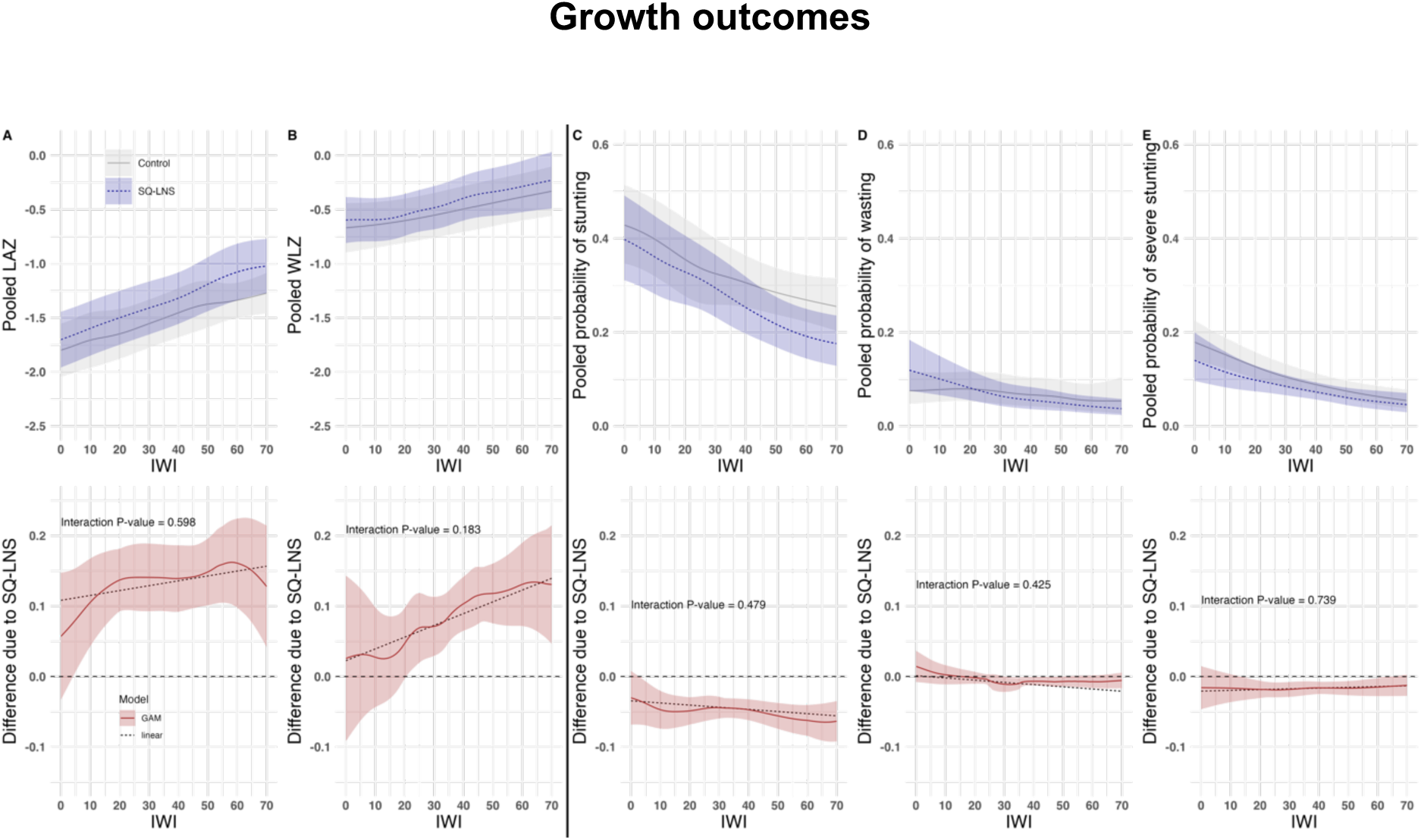
Pooled child growth outcomes by intervention group and differences due to SQ-LNS. A. Pooled length-for-age Z-score. **B.** Pooled weight-for-length Z-score. **C.** Pooled probability of stunting. **D.** Pooled probability of wasting. **E.** Pooled probability of severe stunting. We pooled the child growth outcomes for each intervention arm using pointwise random-effect meta-analysis with restricted maximum likelihood (REML). Intervention effects conditional on IWI were estimated by subtracting the spline fits for the intervention group from the control group within each study. These effects were then pooled using pointwise random-effects meta-analysis with REML. For the linear regression models (dashed lines), intervention effects were estimated by pooling the coefficients for the interaction term between IWI and intervention through random-effects meta-analysis and getting the difference between intervention groups. The pooled-p-for-interaction were from the random-effect meta-analysis of the pooled coefficients of interaction and standard errors across studies. Intervention included maternal supplementation. Shaded areas represent the 95% pointwise confidence intervals. The pooled estimates for length-for-age-Z-score are also noted in Fig. 3. Spline fits per study are found in the Supplementary Materials (Text S1).

We observed better language, gross motor, fine motor, and socioemotional scores among the children who received SQ-LNS, and the pooled mean outcomes were generally higher than those of the control group across the IWI range (**Fig. 5**). Pooled intervention effects did not differ significantly across the IWI range (pooled interaction p-values >0.05) except for socioemotional scores, for which there were increasing benefits as IWI increased (pooled interaction p-value = 0.046, excluded maternal SQ-LNS supplementation) (**Table S1, Fig. S21**).

**Fig. 5.**
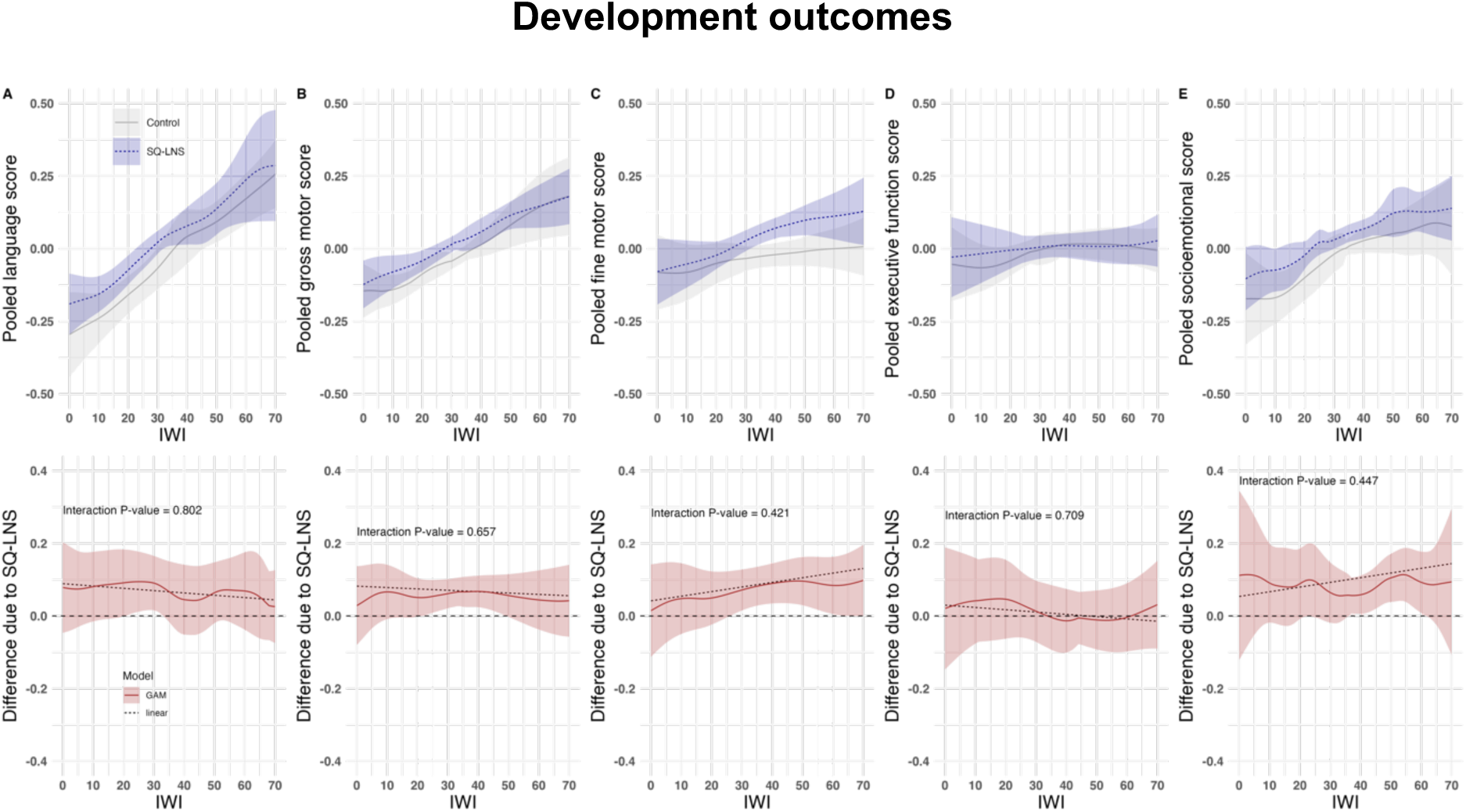
Pooled child developmental outcomes by intervention group and differences due to SQ-LNS. **A.** Pooled language score. **B.** Pooled gross motor score. **C.** Pooled fine motor score. **D.** Pooled executive function score. **E.** Pooled socioemotional score. We pooled the child development outcomes for each intervention arm using pointwise random-effect meta-analysis with restricted maximum likelihood (REML). Intervention effects conditional on IWI were estimated by subtracting the spline fits for the intervention group from the control group within each study. These effects were then pooled using pointwise random-effects meta-analysis with REML. For the linear regression models (dashed lines), intervention effects were estimated by pooling the coefficients for the interaction term between IWI and intervention through random-effects meta-analysis and getting the difference between intervention groups. The pooled-p-for-interaction were from the random-effect meta-analysis of the pooled coefficients of interaction and standard errors across studies. Intervention included maternal supplementation. Shaded areas represent the 95% pointwise confidence intervals. Development scores are internal to each study distribution of the study-specific indicator. Spline fits per study are found in the Supplementary Materials (Text S1).

In general, SQ-LNS significantly improved the mean hemoglobin concentration and reduced anemia prevalence (**Fig. 6**). Consistent with the findings for growth and most of the development outcomes, the pooled intervention effect did not vary by wealth (pooled interaction p-values >0.05).

**Fig. 6.**
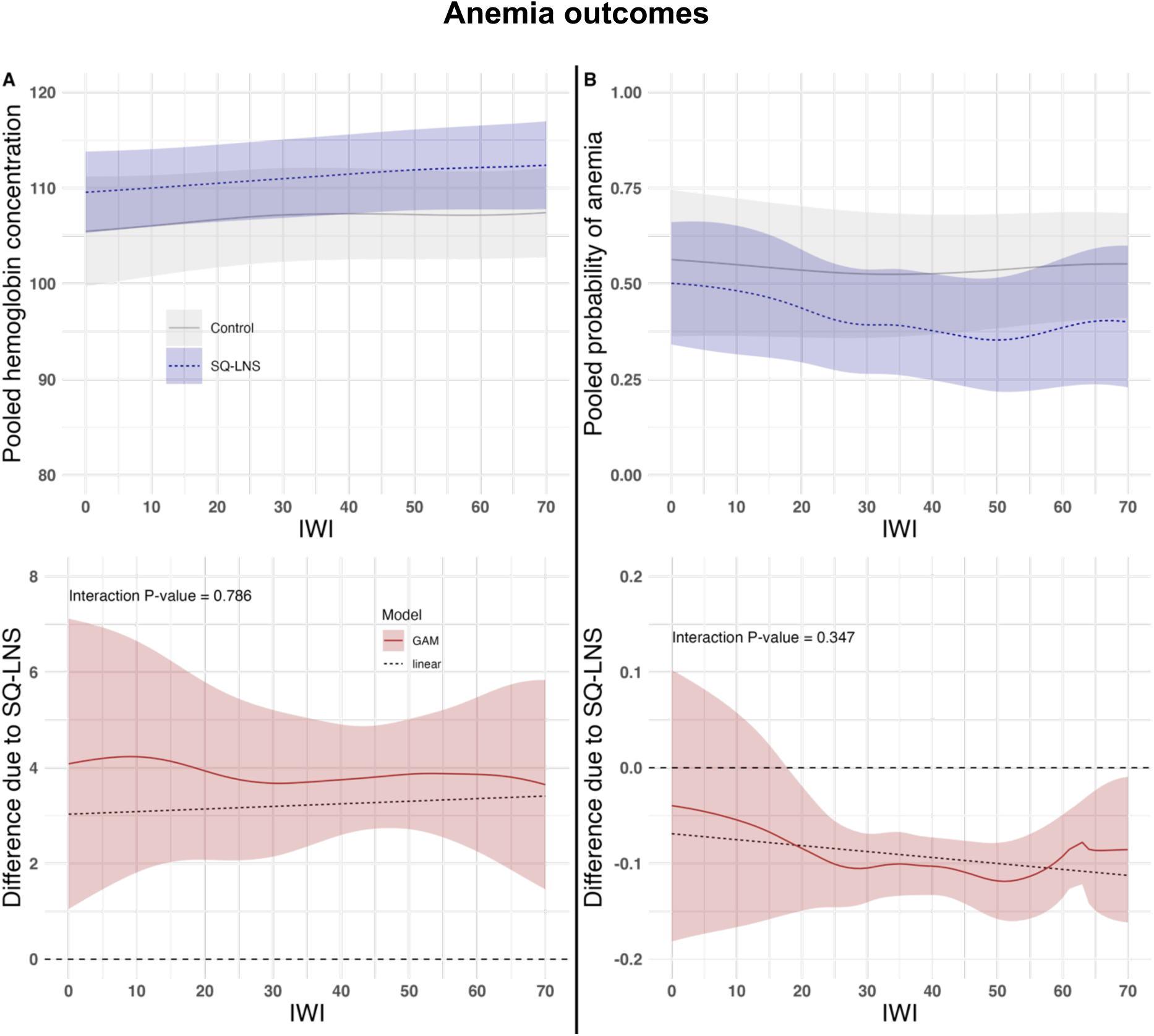
Pooled child anemia outcomes by intervention group and differences due to SQ-LNS. **A.** Pooled hemoglobin concentration. **B.** Pooled probability of anemia. We pooled the child hematological outcomes for each intervention arm using pointwise random-effect meta-analysis with restricted maximum likelihood (REML). Intervention effects conditional on IWI were estimated by subtracting the spline fits for the intervention group from the control group within each study. These effects were then pooled using pointwise random-effects meta-analysis with REML. For the linear regression models (dashed lines), intervention effects were estimated by pooling the coefficients for the interaction term between IWI and intervention through random-effects meta-analysis and getting the difference between intervention groups. The pooled-p-for-interaction were from the random-effect meta-analysis of the pooled coefficients of interaction and standard errors across studies. Intervention included maternal supplementation. Shaded areas represent the 95% pointwise confidence intervals. Spline fits per study are found in the Supplementary Materials (Text S1).

### Additional analysis using sex-stratified data

#### Wealth inequalities in adverse child outcomes across studies stratified by sex

We found relative and absolute wealth inequalities in stunting, wasting and severe stunting among both girls and boys, and these inequalities did not appear to differ by sex. No significant wealth inequalities were found for anemia for either girls or boys (**Text S5**).

#### Effect of SQ-LNS on different child outcomes by wealth stratified by sex

The SQ-LNS intervention significantly improved the growth, development, and anemia outcomes in both sexes (**Text S5**). Among girls, the pooled intervention effect between SQ-LNS and the control group did not vary by wealth for any child outcome (pooled interaction p-values >0.05). In contrast, among boys, the intervention effect varied significantly across the IWI range for WLZ (pooled interaction p-value = 0.048) and socioemotional scores (pooled interaction p-value = 0.020), indicating that boys from households with higher IWI had greater benefits from SQ-LNS.

### Additional analysis using maternal education as socioeconomic indicator

Since the wealth index captures an asset-related dimension of socioeconomic status, we used maternal education in an additional analysis (**Table S5**). Here, we used a categorical variable for maternal education: no formal education (none and incomplete primary), intermediate education (complete primary and incomplete secondary), and secondary and higher education. We observed results similar to those obtained using IWI. Further details are provided in **Text S6**.

## Discussion

This individual participant data meta-analysis demonstrated consistent benefits of SQ-LNS for child growth, development, and anemia outcomes across wealth levels. Despite the large gradients between the poorest and wealthier children in attained growth and development outcomes, SQ-LNS appears to benefit children similarly compared to controls across the wealth spectrum. These results underscore: (i) the large level of inequalities between the poorest and wealthier children in their growth and development in low-income communities in the trials, (ii) that SQ-LNS would benefit all children by a similar magnitude regardless of their current wealth in settings where children experience nutrition deficits, and (iii) that the magnitude of improvement due to SQ-LNS, while important and significant, is far smaller than the observed gradient between the poorest and wealthier children. It is important to note that these studies were conducted in largely poor, rural communities and so the wealth differential is within those limited contexts, which may in part be why the benefit is seen across the wealth spectrum. Our results imply that programs focusing on poverty alleviation; infant and young child feeding; water, sanitation, and handwashing; promotion of girls’ education and empowerment; maternal nutrition; and access to and quality of preventive and curative health cares are needed to help close the remaining gap.

Adherence to the intervention may vary by household wealth. If children from lower IWI households consumed smaller quantities of SQ-LNS, perhaps due to limited accessibility or greater sharing within the household, this could attenuate effects in lower wealth groups under an intent-to-treat analysis. However, in this study, we found that adherence levels were generally similar across the IWI spectrum (**Fig. S42**). This suggests that the similar magnitude of effects of SQ-LNS at lower and higher IWI levels is not explained by differences in adherence across IWI levels.

We observed that SQ-LNS improved a child’s growth and development to a level comparable to that of a child in a higher wealth bracket without changing any other element of household wealth. For example, in the case of LAZ (**Fig. 3B**), the mean LAZ for an IWI of 20 was approximately –1.5 in the SQ-LNS group. A similar mean LAZ was observed in the control group at an IWI of approximately 40. An increase from 20 to 40 IWI represents a 50-percentage point increase in the wealth distribution of the study population. Achieving such a population-level shift in wealth would require complex, multifaceted interventions and policy reforms over a timescale of years to decades. Wealth, however, is only one component of a complex web of social and economic conditions that influence health across the life course. As such, increasing household wealth does not necessarily guarantee better child outcomes. Rather, wealth likely serves as a proxy for other underlying factors that more directly influence child health. In this context, SQ-LNS may provide a more immediate and potentially generalizable solution to directly improve most of the child growth and development outcomes in the near term. This result reinforces the potential of scaling up SQ-LNS as an intervention to improve child health outcomes in the Global South, helping to converge the health gap between poorer, healthier, and wealthier countries^41^.

Although the intervention improved outcomes, this did not mitigate the wealth differential since the inequality measures in child outcomes were very similar in the intervention and control groups (**Fig. 2**) — the latter may serve as a proxy for pre-SQ-LNS status. Inequities may persist even within relatively poor communities. For example, an IWI of 0-20 compared to 61-70 represents a substantial difference in living conditions, and disparities in child outcomes between these wealth levels are both avoidable and unjust if left unaddressed thus making them inequitable. Complementary interventions are needed that can further enhance child outcomes, while addressing inequities that SQ-LNS alone cannot overcome. For example, SQ-LNS can be integrated with complementary interventions such as vaccination, frequent screening for wasting, and others^31,32,42^. Given its economic value, SQ-LNS can serve as both a nutritional intervention and an incentive to encourage participation in these programmatic components to reduce costs and create a synergistic effect, especially if universal implementation is unsustainable. Evidence from several case studies has shown that SQ-LNS provision can be highly cost-effective if well implemented^43^, particularly when delivered alongside behavior change communication (BCC) and screening. One study found that integrating SQ-LNS with BCC and screening reduced the unit cost per child contact for these services by more than the unit cost of SQ-LNS itself^44^. This suggests that SQ-LNS can serve as an effective incentive to increase uptake and efficiency of BCC and screening, compared to delivering BCC and screening alone, which was associated with significantly higher unit costs. In the PROMIS studies, SQ-LNS was offered alongside wasting screening to ensure that more children with severe or moderate acute malnutrition would be identified and referred to treatment services faster, creating a possible synergistic effect between prevention and screening/ treatment^31,32^. Another example that is following a similar approach is the NutriVax Project which has integrated SQ-LNS with vaccination programs^42^. Vaccination not only protects against infections that impair growth and development^45^ but can also serve as a touchpoint for reaching vulnerable populations with nutritional supplementation.

Another critical opportunity is integration within climate-resilient nutrition programs, e.g., targeted implementation of SQ-LNS during periods of heightened vulnerability, such as the hunger season or in regions experiencing food insecurity. Climate change is already threatening global food systems as rising temperatures, extreme weather events, and increased frequency of natural disasters are reducing crop yields, lowering nutritional quality of foods, and limiting access to diverse and sufficient diets, particularly for poor and vulnerable populations^46^. It is projected that, between 2024 and 2050, climate change will result in 40 million additional children with stunting and 28 million additional cases of wasting^1^.

Overall, girls showed better growth outcomes than boys (**Fig. S31-S33**), which is consistent with a previous analysis of this data^16^. Some studies suggest that boys may be more biologically vulnerable than girls to adverse conditions in early life, which could limit their response to nutrition interventions^48,49^. For both sexes, we found no significant interaction between IWI and SQ-LNS for most of the outcomes. However, in boys, the impact of SQ-LNS on WLZ (**Fig. S39**) and socioemotional scores (**Fig. S40**) varied significantly by IWI, with children from households with higher IWI generally experiencing greater benefits. We also found that the overall effect of SQ-LNS (excluding maternal supplementation) on children’s socioemotional scores increased with wealth. While both the linear regression line and spline showed a clear steep trend for WLZ, this pattern was less distinct for socioemotional scores, warranting caution in interpretation. Although boys’ health outcomes have been linked to their mothers’ diet and socioeconomic status^49^, and some evidence suggests preferential care-seeking for boys in wealthier families in some contexts compared to poorer households^50^, the underlying mechanisms driving these patterns remain unclear.

Consistent with prior findings, our results indicate that wealth does not significantly modify the effects of SQ-LNS on growth^16^ or anemia^18^. However, previous analyses indicated significant effect modification by wealth for language, motor, and executive function scores, where greater effects of SQ-LNS were found among poorer households^17^. This discrepancy may be attributed to differences in the wealth indicators used. While the previous analyses relied on above- or below-median household wealth, this study used a continuous, standardized wealth index, which captures more nuanced, non-linear variations in effects across the entire wealth continuum. The same study also found no significant interaction between wealth and the binary development outcomes.

The current study had several limitations. First, in constructing the IWI, we used a formula adapted for three missing asset variables to account for studies with >3 missing variables which may have under-estimated the wealth scores. In a subset of studies, we also used the number of rooms in a household instead of the number of sleeping rooms in the construction of the IWI due to the limited data for the latter which may have led to an overestimation of wealth. Additionally, most trials were conducted in sub-Saharan Africa, with only three trials from Asia and one from the Caribbean, limiting the generalizability of the findings to other regions. Also, the majority of the trials were performed in relatively low-income communities within each country, thus limiting generalizability to middle- and higher-income communities in those countries. Development and anemia outcomes were not available in some studies, reducing the statistical power for some analyses. Using wasting prevalence as an indicator may not fully capture the dynamic nature of the condition, and thus we may have missed important inequalities in wasting patterns.

Despite these limitations, the present study had several strengths. It included a large sample of high-quality RCTs across 14 diverse sites with varying geographic, socioeconomic, and health profiles. The included trials represent a mix of efficacy and effectiveness studies, which may have captured effects under both ideal and real-world conditions. The use of a standardized IWI enhanced the comparability of wealth measures across studies, thereby improving the consistency and reliability of our analyses. Removing one or even two assets from the index was found to have minimal impact on household rankings, and excluding data from specific regions in the developing world did not significantly alter results^39^. The IWI also showed strong correlations with national Demographic and Health Surveys wealth indices, further supporting its validity^39^. It was also shown to be a strong predictor of child outcomes such as height-for-age z-scores and infant mortality^51^. Using maternal education instead of IWI gave similar results except for the hemoglobin concentration outcome, where we found a significant interaction (maternal education × intervention) p-value using the one-stage meta-analysis (vs. when using a two-stage meta-analysis for each factor level). However, this should be interpreted with caution, as interaction terms in one-stage mixed-effects models may be susceptible to ecological bias, conflating within-study and cross-study effects^40,52^. Additionally, the 95% CIs for the two pooled interaction coefficients (intermediate × SQ-LNS and Secondary/Higher × SQ-LNS) derived from the two-stage meta-analysis overlapped, indicating no significant interactions.

In conclusion, we highlight large inequalities in growth and development outcomes between the poorest and wealthier children engaged in SQ-LNS trials across multiple countries, as well as the potential of SQ-LNS to benefit children across the wealth spectrum. While SQ-LNS demonstrated benefits in both low and higher wealth categories, these equal benefits were not sufficient to reduce existing inequities in child outcomes. The mean impact of SQ-LNS is far smaller than the magnitude of existing inequalities across wealth strata, underscoring the need for additional complementary interventions. The findings also imply that if decision makers target an SQ-LNS intervention to the most disadvantaged populations, they should expect an impact on most child outcomes comparable to what is observed among wealthier populations not provided with the intervention. This study emphasizes the usefulness of incorporating an equity lens when evaluating intervention effects to ensure that “no child is left behind.”

## Methods

### Individual participant data

We utilized individual participant data previously collated for a prior analysis^22^, encompassing 14 prospective studies of small-quantity lipid-based nutrient supplements (SQ-LNS) provided to children aged 6-24 months. Eligible trials were individual or cluster randomized controlled trials (RCTs) conducted in the Global South, featuring longitudinal or repeated cross-sectional data collection. The DOSE trial used a single set of randomization blocks as a technique for individual level randomization^25^. Trials provided SQ-LNS (<125 kcal/day) for at least three months during the age range of 6-24 months and reported at least one relevant outcome.

The majority of trials initiated child SQ-LNS at 6 months of age, with a duration of 6–12 months, except for DYAD-G and DYAD-M, which also provided maternal SQ-LNS^23–36^. RDNS and MAHAY included arms with both child and maternal SQ-LNS as well as an arm with child SQ-LNS alone. Children were eligible if their age at baseline allowed them to receive the intervention (supplementation or control components) for at least 3 months between 6 and 24 months of age (i.e. children who entered the study at >21 months of age were excluded from the analyses). Complete details of the individual participant data have been published elsewhere^22^. The SHINE trial was specifically designed to present the results separately for HIV-exposed and HIV-unexposed children; only HIV-unexposed children were included in this analysis. The two PROMIS trials in Burkina Faso and Mali included independent longitudinal cohorts and repeated cross-sectional samples. Thus, we present longitudinal and repeated cross-sectional findings as separate comparisons for each trial. The number of studies included in each analysis depended on the availability of the outcome of interest.

The exclusion criteria were as follows: 1) trials limited to children with moderate or severe malnutrition where LNS was used for treatment rather than prevention; 2) trials conducted in hospitalized populations or among children with pre-existing diseases; or 3) trials where SQ-LNS was combined with additional food or nutrients within a single arm without a comparator group isolating the SQ-LNS effect.

Trials with multiple SQ-LNS interventions (e.g., varying dosages or formulations), those combining child SQ-LNS with maternal LNS supplementation, or those including non-nutritional interventions (e.g., water, sanitation and handwashing) were included. Study arms involving non-LNS child supplementation (e.g., multiple micronutrient powders or fortified blended food) were excluded.

We conducted a complete case and intention-to-treat analysis.

### Defining the intervention

All arms providing SQ-LNS were combined into one intervention group (SQ-LNS), whereas non-LNS arms (no LNS for mother or child) were grouped into a single comparator group (hereafter, control).

Study arms that provided maternal SQ-LNS supplementation in addition to child SQ-LNS (**Table 1**) were excluded from the primary analysis but were included in the secondary analysis. This distinction was made because maternal SQ-LNS supplementation as compared to iron and folic acid supplementation has been shown to independently benefit infant and young child growth by improving birth weight and birth length as well as reducing the incidence of small-for-gestational-age births and newborn stunting^53^. However, the results of the primary analysis were consistent with those of maternal sensitivity analysis (**Text S1**). To maximize the study inclusion and participant sample size, in alignment with previous individual participant data analyses, we present the findings from the secondary analysis as the main results.

### Defining the outcomes of interest

The outcomes of interest in this study included growth, development, and anemia measures at the endline. Endline was defined as the main post-intervention time point for trials with less frequent child assessments or as the age nearest to the end of the supplementation period for trials with monthly assessments^16^. Growth outcomes were assessed continuously, using length-for-age z-scores (LAZ) and weight-for-length z-scores (WLZ), and as binary indicators, including stunting (LAZ < -2 standard deviation), wasting (WLZ < -2 SD), and severe stunting (LAZ < -3 SD). We excluded severe wasting (WLZ < -3 SD) due to very small number of events across studies (**Fig. S11 and S12)**. Development outcomes were evaluated using continuous scores for language, motor function (gross and fine), executive function, and socioemotional scores. The z-scores were standardized within each study by regressing the raw development scores on child age and sex to obtain standardized residuals, analogous to LAZ in representing deviations from the mean for a given age and sex in SD units, but calculated relative to within-study distribution rather than external standard^17^. The measures and tools used have been described and published elsewhere^17^. For anemia, hemoglobin concentration (g/L) was measured as a continuous variable, whereas anemia status was classified as a binary outcome (hemoglobin < 110 g/L).

### Defining the effect modifier

The International Wealth Index (IWI) was first introduced by the Global Data Lab (https://globaldatalab.org/iwi/computing/) as a composite measure of household wealth using 12 asset-based variables^39^. These variables reflect the ownership of consumer goods (such as a TV, refrigerator, phone, car, bicycle), cheap items (e.g., basic furniture such as chairs), expensive items (e.g., air conditioners), household characteristics (including flooring material, toilet facilities, and number of rooms), and access to public utilities such as electricity and water.

In total, the IWI consists of 20 indicators: eight related to ownership of consumer durables and electricity access; nine (grouped into three sets) assessing the quality of the water source, flooring material, and toilet facilities; and three measuring the number of rooms in the household. In constructing the IWI, we collated the 12 asset-based variables for each study and recorded the variables using a similar categorization used by the Global Data Lab to harmonize them. We then ranked the households on the IWI scale, and its values for the 20 indicators were included in the following formula:

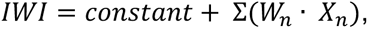

where *X_n_* is the asset indicator variable of the n^th^ asset and *W_n_* is the estimated IWI weight of the nth asset. The weights were derived through principal component analysis of these 12 common assets and indicators in a pooled database of 165 household surveys from 1996 to 2011 in 97 countries in the Global South, covering 2.1 million households^39^. The IWI scores ranged from 0 to 100. An IWI value of 0 is assigned to households lacking durable assets, no access to public services, and the lowest-quality housing. By contrast, a value of 100 is given to households that own all durable assets, have the best public utilities (such as electricity and water), and live in the highest-quality housing.

The Global Data Lab tested its reliability as a comparable measure of household wealth^39^. Their findings indicate that removing one or even two assets from the index has a minimal impact on household rankings. Similarly, excluding data from specific regions in the developing world did not significantly alter the results. Within Demographic and Health Survey (DHS) countries, the IWI demonstrated high correlations with the national DHS wealth indices^39^. Additionally, their analysis showed that the 2.1 million households included in the dataset were evenly distributed across the IWI scale, avoiding issues such as clumping or truncation^39^.

Some studies had missing asset variables for which we used separate formulas used by the Global Data Lab based on different combination of missing variable(s). Missing asset values were treated as zero, and the constants were adjusted based on the available variables.

Additionally, certain asset variables did not include the full range of quality levels (treated as dummy or indicator variables) (**Table S2B**). For instance, in JiVitA-4, the water source quality included only “low” and “medium” levels, with no “high” level, as defined by the Global Data Lab. In contrast, RDNS lacked a “low” quality level. We found that the IWI and study-specific wealth indices were highly correlated in the majority of studies (**Figure S27**). Therefore, we opted to use IWI in the main analysis.

We calculated the median and SD of the IWI and assessed the distribution of assets and indicators used in its construction by determining the number and percentage of participants owning each, stratified by study arm. Additionally, we evaluated asset ownership and indicator distribution across the IWI tertiles to explore the wealth gradient. We then plotted the distribution of IWI by study type.

### Statistical Analyses

#### Descriptive Statistics

Before addressing these aims, we first assessed the mean child growth, development, and anemia measures attained at endline between study arms for each study.

#### Assessing wealth inequalities in different adverse child outcomes across studies

We estimated the relative and absolute inequalities for each binary adverse outcome in each study, stratified by study arms, by calculating the relative index of inequality (RII) and slope index of inequality (SII)^54^. These are regression-based indicators that are commonly used to measure health inequalities. Relative inequalities are typically greater when the overall prevalence of an outcome is low, whereas absolute inequalities are more pronounced at intermediate prevalence levels^55^. Reporting both relative and absolute inequalities is crucial as conclusions may differ based on the scales chosen.

We restricted our analysis to binary adverse outcomes because the formulas for these indicators were specifically designed to accommodate binary data. The RII represents the ratio of the value at the bottom of the socioeconomic status (intercept) to the value at the top (slope + intercept), while the SII represents the difference between these values for binary outcomes. We used log-binomial regression for binary outcomes to model the association between participants’ relative ranks in the cumulative distribution of the IWI deciles and each adverse child outcome. For cluster-randomized studies, clustering was accounted for by using robust standard errors. If log-binomial models failed to converge (for anemia), we used modified Poisson regression with robust standard errors as an alternative. The Delta method was used to calculate the 95% CIs for the SII, while the Wald-type 95% CI was estimated for the RII.

Second, we pooled the RIIs and SIIs and their standard errors across studies using a random-effects meta-analysis to obtain pooled estimates and their 95% confidence intervals using the inverse variance method.

An RII > 1 (and 95% CI does not include 1) and an SII > 0 (and 95% CI does not include 0) indicate relative and absolute inequalities, respectively, where adverse child outcomes are more prevalent among children born in the poorest households. Conversely, an RII < 1 (and 95% CI does not include 1) and an SII < 0 (and 95% CI does not include 0) signify that these adverse outcomes are more common among wealthier households. An RII (and 95% CI) including 1 and an SII (and 95% CI) including 0 reflect non-significant relative and absolute inequalities.

#### Estimating the effect of SQ-LNS on different child outcomes by wealth

Following the original individual participant data meta-analyses, we used a two-stage hierarchical approach: a) first, we estimated the study-level effects derived from individual-level data for each trial; and b) second, we pooled estimates across the studies using a random-effect meta-analysis of the individual study spline fits.

To do this, we used pointwise meta-analysis^40^. In this approach, the predicted outcomes from the first stage are aggregated rather than regression coefficients. We aggregated the predicted outcomes for each value of X (i.e., IWI). We followed the approach of Belias et al. in conducting a two-stage pointwise meta-analysis^40^.

#### First stage

To assess the effect of the intervention on child outcomes by continuous IWI, we fitted a generalized additive model (GAM)^56^ to capture potential nonlinear relationships between IWI and each child outcome. A common approach to investigating intervention effect modification is to model the interaction between a potential effect modifier and intervention^40^. In our analysis, we modeled the relationship between the effect modifier and outcome by including the spline-transformed modifier both as a main effect and in interaction with the intervention. We used the mgcv package in R to implement the GAM, which represents smooth functions using penalized regression splines (specifically, the cubic regression spline), with basis functions designed to be optimal^57^.

To avoid overfitting, we used a nonparametric smoother, the cubic regression spline^58^, fitting the model using restricted maximum likelihood (REML). We accounted for clustering in the cluster-randomized studies by incorporating cluster-level random effects. We applied the Gaussian family for continuous outcomes and the binomial family with a logit link for binary outcomes. Outcomes were estimated as a function of the IWI for each study and study arm, with the absolute risk difference and confidence intervals estimated per study across the IWI range. For binary outcomes, we back-transformed the predicted outcomes and absolute risk differences using an inverse logit function. Predictions were made conditional on IWI values between 0 and 70, because very few samples had an IWI of 70 or higher (**Fig. 1**).

#### Second stage

In the second stage, we pooled both the predicted outcome curves for each study arm (on the identity scale for continuous outcomes and the logit scale for binary outcomes) and their absolute risk differences. This was performed using pointwise random-effects meta-analyses with REML estimators (and DerSimonian-Laird estimator when the model failed to converge). We then back-transformed the predicted binary outcomes using the inverse logit function.

#### Pooled interaction p-value

To assess whether the pooled intervention effect between the SQ-LNS and control arms (i.e., difference due to SQ-LNS) was statistically different by IWI, we estimated the pooled p-value for the interaction.

First, for interpretability and simplicity, we fitted linear regressions to estimate the effect of SQ-LNS (versus control) on each child outcome (instead of using GAMs), including the interaction term between SQ-LNS (versus control) and IWI. For cluster-randomized studies, we accounted for clustering using robust standard errors, implemented via the lm_robust function from the estimatr package in R^59^.

Second, we extracted the coefficients and standard errors of the interaction terms from each study. These data were pooled using a random-effects meta-analysis with REML. Based on the pooled coefficients, we predicted child outcomes across the range of IWI (0-70) for both the SQ-LNS and control groups.

We calculated the pooled point estimates for the difference due to SQ-LNS in each child outcome across the range of IWI (i.e., 0-70) as follows:

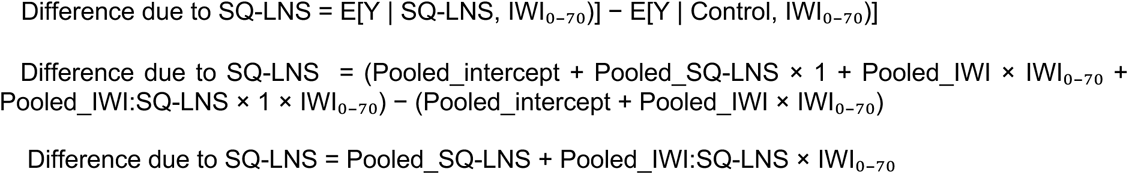

Finally, the pooled interaction p-value was derived from the random-effects meta-analysis, which represented the p-value for the pooled intervention effect.

#### Sex-stratified analyses

To account for potential differences in the effects of SQ-LNS on each child outcome based on sex, we stratified the data by child sex before estimating wealth inequalities in adverse child outcomes and the effects of SQ-LNS on child outcomes based on IWI. The analyses were repeated separately for girls and boys.

#### Additional analysis using maternal education as socioeconomic indicator

Since the wealth index captures a specific dimension of socioeconomic status, we used maternal education in an additional analysis. Maternal education is considered a reliable predictor of both short- and long-term child health and well-being. We used the categorical variable of maternal education: no formal education (none and incomplete primary), intermediate (complete primary and incomplete secondary), and secondary and higher education. We assessed the interaction with the SQ-LNS intervention in two ways: (i) comparing models with and without the interaction term (maternal education × intervention) by fitting the models through a one-stage meta-analysis using the pooled data using Wald-type F test; and ii) fitting the model with interaction (maternal education × intervention) for each study and pooling each level of the interaction coefficients across studies for each child outcome using a random-effects meta-analysis.

## Inclusion and Ethics

The original IPD meta-analysis protocol was registered with PROSPERO CRD42019146592 (https://www.crd.york.ac.uk/prospero) and approved by the institutional review board of the University of California, Davis (1463609-1). Amendments to the protocol were approved by the same board (1463609-4). All individual trial protocols were approved by the respective institutional ethics committees.

## Data availability

The pre-analysis plan is available through the Open Science Framework (OSF, https://osf.io/c7f5g/)^60^. The data will not be made available because they were compiled from 14 different trials, with access controlled by the investigators of each trial.

## Code availability

All analyses were conducted in R (version 4.3.3 “Angel Food Cake”). Codes and instructions to reproduce all analyses are available in the Open Science Framework (OSF, https://osf.io/c7f5g/)^60^.

## Acknowledgements

The SQ-LNS individual participant data meta-analysis project was funded by the Bill and Melinda Gates Foundation to the University of California, Davis (OPP49817). This study also had additional support from the National Institute of Allergy and Infectious Diseases (R01AI166671 and R03AI188012 to B.F.A.).

## Author contributions

P.A.A.-T. and B.F.A developed and drafted the statistical analysis plan with input from K.G.D., C.P.S., C.D.A., K.R.W., L.H., S.P.L., E.G., T.B., A.J.P., and P.C.

P.A.A.-T. conducted the data analysis with input and guidance from B.F.A.

P.A.A-T. constructed the tables and figures with input and guidance from B.F.A.; and input from K.G.D., C.D.A., C.P.S., and K.R.W.

K.R.W. and C.D.A. collated the data from all trials during the previous individual participant data meta-analysis. P.A.A.-T, C.D.A., and B.F.A. collected additional asset variables from relevant trials. Investigators from JiVitA-4, PROMIS-BF, iLiNS-Zinc, Ghana, DYAD-G, iLiNS-DOSE, PROMIS-Mali, and SHINE provided additional asset data for the construction of the international wealth index.

C.D.A. provided data resources. B.F.A. and C.D.A. provided statistical resources.

B.F.A. and K.G.D. provided guidance and supervision in the evolution of this study.

P.A.A.-T. wrote the initial draft of this manuscript with input and conceptual guidance from B.F.A.; and K.G.D., C.P.S., C.D.A., and K.R.W., helped in the interpretation of the preliminary results.

All authors contributed to the interpretation and subsequent revisions.

All authors read and approved the manuscript.

