## Supplementary Material for "Assessing equity in effects of nutritional supplementation for child growth, development, and anemia"

TEXT S1. CHILD GROWTH, DEVELOPMENT AND ANEMIA OUTCOMES CONDITIONAL ON INTERNATIONAL WEALTH INDEX PER INTERVENTION ARM AND STUDY. 2

TEXT S2. INTERNATIONAL WEALTH INDEX. 30

TEXT S3. DESCRIPTIVE STATISTICS OF CHILD OUTCOMES. 42

TEXT S4. STUDY-SPECIFIC WEALTH INEQUALITIES IN ADVERSE CHILD OUTCOMES. 47

TEXT S5. SEX-STRATIFIED ANALYSES. 53

TEXT S6. MATERNAL EDUCATION AS SOCIOECONOMIC INDICATOR. 65

TEXT S7. ADHERENCE LEVEL BY INTERNATIONAL WEALTH INDEX. 67

This supplemetary material is divided into **Text S1, Text S2, Text S3, Text S4, Text S5, Text S6** and **Text S7**. Supplementary tables and figures can be found in each Text.

#### Text S1. Child growth, development and anemia outcomes conditional on international wealth index per intervention arm and study.

**Figures S1-S12** show the spline fits for each study and child growth outcomes (length-for-age z-score, weight-for-length z-score, stunting, wasting, severe stunting and severe wasting). **Figures S13-S22** show the spline fits for each study and child development outcomes (language score, gross motor score, fine motor score, executive function score and socioemotional score). **Figures S23-S26** show the spline fits for each study and child anemia (hemoglobin concentration and anemia).

Each outcome has one result for the primary analysis (i.e., excluding maternal SQ-LNS supplementation) and one result for the secondary analysis (i.e., including maternal SQ-LNS supplementation). We found the findings in the secondary analysis consistent with the findings of the primary analysis. Thus, we presented the results from the secondary analysis in the main text.

**Table S1. Pooled interaction p-value for each child growth, development and anemia indicators.** They were derived from the random-effect meta-analysis of the pooled coefficients of interaction between the International Wealth Index and the intervention, and standard errors across studies. LAZ: length-for-age z-score, WLZ: weight-for-length z-score.

| Outcome | Pooled interaction p-value (excluded maternal supplementation) | Pooled interaction p-value (included maternal supplementation) |
|---|---|---|
| **Growth** | | |
| **LAZ** | 0.404 | 0.598 |
| **WLZ** | 0.096 | 0.183 |
| **Stunting** | 0.535 | 0.479 |
| **Wasting** | 0.404 | 0.425 |
| **Severe stunting** | 0.560 | 0.739 |
| **Development** | | |
| **Language score** | 0.742 | 0.802 |
| **Gross motor score** | 0.949 | 0.657 |
| **Fine motor score** | 0.543 | 0.421 |
| **Executive function score** | 0.492 | 0.709 |
| **Socioemotional score** | 0.046 | 0.447 |
| **Anemia** | | |
| **Hemoglobin** | 0.382 | 0.786 |
| **Anemia** | 0.430 | 0.347 |

**Figure S1. Length-for-age Z-score by international wealth index and intervention group. A.** Study-specific relationships between the International Wealth Index (IWI) and Length-for-age Z-scores (LAZ) by intervention group. **B.** Pooled estimates from a two-stage random effects meta-analysis of LAZ by IWI and intervention group across all studies in panel A. **C.** Pooled difference in LAZ between groups over levels of IWI. Intervention effects conditional on IWI were estimated by subtracting the spline fits for the treated group from the control group within each study. These effects were then pooled using pointwise random-effects meta-analysis with Restricted Maximum Likelihood. In all panels, shaded bands represent pointwise 95% confidence intervals. Intervention excluded maternal supplementation.

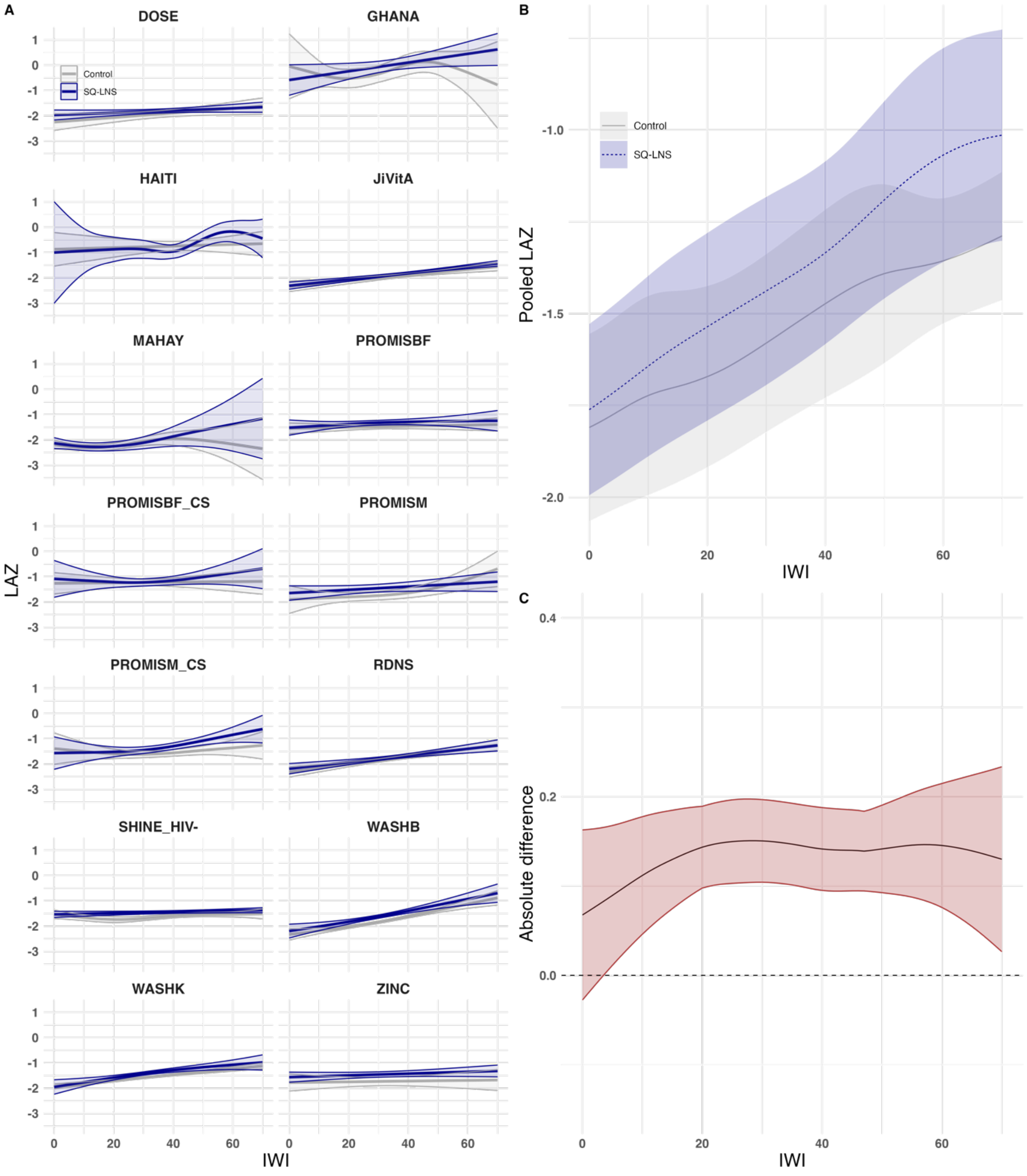

**Figure S2. Length-for-age Z-score by international wealth index and intervention group. A.** Study-specific relationships between the International Wealth Index (IWI) and Length-for-age Z-scores (LAZ) by intervention group. **B.** Pooled estimates from a two-stage random effects meta-analysis of LAZ by IWI and intervention group across all studies in panel A. **C.** Pooled difference in LAZ between groups over levels of IWI. Intervention effects conditional on IWI were estimated by subtracting the spline fits for the treated group from the control group within each study. These effects were then pooled using pointwise random-effects meta-analysis with Restricted Maximum Likelihood. In all panels, shaded bands represent pointwise 95% confidence intervals. Intervention included maternal supplementation.

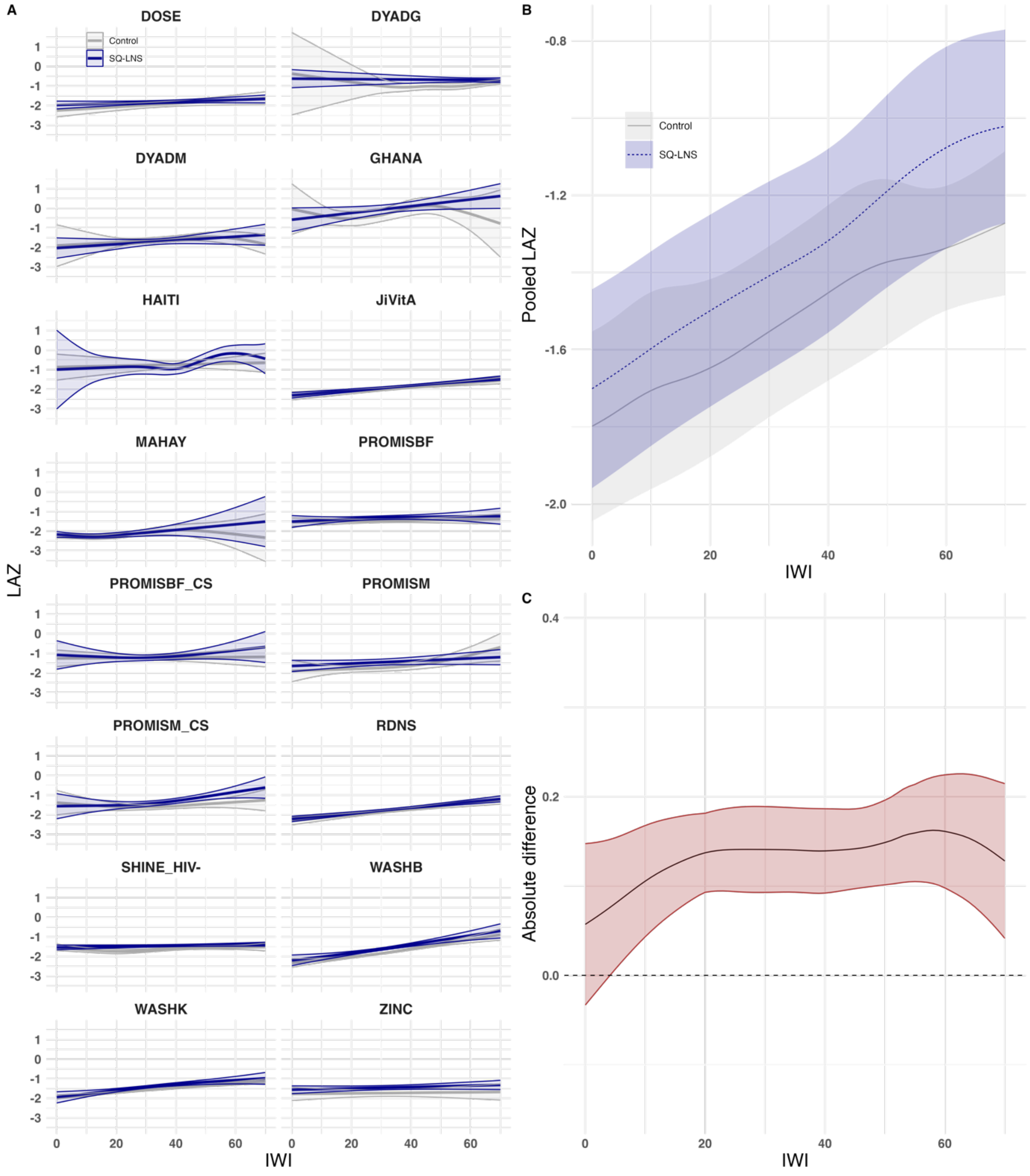

**Figure S3. Weight-for-length Z-score by international wealth index and intervention group. A.** Study-specific relationships between the International Wealth Index (IWI) and Weight-for-length Z-scores (WLZ) by intervention group. **B.** Pooled estimates from a two-stage random effects meta-analysis of WLZ by IWI and intervention group across all studies in panel A. **C.** Pooled difference in WLZ between groups over levels of IWI. Intervention effects conditional on IWI were estimated by subtracting the spline fits for the treated group from the control group within each study. These effects were then pooled using pointwise random-effects meta-analysis with Restricted Maximum Likelihood. In all panels, shaded bands represent pointwise 95% confidence intervals. Intervention excluded maternal supplementation.

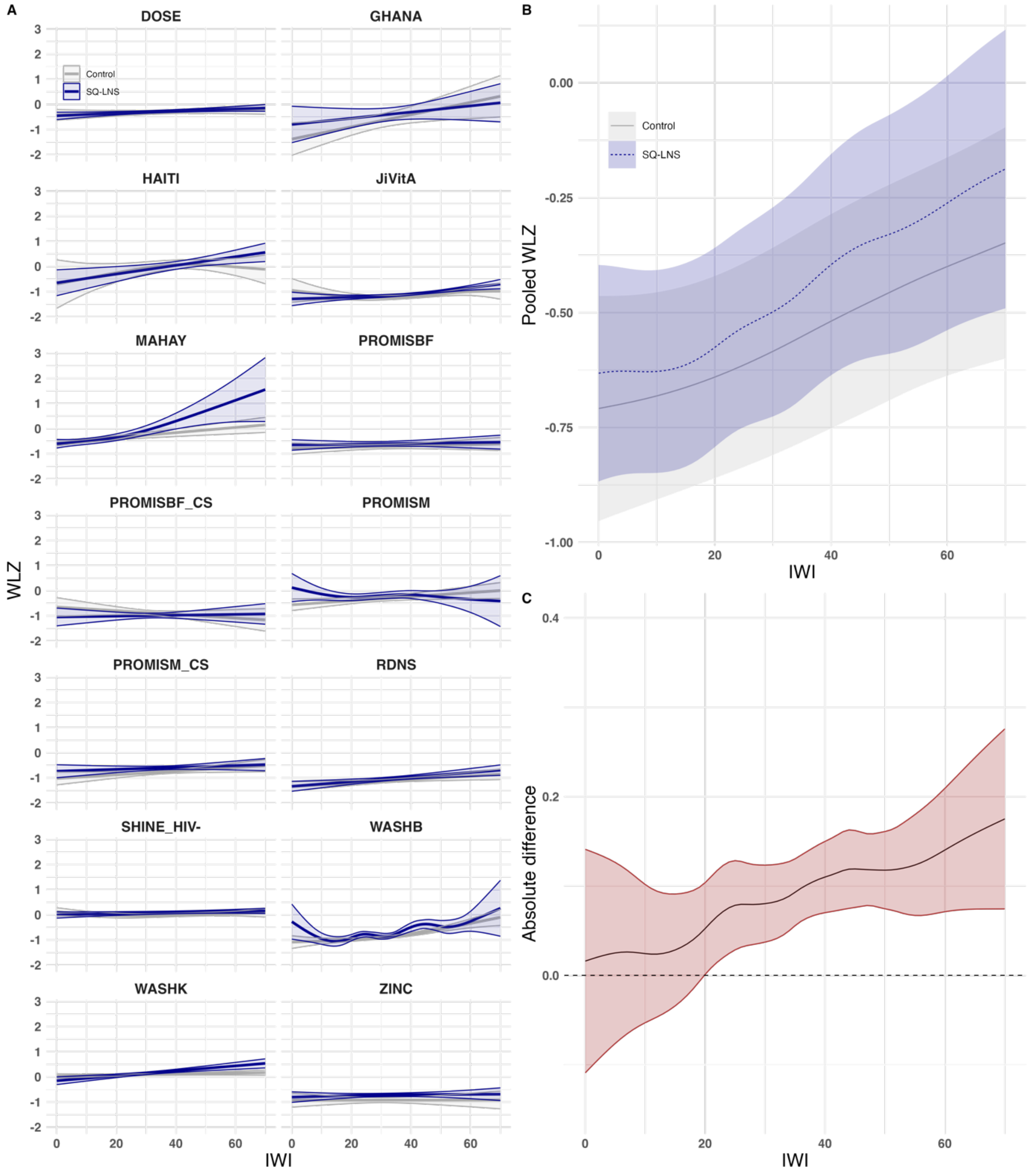

**Figure S4. Weight-for-length Z-score by international wealth index and intervention group. A.** Study-specific relationships between the International Wealth Index (IWI) and Weight-for-length Z-scores (WLZ) by intervention group. **B.** Pooled estimates from a two-stage random effects meta-analysis of WLZ by IWI and intervention group across all studies in panel A. **C.** Pooled difference in WLZ between groups over levels of IWI. Intervention effects conditional on IWI were estimated by subtracting the spline fits for the treated group from the control group within each study. These effects were then pooled using pointwise random-effects meta-analysis with Restricted Maximum Likelihood. In all panels, shaded bands represent pointwise 95% confidence intervals. Intervention included maternal supplementation.

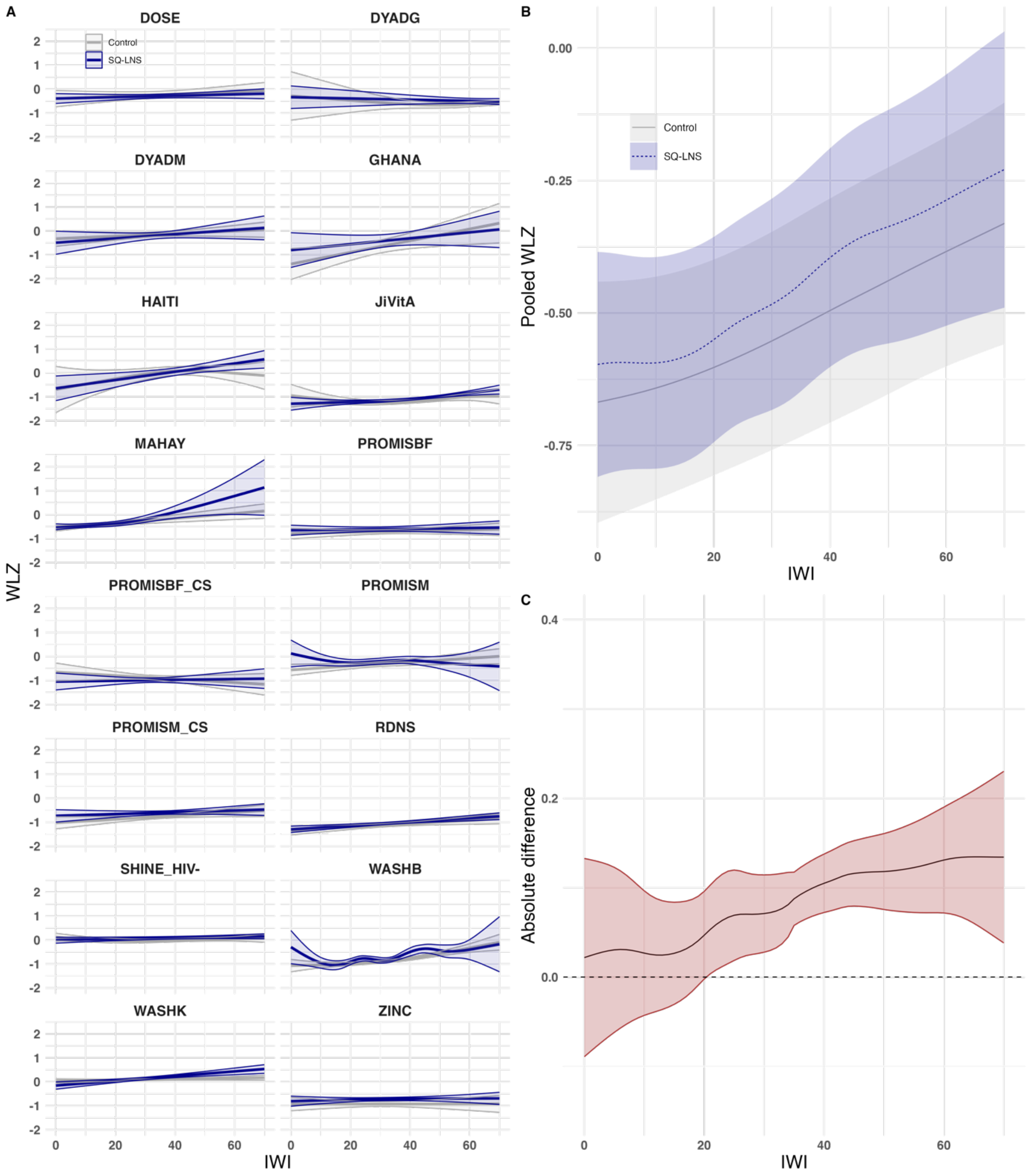

**Figure S5. Probability of stunting by international wealth index and intervention group. A.** Study-specific relationships between the International Wealth Index (IWI) and probability of stunting by intervention group. **B.** Pooled estimates from a two-stage random effects meta-analysis of probability of stunting by IWI and intervention group across all studies in panel A. **C.** Pooled difference in probability of stunting between groups over levels of IWI. Intervention effects conditional on IWI were estimated by subtracting the spline fits for the treated group from the control group within each study. These effects were then pooled using pointwise random-effects meta-analysis with Restricted Maximum Likelihood. In all panels, shaded bands represent pointwise 95% confidence intervals. <u>Intervention excluded maternal supplementation.</u>

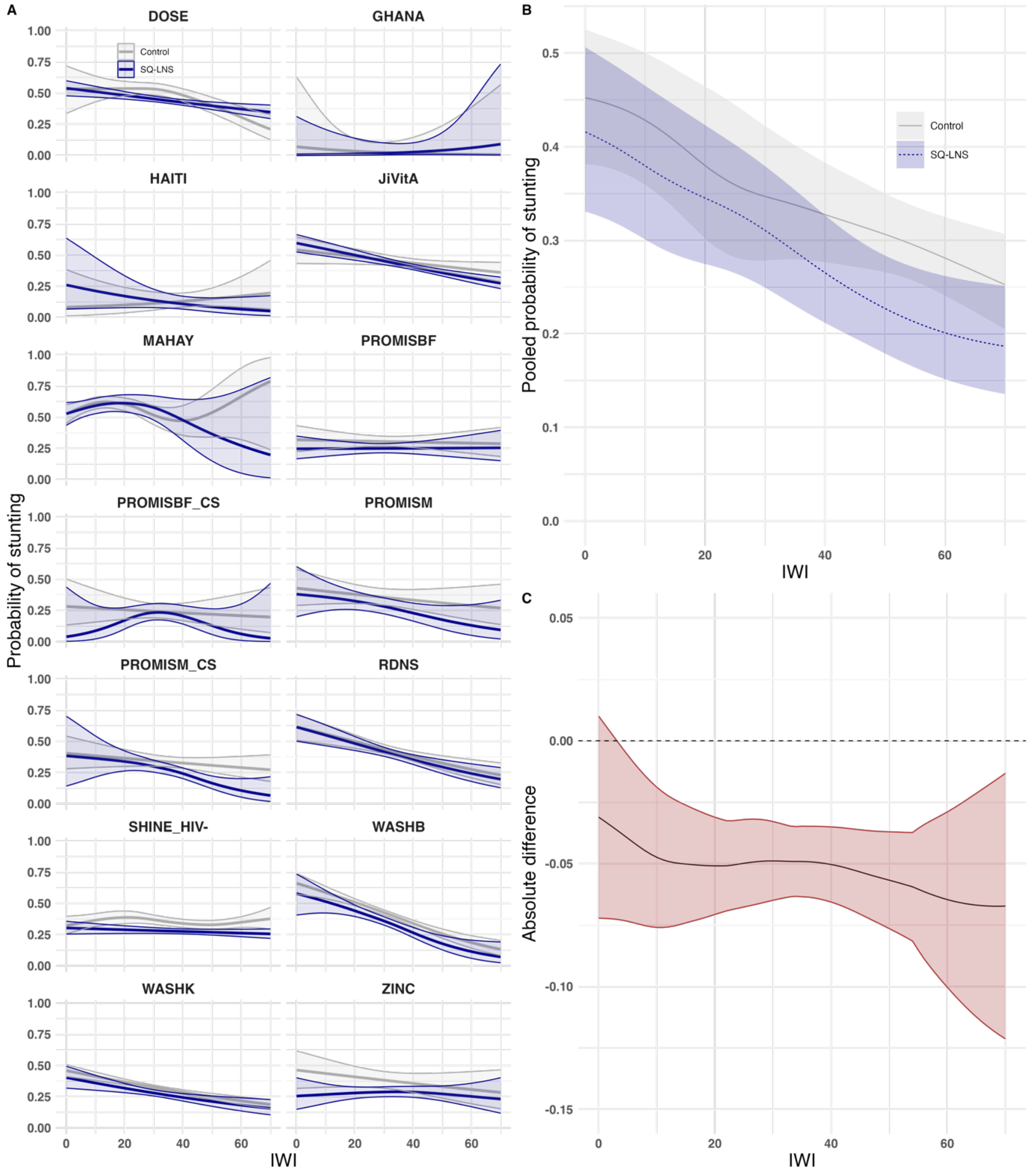

**Figure S6. Probability of stunting by international wealth index and intervention group. A.** Study-specific relationships between the International Wealth Index (IWI) and probability of stunting by intervention group. **B.** Pooled estimates from a two-stage random effects meta-analysis of probability of stunting by IWI and intervention group across all studies in panel A. **C.** Pooled difference in probability of stunting between groups over levels of IWI. Intervention effects conditional on IWI were estimated by subtracting the spline fits for the treated group from the control group within each study. These effects were then pooled using pointwise random-effects meta-analysis with Restricted Maximum Likelihood. In all panels, shaded bands represent pointwise 95% confidence intervals. <u>Intervention included maternal supplementation.</u>

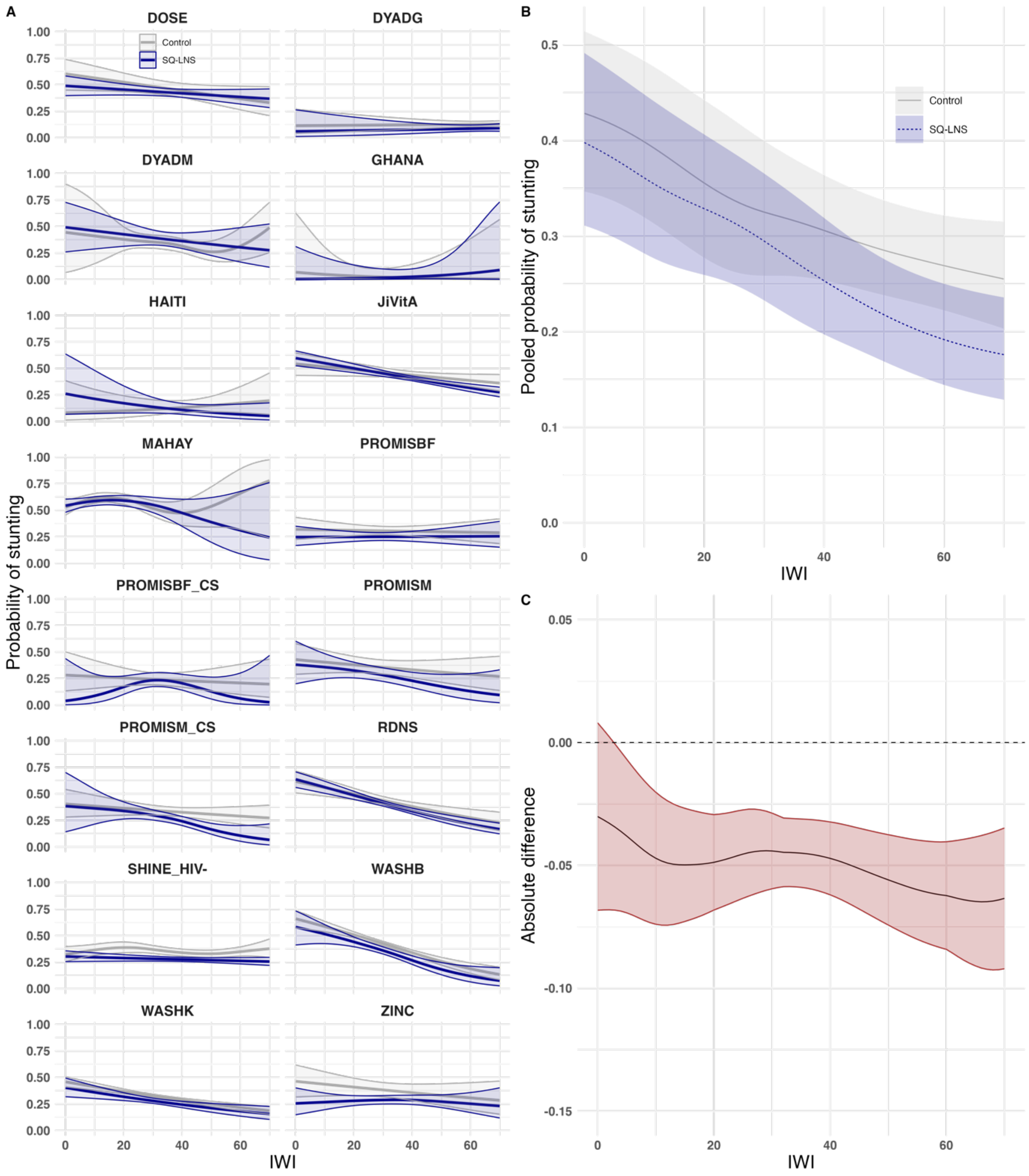

**Figure S7. Probability of wasting by international wealth index and intervention group. A.** Study-specific relationships between the International Wealth Index (IWI) and probability of wasting by intervention group. **B.** Pooled estimates from a two-stage random effects meta-analysis of probability of wasting by IWI and intervention group across all studies in panel A. **C.** Pooled difference in probability of wasting between groups over levels of IWI. Intervention effects conditional on IWI were estimated by subtracting the spline fits for the treated group from the control group within each study. These effects were then pooled using pointwise random-effects meta-analysis with Restricted Maximum Likelihood. In all panels, shaded bands represent pointwise 95% confidence intervals. Intervention excluded maternal supplementation.

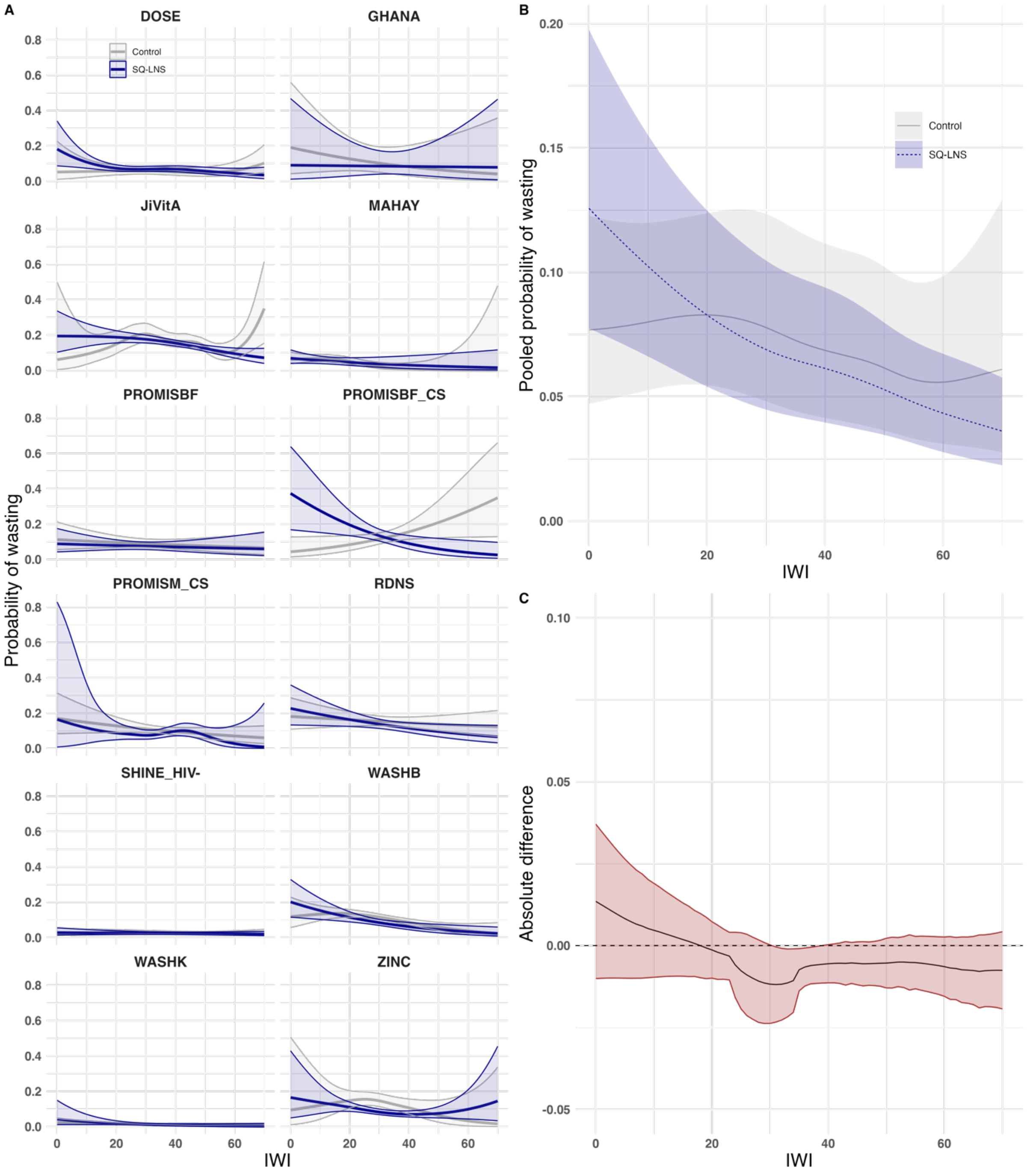

**Figure S8. Probability of wasting by international wealth index and intervention group. A.** Study-specific relationships between the International Wealth Index (IWI) and probability of wasting by intervention group. **B.** Pooled estimates from a two-stage random effects meta-analysis of probability of wasting by IWI and intervention group across all studies in panel A. **C.** Pooled difference in probability of wasting between groups over levels of IWI. Intervention effects conditional on IWI were estimated by subtracting the spline fits for the treated group from the control group within each study. These effects were then pooled using pointwise random-effects meta-analysis with Restricted Maximum Likelihood. In all panels, shaded bands represent pointwise 95% confidence intervals. Intervention included maternal supplementation.

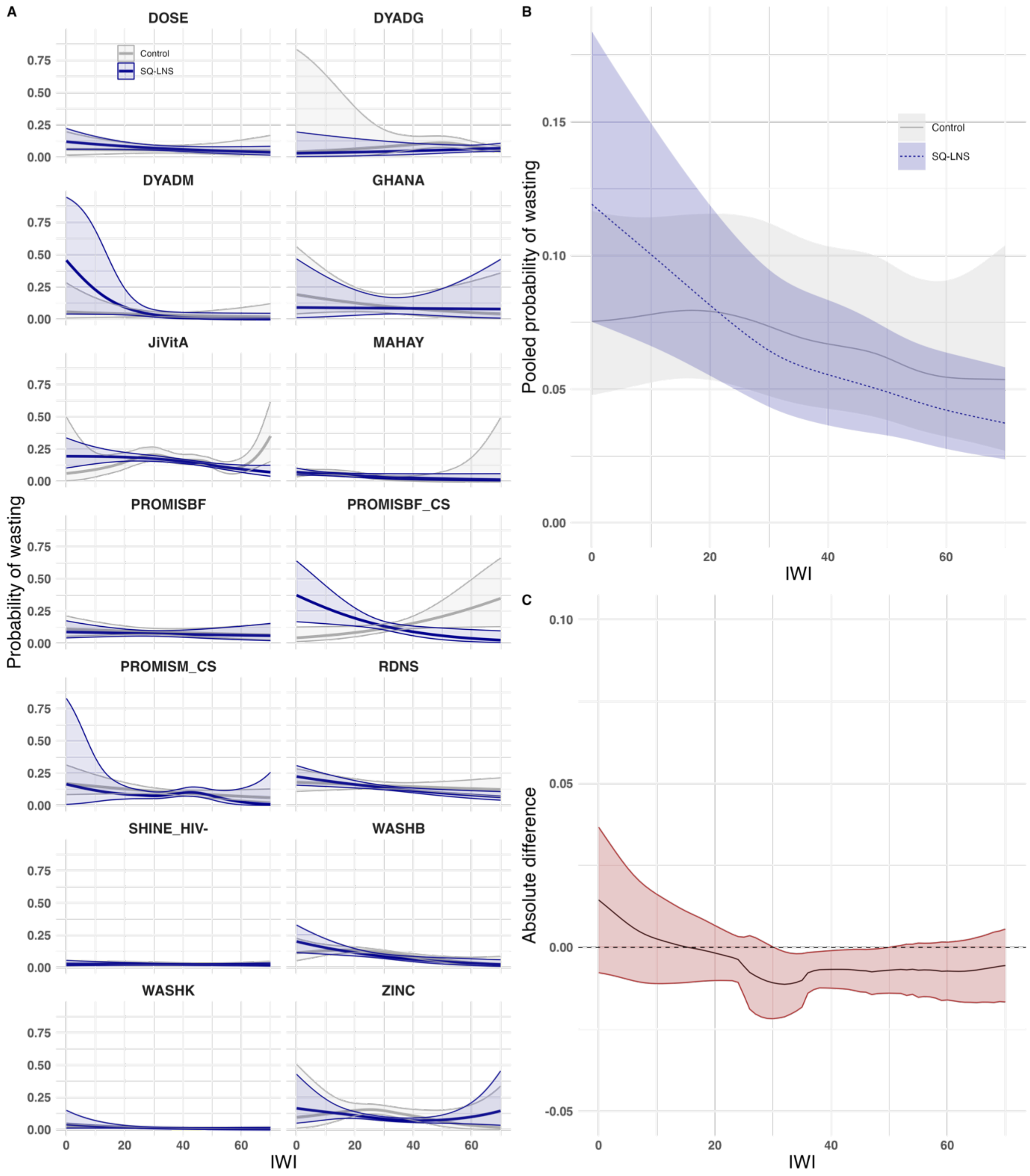

**Figure S9. Probability of severe stunting by international wealth index and intervention group. A.** Study-specific relationships between the International Wealth Index (IWI) and probability of severe stunting by intervention group. **B.** Pooled estimates from a two-stage random effects meta-analysis of probability of severe stunting by IWI and intervention group across all studies in panel A. **C.** Pooled difference in probability of severe stunting between groups over levels of IWI. Intervention effects conditional on IWI were estimated by subtracting the spline fits for the treated group from the control group within each study. These effects were then pooled using pointwise random-effects meta-analysis with Restricted Maximum Likelihood. In all panels, shaded bands represent pointwise 95% confidence intervals. Intervention excluded maternal supplementation.

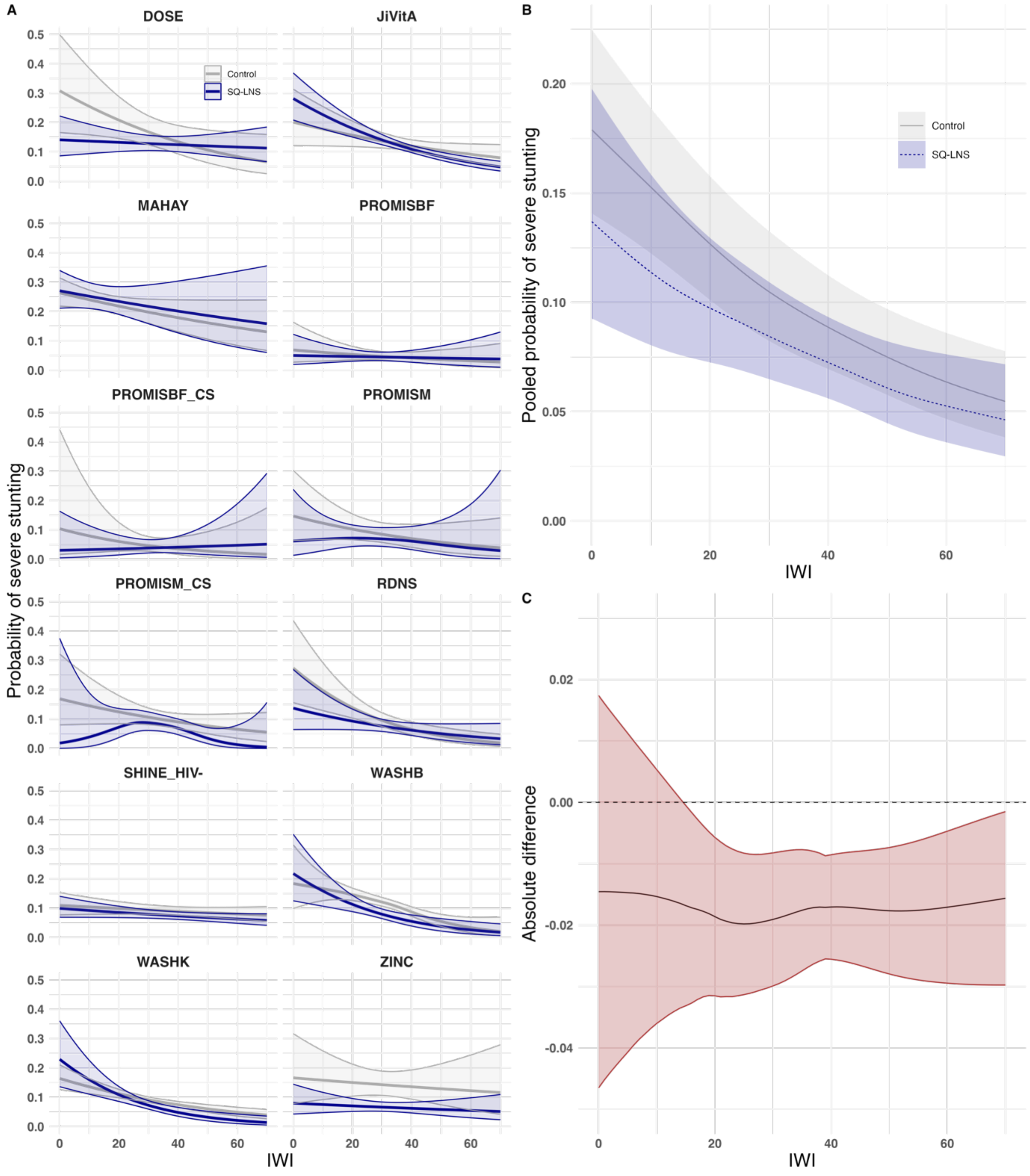

**Figure S10. Probability of severe stunting by international wealth index and intervention group. A.** Study-specific relationships between the International Wealth Index (IWI) and probability of severe stunting by intervention group. **B.** Pooled estimates from a two-stage random effects meta-analysis of probability of severe stunting by IWI and intervention group across all studies in panel A. **C.** Pooled difference in probability of severe stunting between groups over levels of IWI. Intervention effects conditional on IWI were estimated by subtracting the spline fits for the treated group from the control group within each study. These effects were then pooled using pointwise random-effects meta-analysis with Restricted Maximum Likelihood. In all panels, shaded bands represent pointwise 95% confidence intervals. Intervention included maternal supplementation.

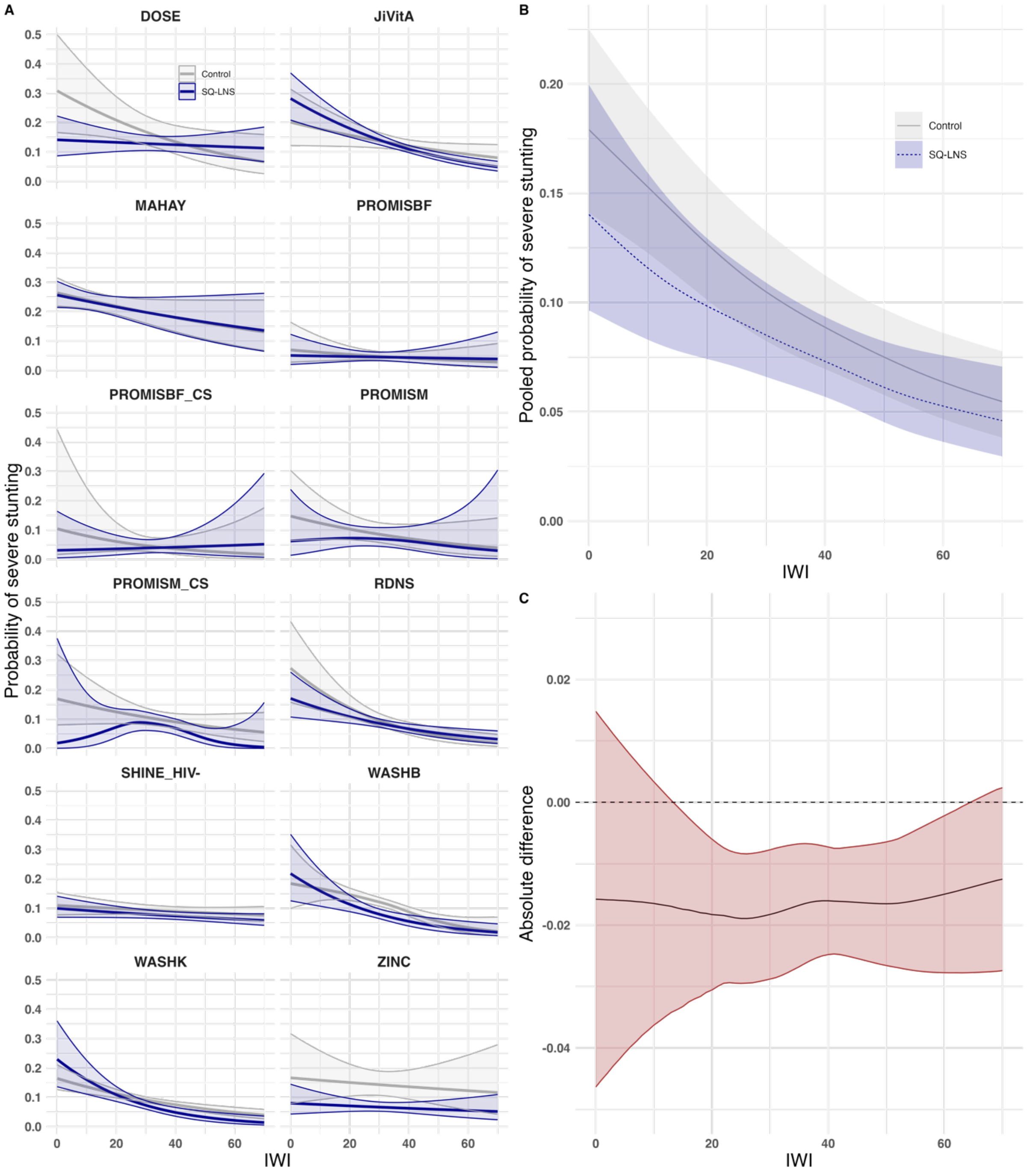

**Figure S11. Probability of severe wasting by international wealth index and intervention group. A.** Study-specific relationships between the International Wealth Index (IWI) and probability of severe wasting by intervention group. **B.** Pooled estimates from a two-stage random effects meta-analysis of probability of severe wasting by IWI and intervention group across all studies in panel A. **C.** Pooled difference in probability of severe wasting between groups over levels of IWI. Intervention effects conditional on IWI were estimated by subtracting the spline fits for the treated group from the control group within each study. These effects were then pooled using pointwise random-effects meta-analysis with Restricted Maximum Likelihood. In all panels, shaded bands represent pointwise 95% confidence intervals. Intervention excluded maternal supplementation. Number of events/ children: DOSE (11/ 926), GHANA (5/ 154), HAITI (0/ 297), JiVitA (88/ 3,964), MAHAY (33/ 2,513), PROMISBF (14/ 1,749), PROMISBF_CS (12/ 852), PROMISM (1/ 1,009), PROMISM_CS (31/ 1,840), RDNS (23/ 1,572), SHINE_HIV- (16/ 3,637), WASHB (47/ 4,506), WASHK (14/ 6,559) and ZINC (35/ 2,572).

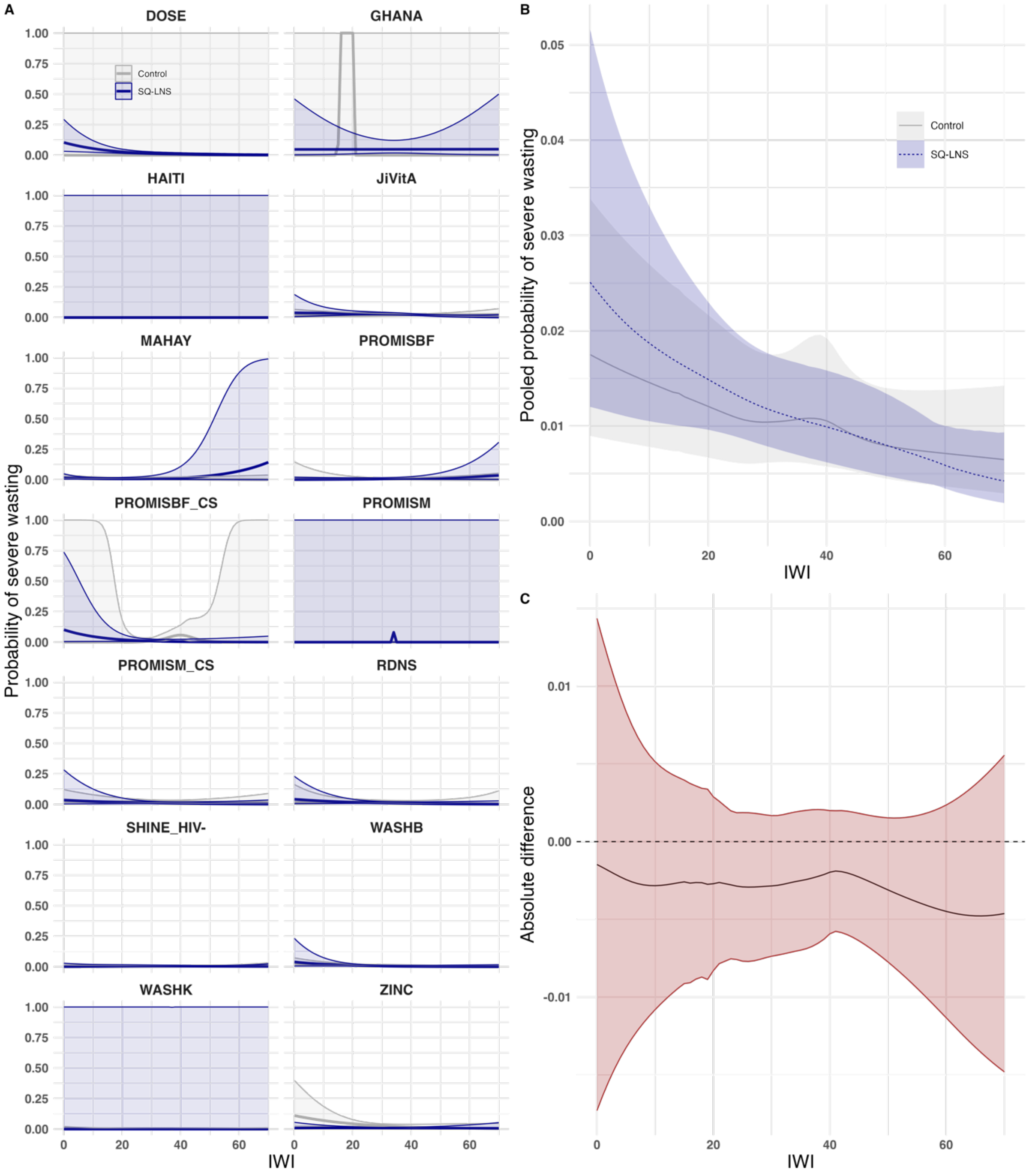

**Figure S12. Probability of severe wasting by international wealth index and intervention group. A.** Study-specific relationships between the International Wealth Index (IWI) and probability of severe wasting by intervention group. **B.** Pooled estimates from a two-stage random effects meta-analysis of probability of severe wasting by IWI and intervention group across all studies in panel A. **C.** Pooled difference in probability of severe wasting between groups over levels of IWI. Intervention effects conditional on IWI were estimated by subtracting the spline fits for the treated group from the control group within each study. These effects were then pooled using pointwise random-effects meta-analysis with Restricted Maximum Likelihood. In all panels, shaded bands represent pointwise 95% confidence intervals. <u>Intervention included maternal supplementation.</u> Number of events/ children: DOSE (11/ 926), DYADG (10/ 1,027), DYADM (6/ 650), GHANA (5/ 154), HAITI (0/ 297), JiVitA (88/ 3,964), MAHAY (38/ 3,344), PROMISBF (14/ 1,749), PROMISBF_CS (12/ 852), PROMISM (1/1,009), PROMISM_CS (31/ 1,840), RDNS (34/ 2,442), SHINE_HIV- (16/ 3,637), WASHB (47/4,506), WASHK (14/ 6,559), ZINC (35/ 2,527).

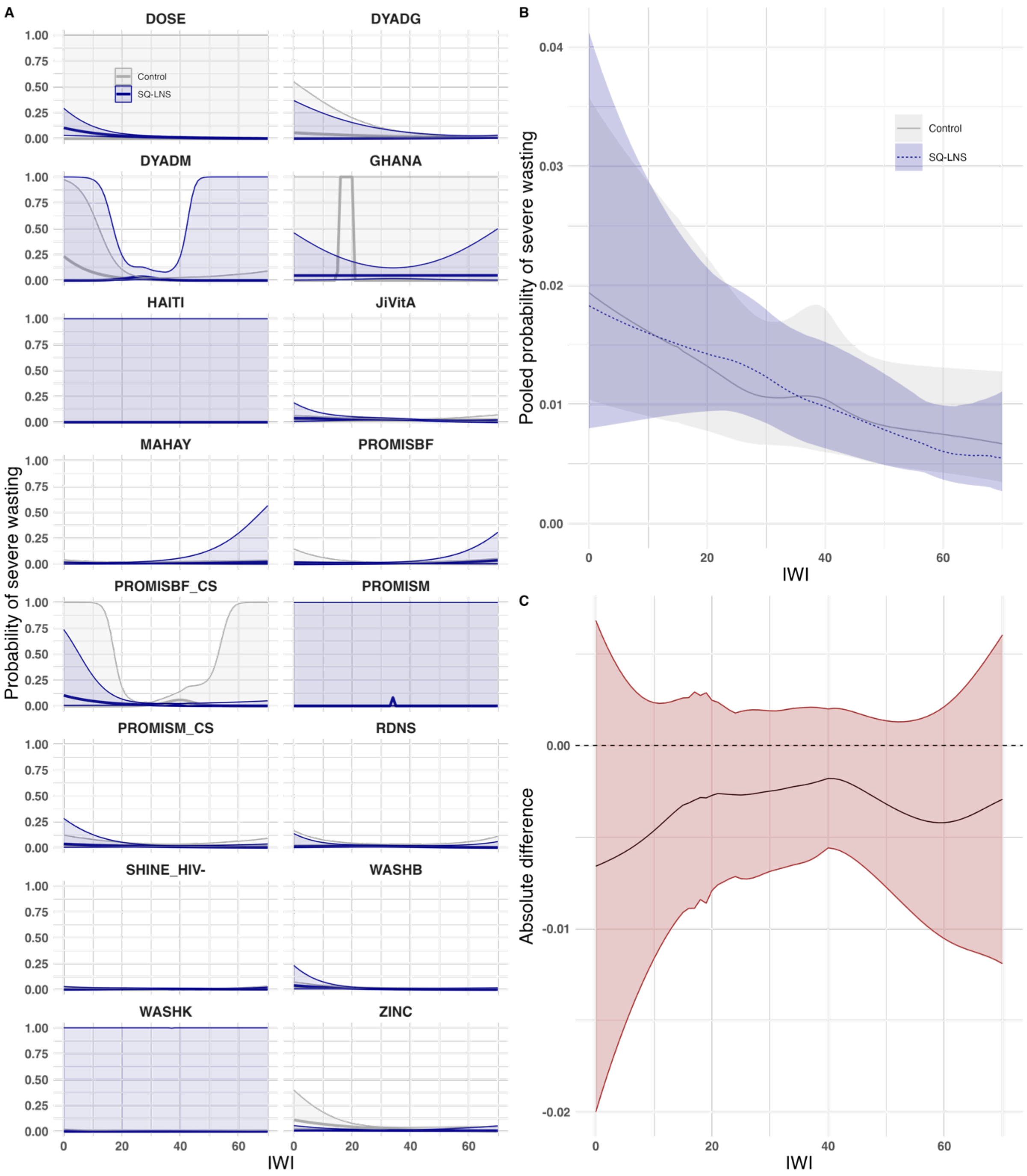

**Figure S13. Language score by international wealth index and intervention group. A.** Study-specific relationships between the International Wealth Index (IWI) and language score by intervention group. **B.** Pooled estimates from a two-stage random effects meta-analysis of language score by IWI and intervention group across all studies in panel A. **C.** Pooled difference in language score between groups over levels of IWI. Intervention effects conditional on IWI were estimated by subtracting the spline fits for the treated group from the control group within each study. These effects were then pooled using pointwise random-effects meta-analysis with Restricted Maximum Likelihood. In all panels, shaded bands represent pointwise 95% confidence intervals. <u>Intervention excluded maternal supplementation.</u>

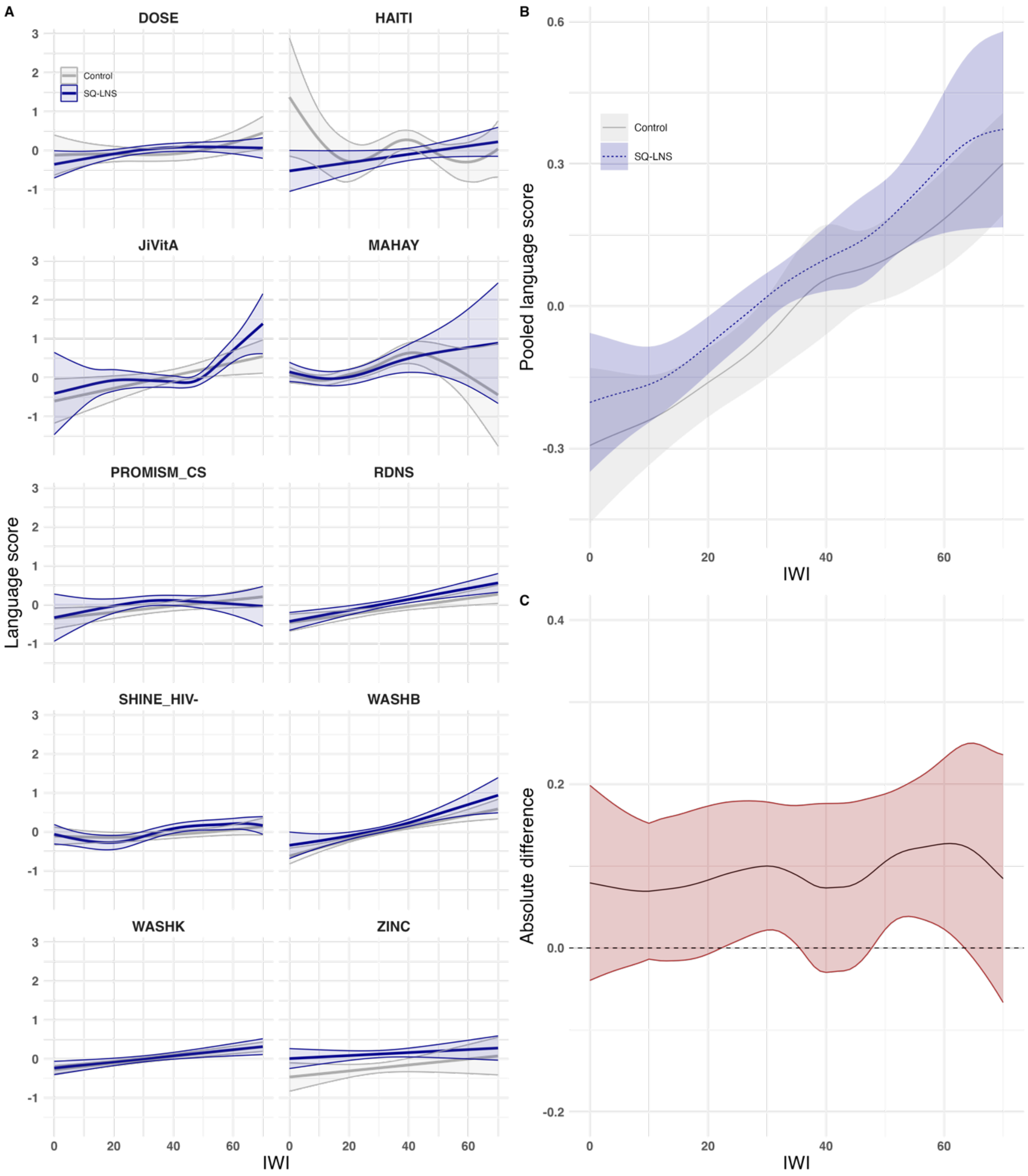

**Figure S14. Language score by international wealth index and intervention group. A.** Study-specific relationships between the International Wealth Index (IWI) and language score by intervention group. **B.** Pooled estimates from a two-stage random effects meta-analysis of language score by IWI and intervention group across all studies in panel A. **C.** Pooled difference in language score between groups over levels of IWI. Intervention effects conditional on IWI were estimated by subtracting the spline fits for the treated group from the control group within each study. These effects were then pooled using pointwise random-effects meta-analysis with Restricted Maximum Likelihood. In all panels, shaded bands represent pointwise 95% confidence intervals. <u>Intervention included maternal supplementation.</u>

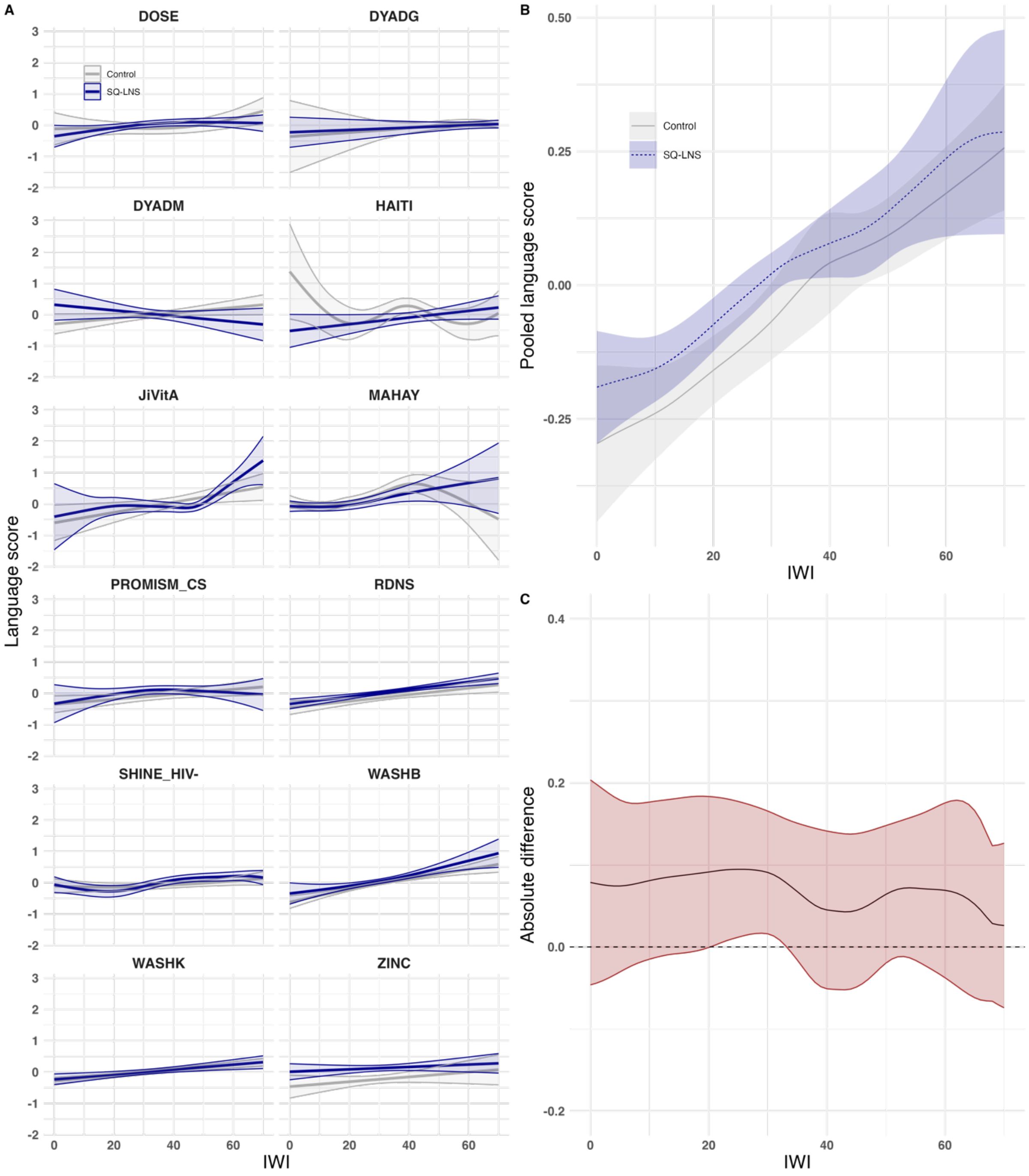

**Figure S15. Gross motor score by international wealth index and intervention group. A.** Study-specific relationships between the International Wealth Index (IWI) and gross motor score by intervention group. **B.** Pooled estimates from a two-stage random effects meta-analysis of gross motor score by IWI and intervention group across all studies in panel A. **C.** Pooled difference in gross motor score between groups over levels of IWI. Intervention effects conditional on IWI were estimated by subtracting the spline fits for the treated group from the control group within each study. These effects were then pooled using pointwise random-effects meta-analysis with Restricted Maximum Likelihood. In all panels, shaded bands represent pointwise 95% confidence intervals. <u>Intervention excluded maternal supplementation.</u>

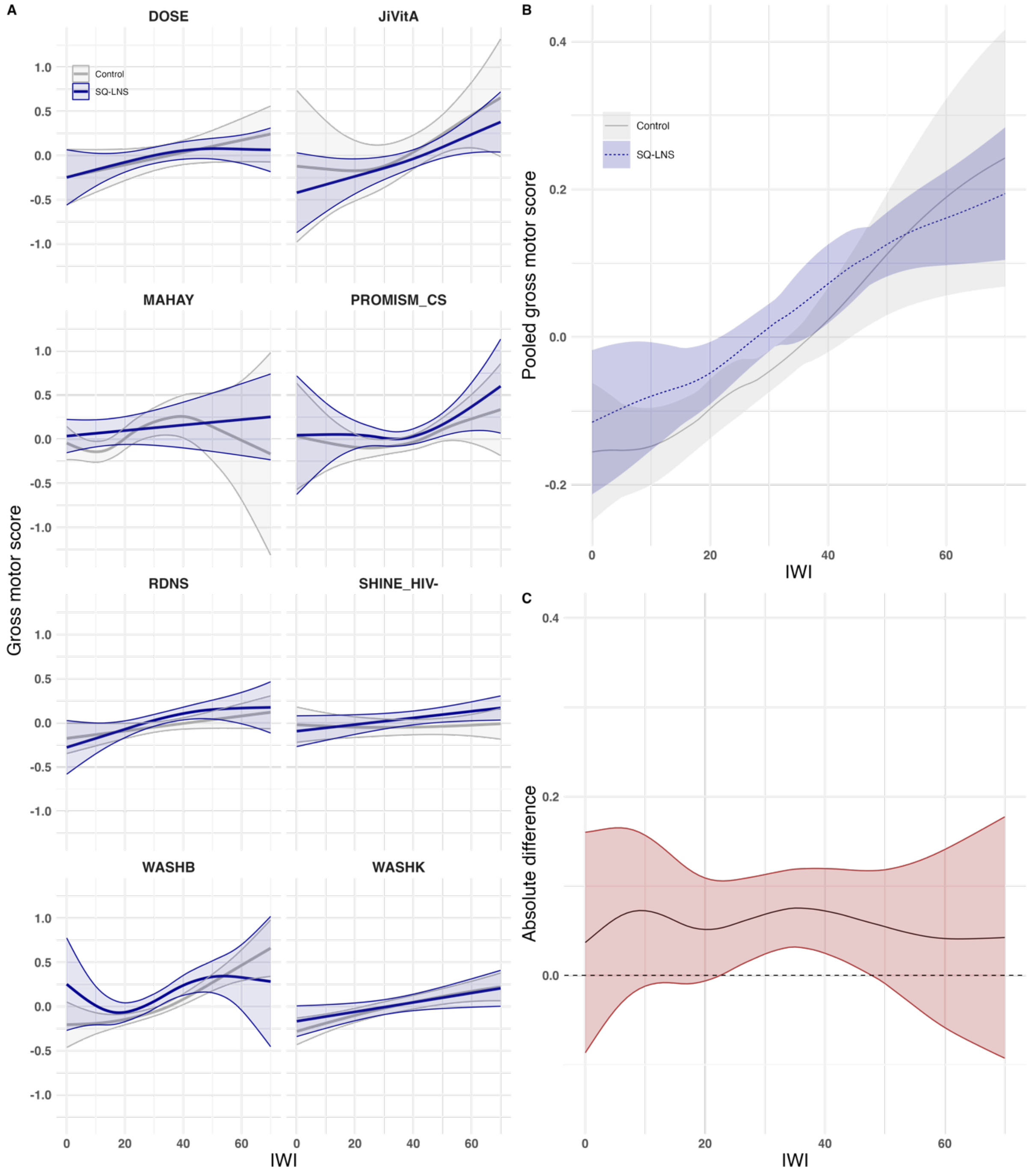

**Figure S16. Gross motor score by international wealth index and intervention group. A.** Study-specific relationships between the International Wealth Index (IWI) and gross motor score by intervention group. **B.** Pooled estimates from a two-stage random effects meta-analysis of gross motor score by IWI and intervention group across all studies in panel A. **C.** Pooled difference in gross motor score between groups over levels of IWI. Intervention effects conditional on IWI were estimated by subtracting the spline fits for the treated group from the control group within each study. These effects were then pooled using pointwise random-effects meta-analysis with Restricted Maximum Likelihood. In all panels, shaded bands represent pointwise 95% confidence intervals. Intervention included maternal supplementation.

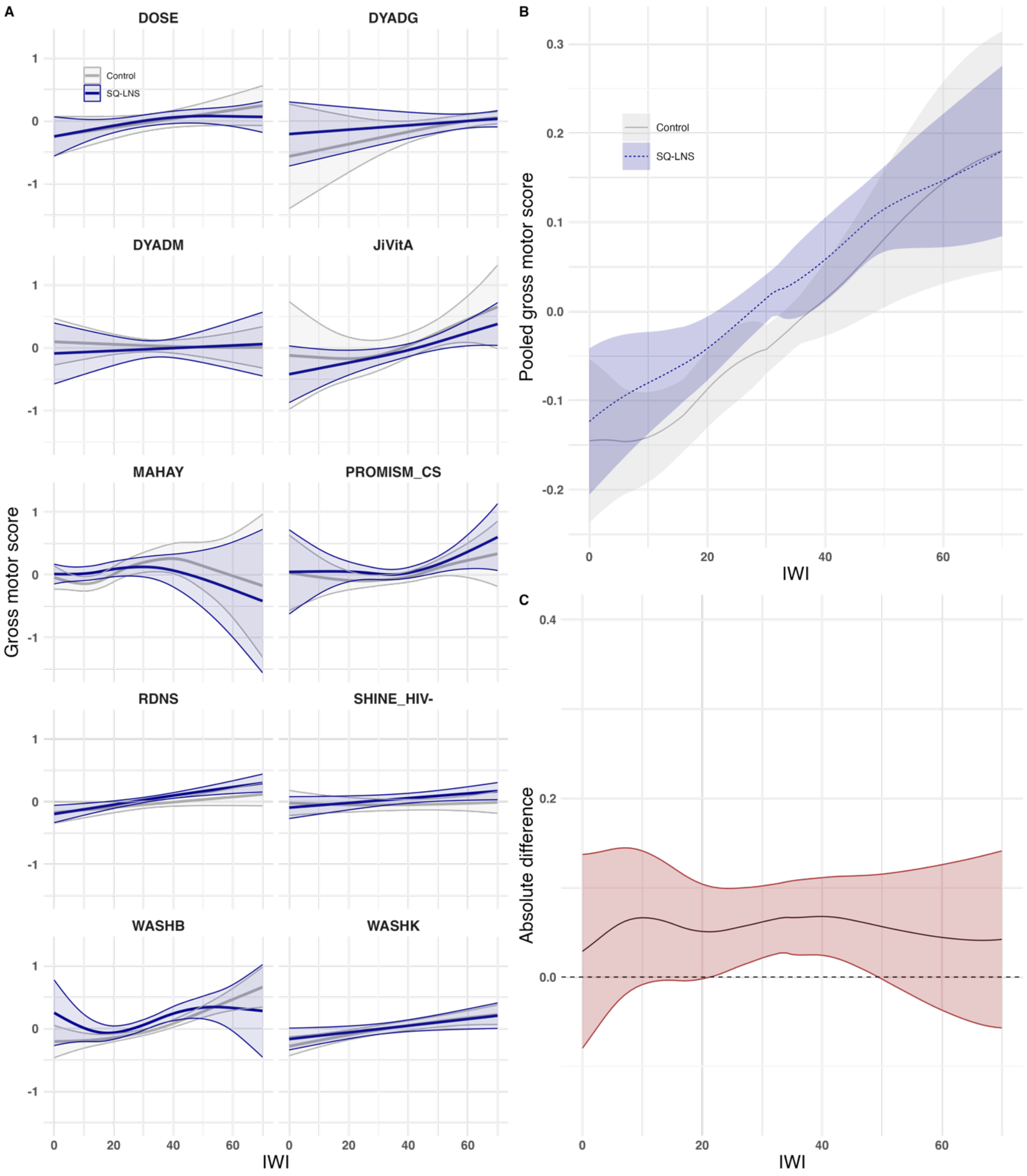

**Figure S17. Fine motor score by international wealth index and intervention group. A.** Study-specific relationships between the International Wealth Index (IWI) and fine motor score by intervention group. **B.** Pooled estimates from a two-stage random effects meta-analysis of fine motor score by IWI and intervention group across all studies in panel A. **C.** Pooled difference in fine motor score between groups over levels of IWI. Intervention effects conditional on IWI were estimated by subtracting the spline fits for the treated group from the control group within each study. These effects were then pooled using pointwise random-effects meta-analysis with Restricted Maximum Likelihood. In all panels, shaded bands represent pointwise 95% confidence intervals. Intervention excluded maternal supplementation.

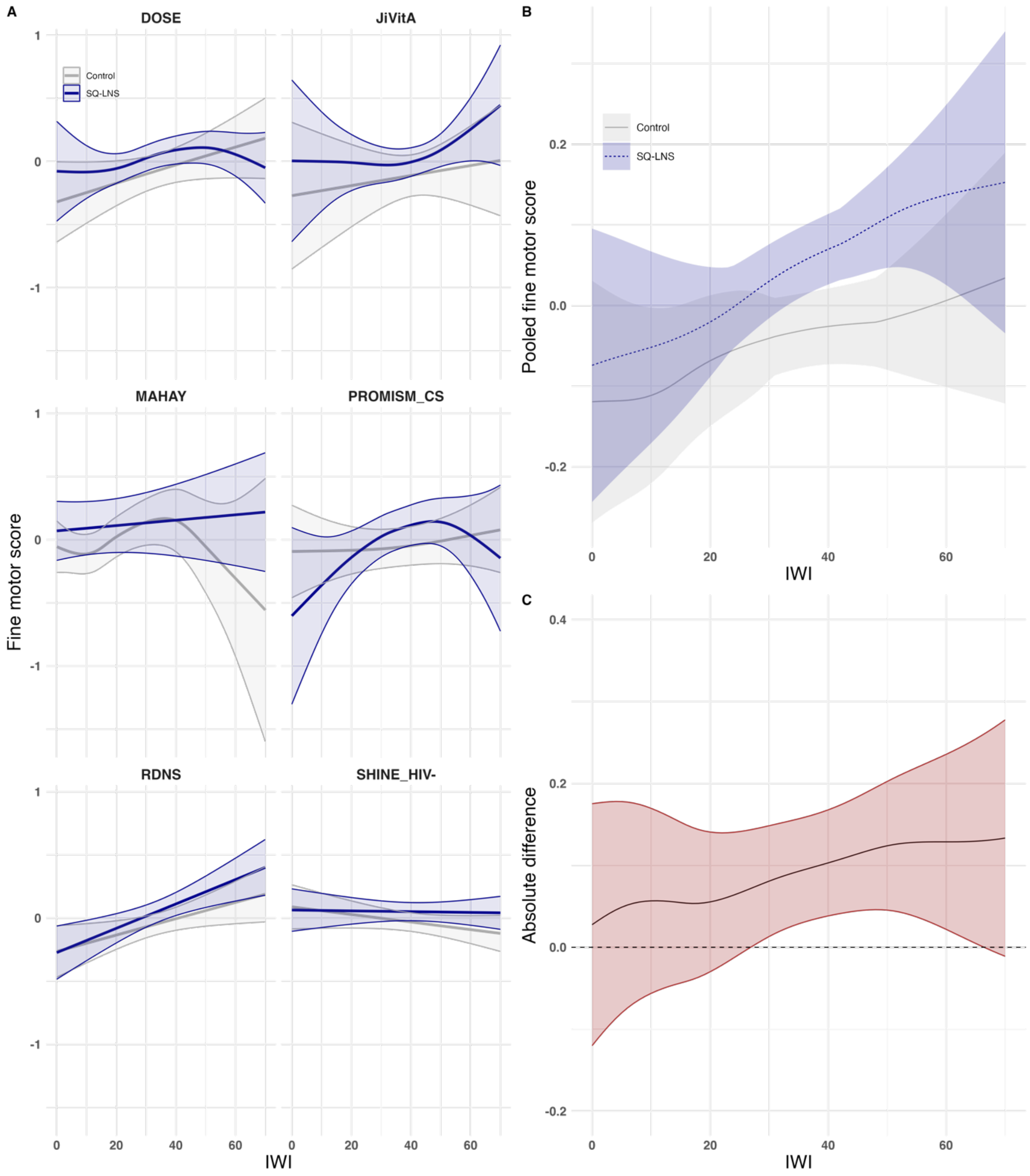

**Figure S18. Fine motor score by international wealth index and intervention group. A.** Study-specific relationships between the International Wealth Index (IWI) and fine motor score by intervention group. **B.** Pooled estimates from a two-stage random effects meta-analysis of fine motor score by IWI and intervention group across all studies in panel A. **C.** Pooled difference in fine motor score between groups over levels of IWI. Intervention effects conditional on IWI were estimated by subtracting the spline fits for the treated group from the control group within each study. These effects were then pooled using pointwise random-effects meta-analysis with Restricted Maximum Likelihood. In all panels, shaded bands represent pointwise 95% confidence intervals. <u>Intervention included maternal supplementation</u>.

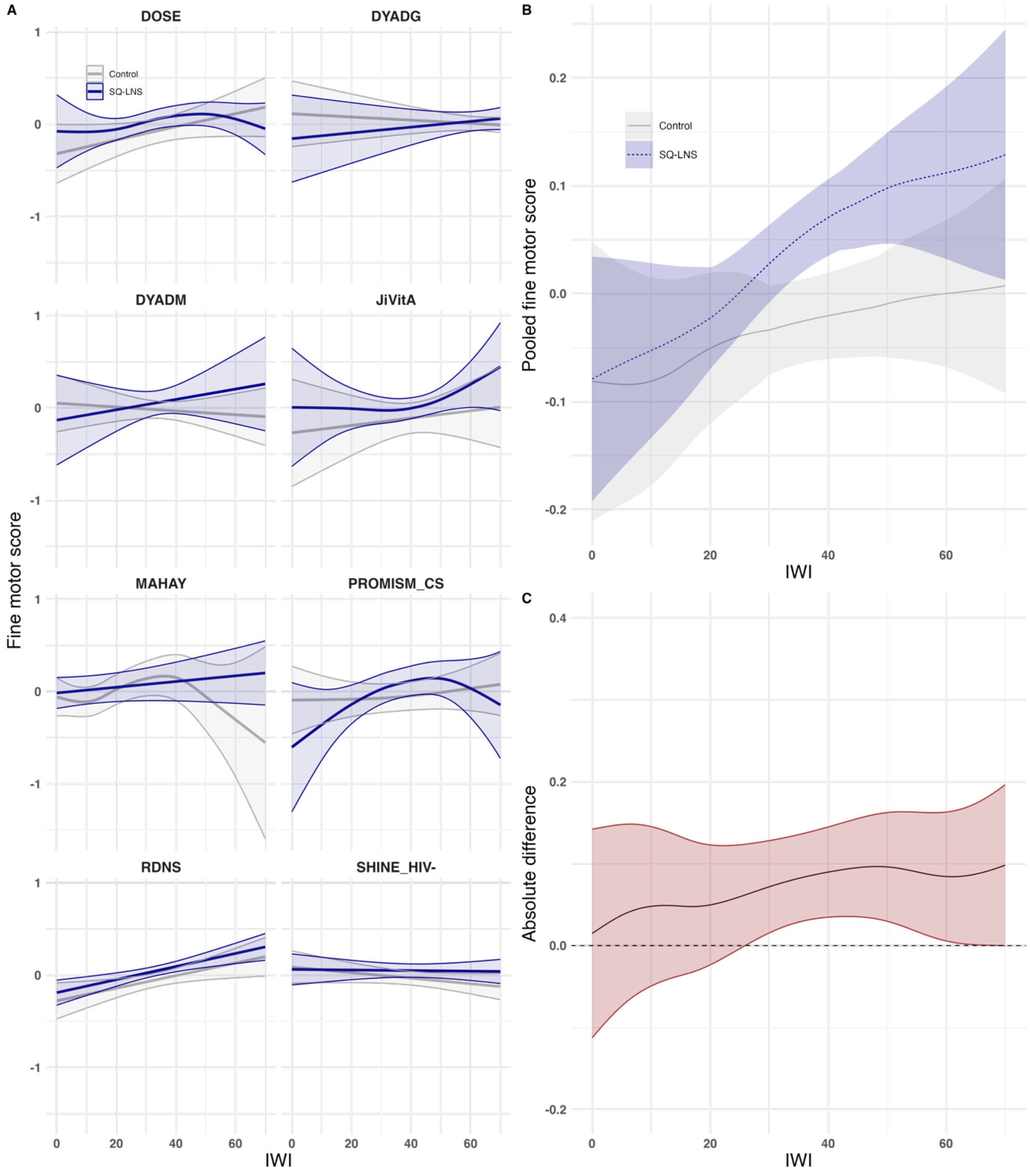

**Figure S19. Executive function score by international wealth index and intervention group. A.** Study-specific relationships between the International Wealth Index (IWI) and executive function score by intervention group. **B.** Pooled estimates from a two-stage random effects meta-analysis of executive function score by IWI and intervention group across all studies in panel A. **C.** Pooled difference in executive function score between groups over levels of IWI. Intervention effects conditional on IWI were estimated by subtracting the spline fits for the treated group from the control group within each study. These effects were then pooled using pointwise random-effects meta-analysis with Restricted Maximum Likelihood. In all panels, shaded bands represent pointwise 95% confidence intervals. Intervention excluded maternal supplementation.

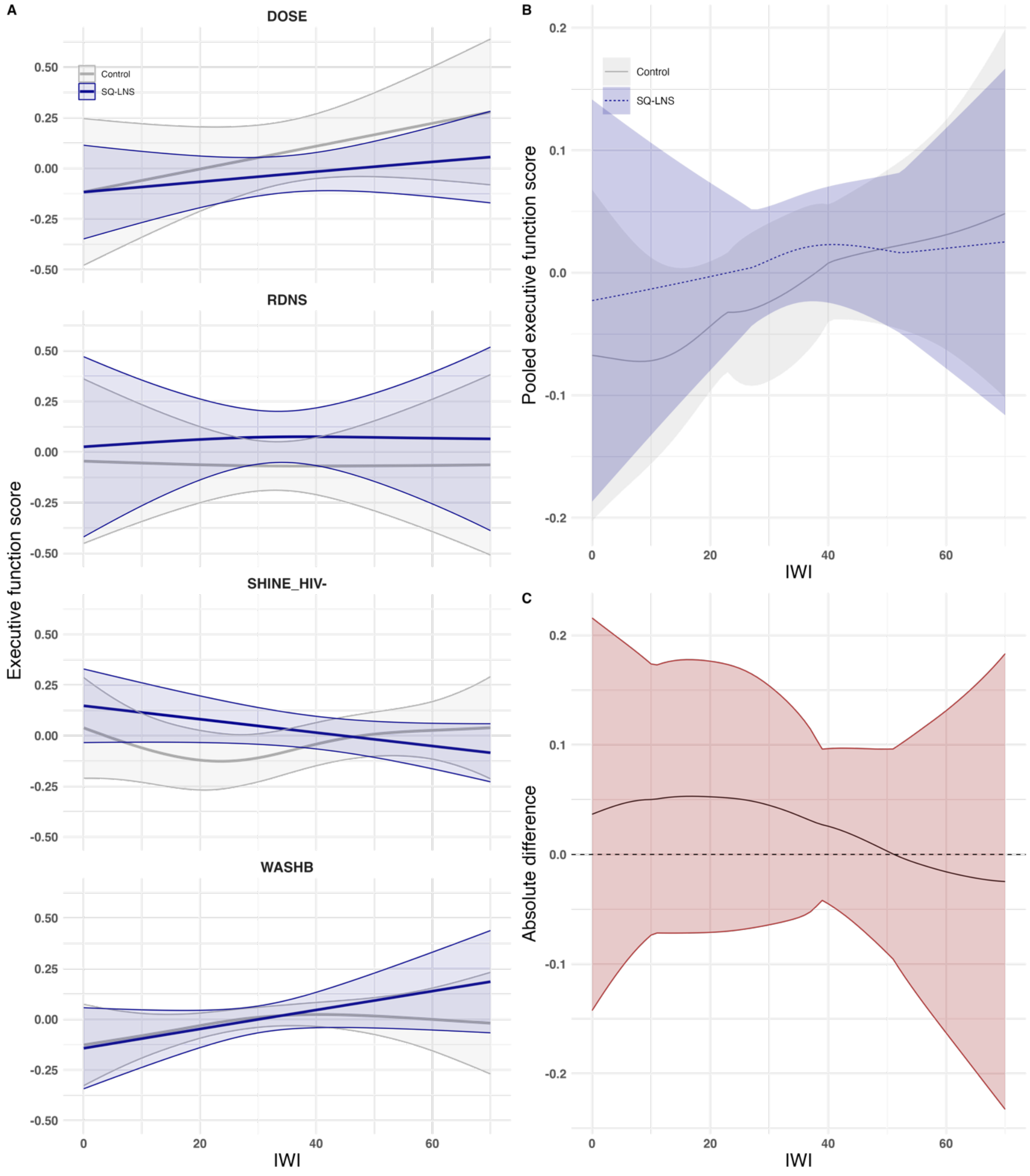

**Figure S20. Executive function score by international wealth index and intervention group. A.** Study-specific relationships between the International Wealth Index (IWI) and executive function score by intervention group. **B.** Pooled estimates from a two-stage random effects meta-analysis of executive function score by IWI and intervention group across all studies in panel A. **C.** Pooled difference in executive function score between groups over levels of IWI. Intervention effects conditional on IWI were estimated by subtracting the spline fits for the treated group from the control group within each study. These effects were then pooled using pointwise random-effects meta-analysis with Restricted Maximum Likelihood. In all panels, shaded bands represent pointwise 95% confidence intervals. Intervention included maternal supplementation.

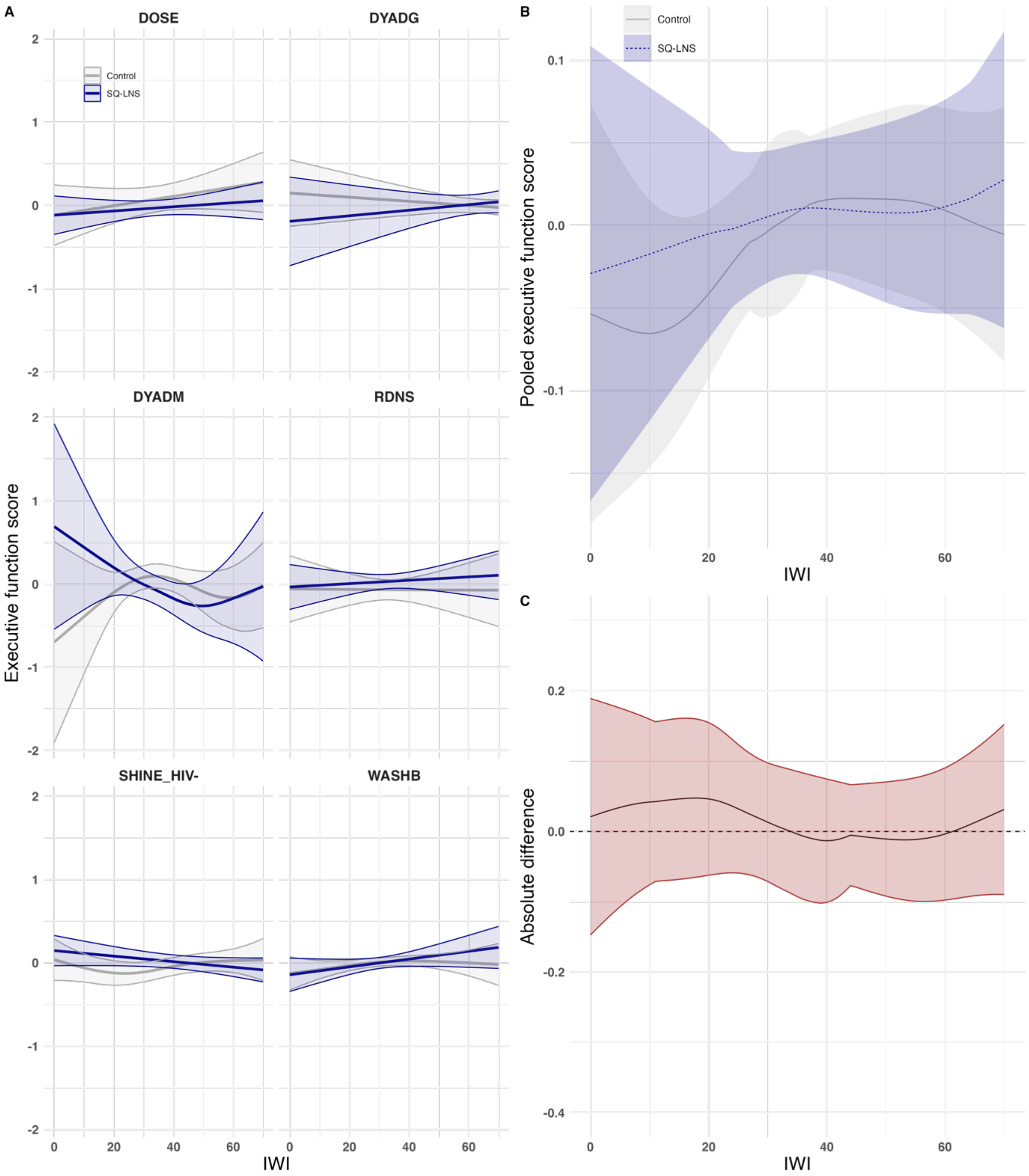

**Figure S21. Socioemotional score by international wealth index and intervention group. A.** Study-specific relationships between the International Wealth Index (IWI) and socioemotional score by intervention group. **B.** Pooled estimates from a two-stage random effects meta-analysis of socioemotional score by IWI and intervention group across all studies in panel A. **C.** Pooled difference in socioemotional score between groups over levels of IWI. Intervention effects conditional on IWI were estimated by subtracting the spline fits for the treated group from the control group within each study. These effects were then pooled using pointwise random-effects meta-analysis with Restricted Maximum Likelihood. In all panels, shaded bands represent pointwise 95% confidence intervals. Intervention excluded maternal supplementation.

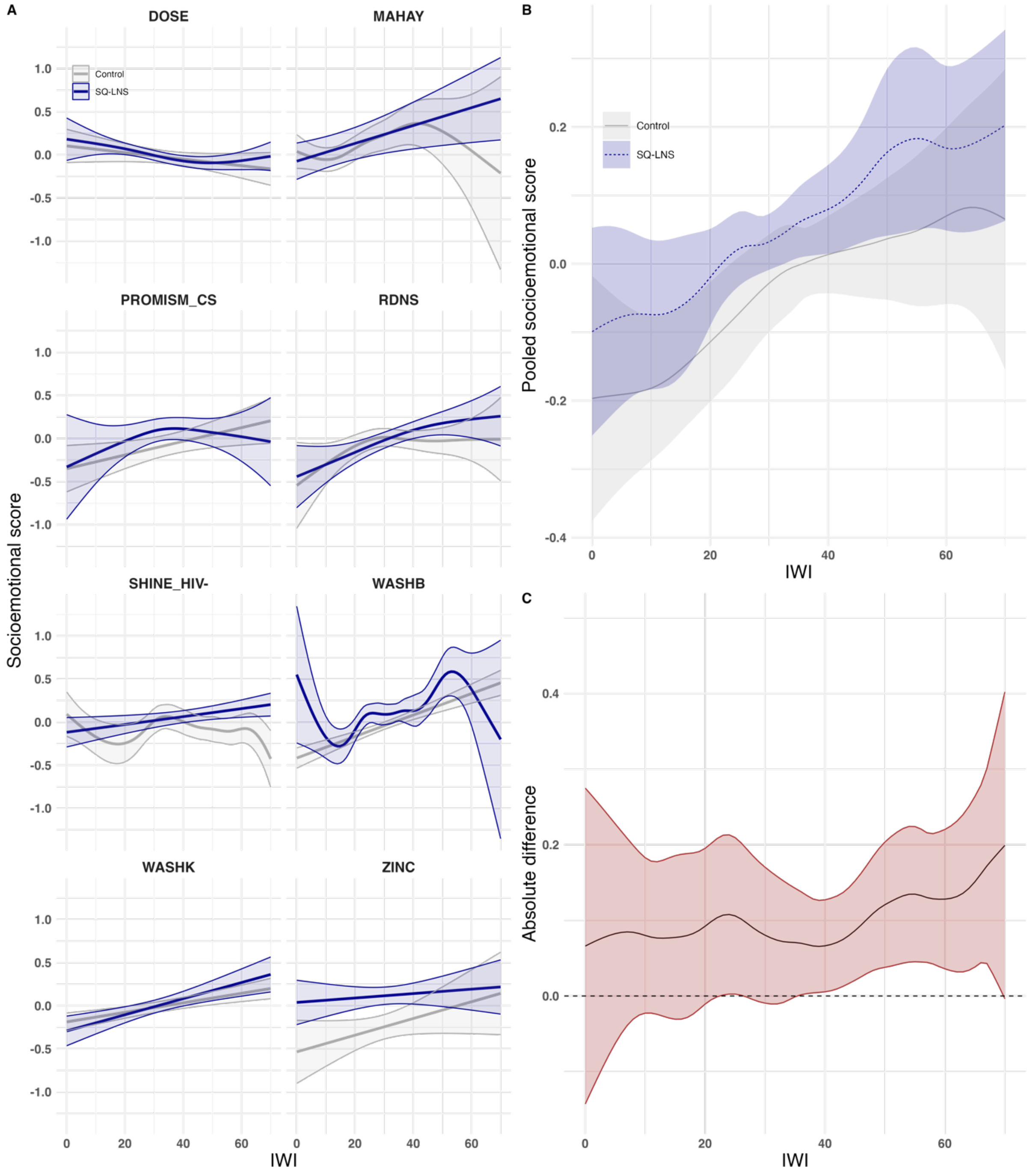

**Figure S22. Socioemotional score by international wealth index and intervention group. A.** Study-specific relationships between the International Wealth Index (IWI) and socioemotional score by intervention group. **B.** Pooled estimates from a two-stage random effects meta-analysis of socioemotional score by IWI and intervention group across all studies in panel A. **C.** Pooled difference in socioemotional score between groups over levels of IWI. Intervention effects conditional on IWI were estimated by subtracting the spline fits for the treated group from the control group within each study. These effects were then pooled using pointwise random-effects meta-analysis with Restricted Maximum Likelihood. In all panels, shaded bands represent pointwise 95% confidence intervals. Intervention included maternal supplementation.

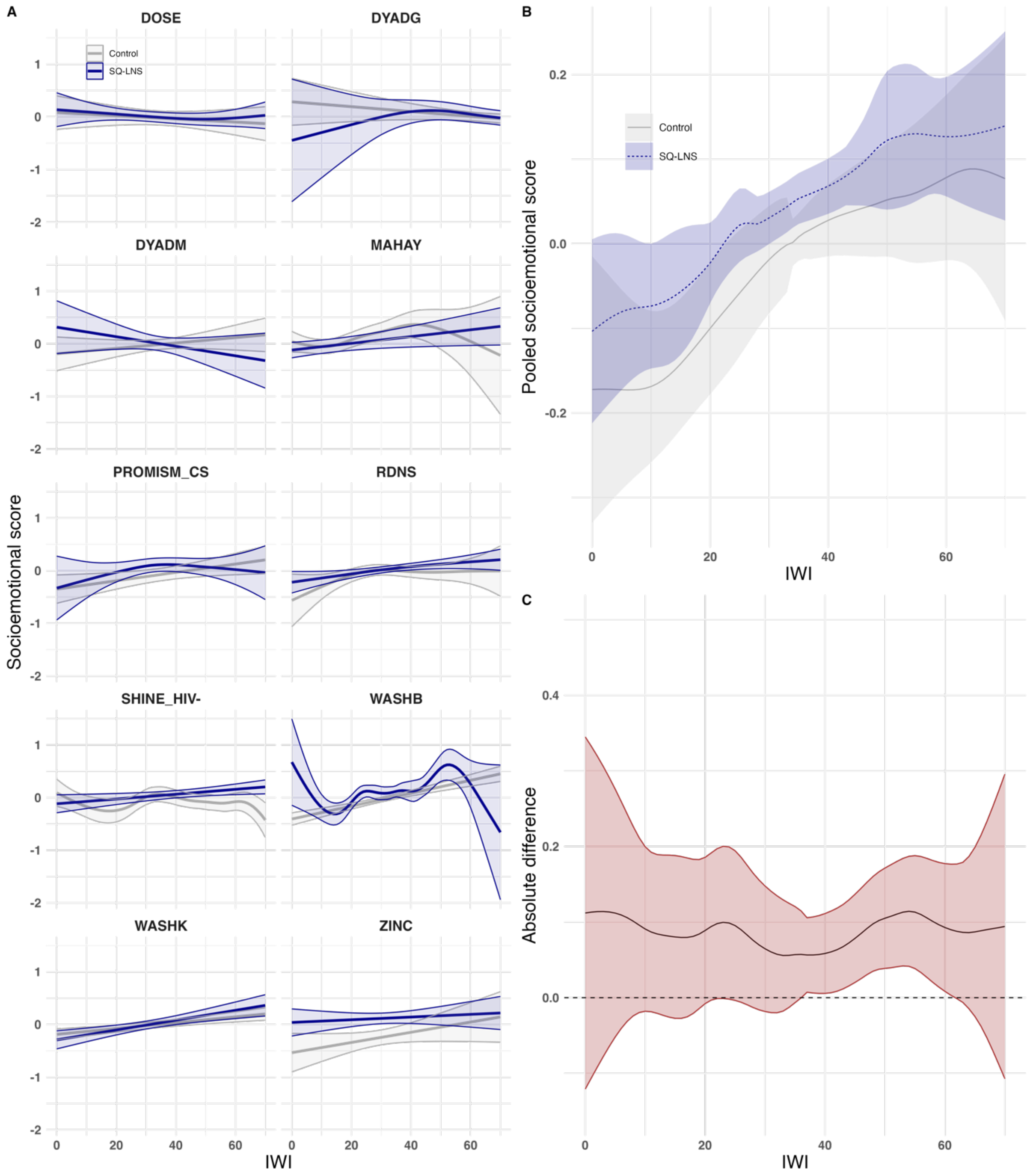

**Figure S23. Hemoglobin concentration by international wealth index and intervention group. A.** Study-specific relationships between the International Wealth Index (IWI) and hemoglobin concentration by intervention group. **B.** Pooled estimates from a two-stage random effects meta-analysis of hemoglobin concentration by IWI and intervention group across all studies in panel A. **C.** Pooled difference in hemoglobin concentration between groups over levels of IWI. Intervention effects conditional on IWI were estimated by subtracting the spline fits for the treated group from the control group within each study. These effects were then pooled using pointwise random-effects meta-analysis with Restricted Maximum Likelihood. In all panels, shaded bands represent pointwise 95% confidence intervals. Intervention excluded maternal supplementation.

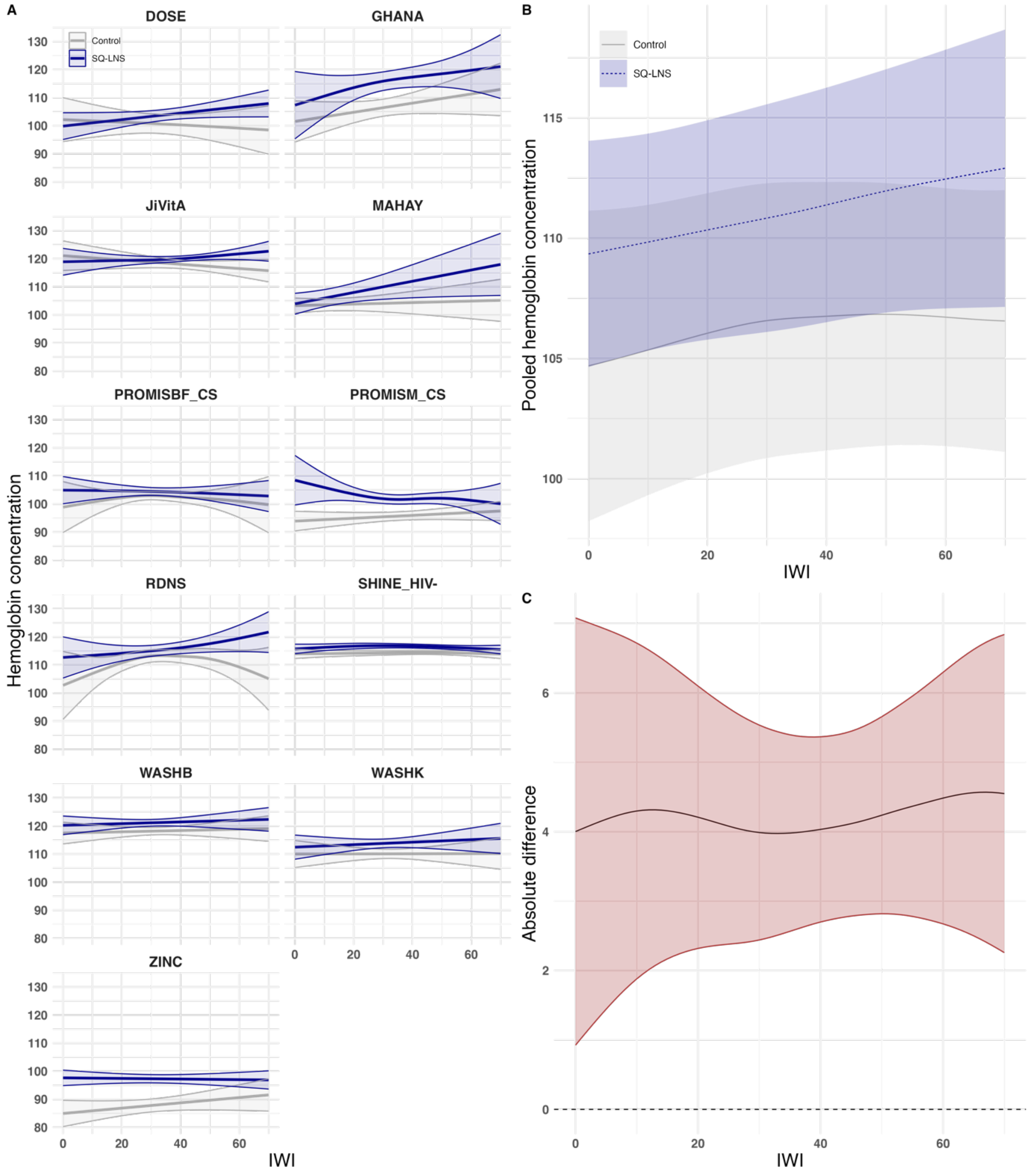

**Figure S24. Hemoglobin concentration by international wealth index and intervention group. A.** Study-specific relationships between the International Wealth Index (IWI) and hemoglobin concentration by intervention group. **B.** Pooled estimates from a two-stage random effects meta-analysis of hemoglobin concentration by IWI and intervention group across all studies in panel A. **C.** Pooled difference in hemoglobin concentration between groups over levels of IWI. Intervention effects conditional on IWI were estimated by subtracting the spline fits for the treated group from the control group within each study. These effects were then pooled using pointwise random-effects meta-analysis with Restricted Maximum Likelihood. In all panels, shaded bands represent pointwise 95% confidence intervals. Intervention included maternal supplementation.

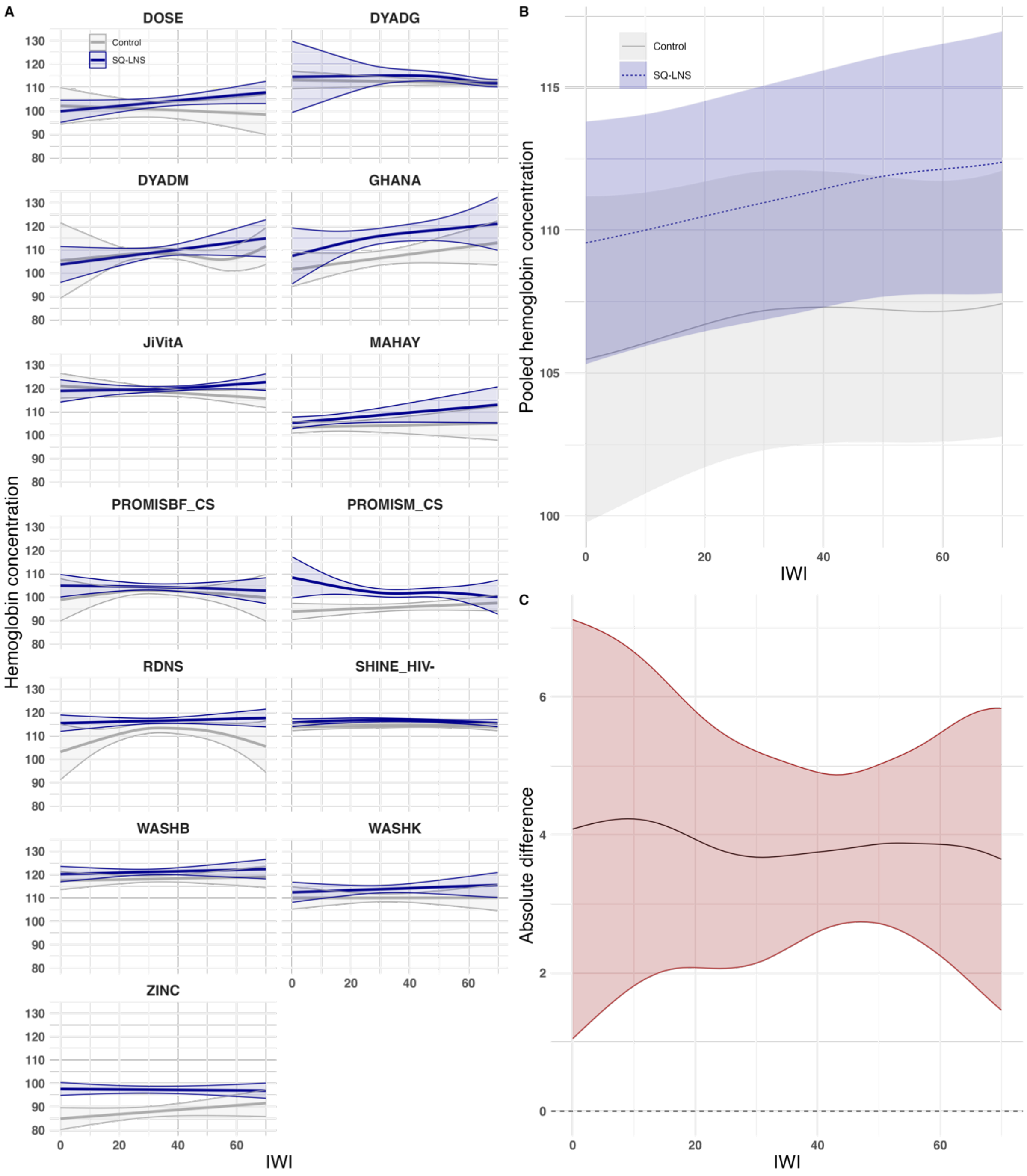

**Figure S25. Probability of anemia by international wealth index and intervention group. A.** Study-specific relationships between the International Wealth Index (IWI) and probability of anemia by intervention group. **B.** Pooled estimates from a two-stage random effects meta-analysis of probability of anemia by IWI and intervention group across all studies in panel A. **C.** Pooled difference in probability of anemia between groups over levels of IWI. Intervention effects conditional on IWI were estimated by subtracting the spline fits for the treated group from the control group within each study. These effects were then pooled using pointwise random-effects meta-analysis with Restricted Maximum Likelihood. In all panels, shaded bands represent pointwise 95% confidence intervals. Intervention excluded maternal supplementation.

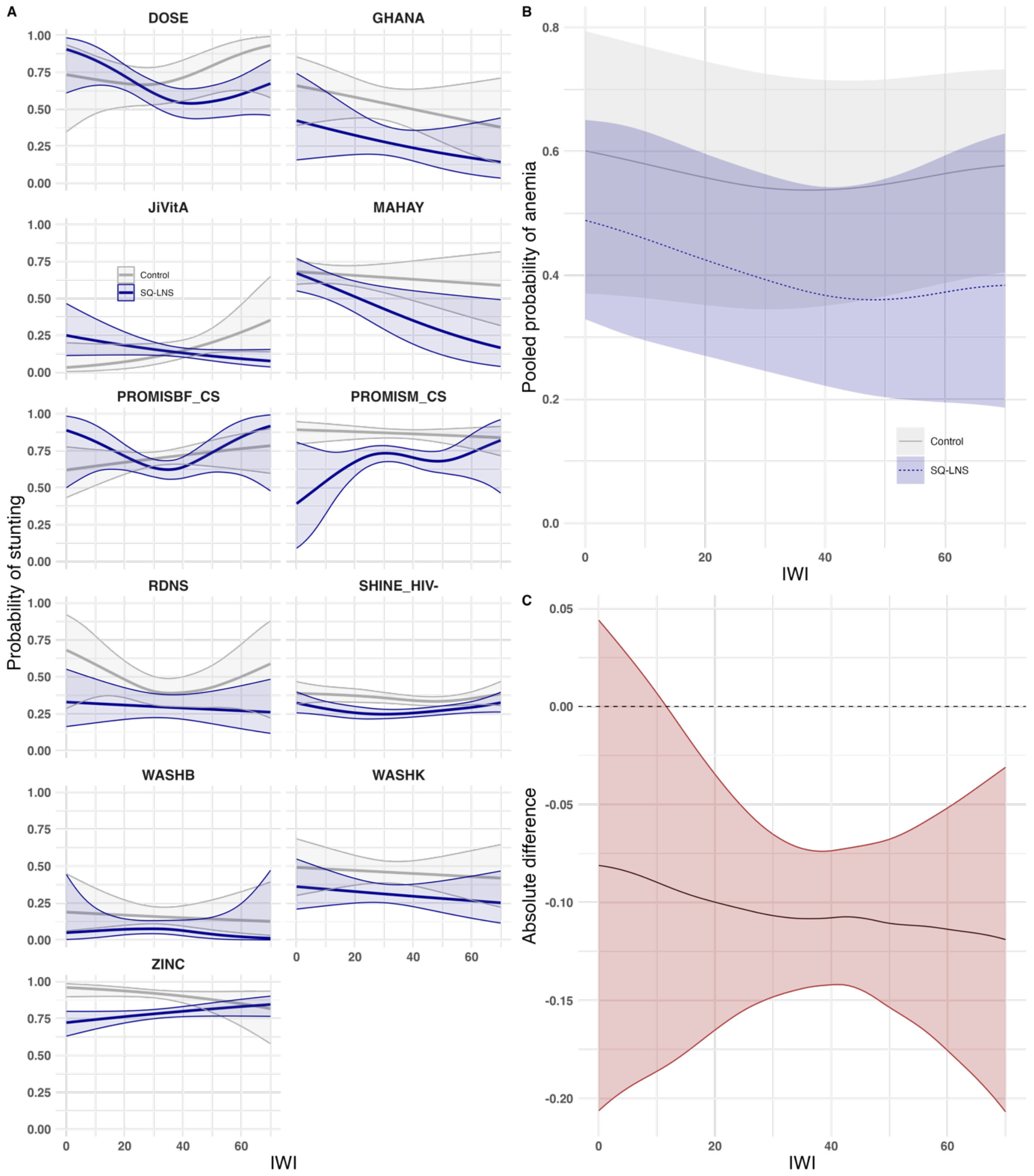

**Figure S26. Probability of anemia by international wealth index and intervention group. A.** Study-specific relationships between the International Wealth Index (IWI) and probability of anemia by intervention group. **B.** Pooled estimates from a two-stage random effects meta-analysis of probability of anemia by IWI and intervention group across all studies in panel A. **C.** Pooled difference in probability of anemia between groups over levels of IWI. Intervention effects conditional on IWI were estimated by subtracting the spline fits for the treated group from the control group within each study. These effects were then pooled using pointwise random-effects meta-analysis with Restricted Maximum Likelihood. In all panels, shaded bands represent pointwise 95% confidence intervals. Intervention included maternal supplementation.

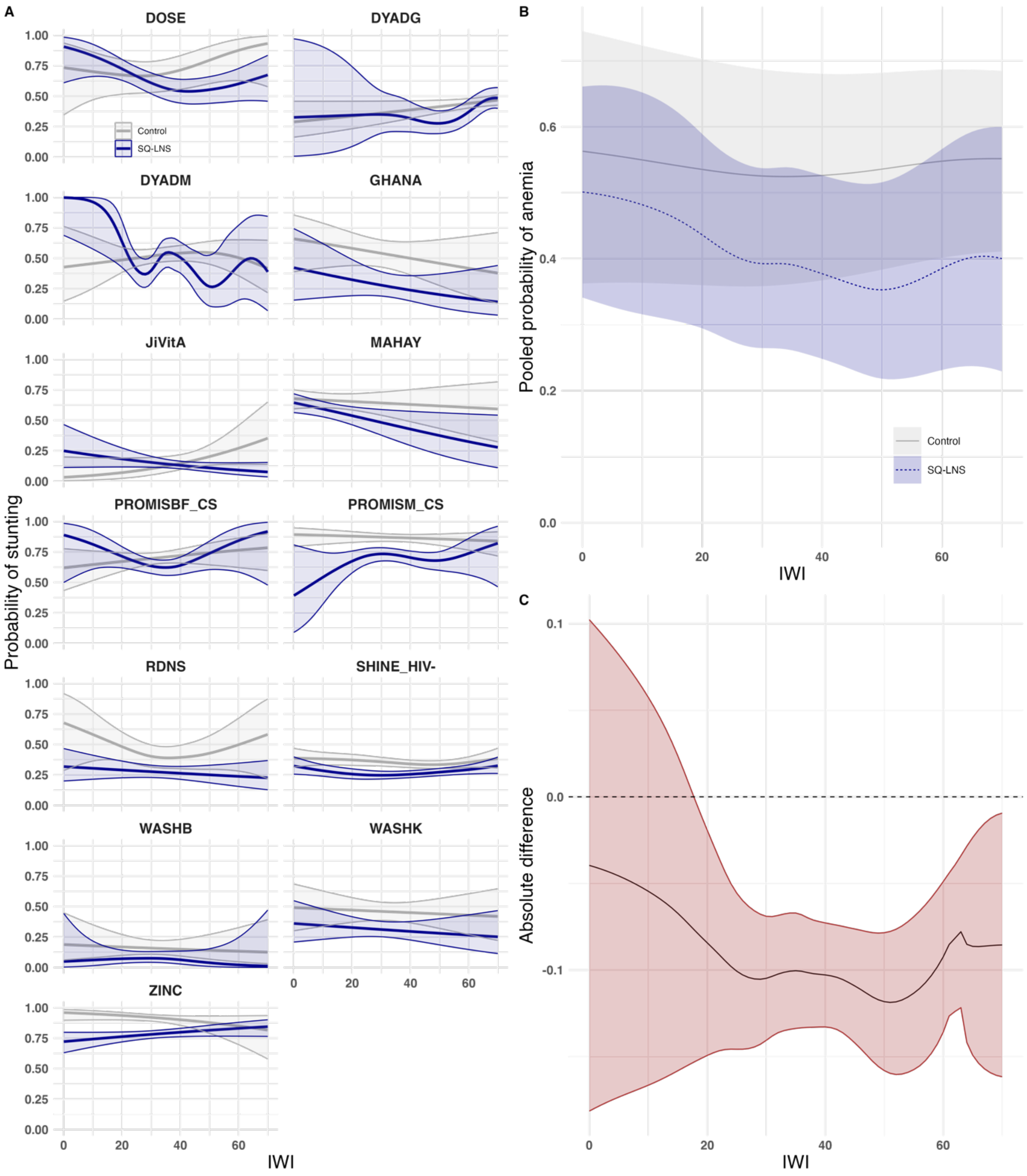

#### Text S2. International Wealth Index.

The following tables and figure illustrate the characteristics of the International Wealth Index. **Tables S2A and S2B** show the distribution of the asset variables included in the construction of the IWI between the control and intervention groups. The median IWI (SD) for the control group ranged from 13.1 (9.79) to 66.6 (13.9). For the SQ-LNS group, the median IWI ranged from 13.1 (8.95) to 65.7 (14.3).  **Tables S3A and S3B** show the distribution of the assets by tertiles of the IWI for each arm. **Figure S27** display the correlation between IWI and study-specific wealth indices.

**Table S2A. Asset variables included in the construction of the International Wealth Index.** Excluded missing values in the intervention variable.

| Study | Arms | IWI Median (SD) | TV N (%) | Phone N (%) | Car N (%) | Bike N (%) | Cheap item N (%) | Expensive item N (%) | Electricity N (%) | Refrigerator N (%) |
|---|---|---|---|---|---|---|---|---|---|---|
| **JiVitA-4** | **Control (N=1423)** | 40.6 (11.4) | 281 (19.7%) | 1082 (76.0%) | 93 (6.5%) | 586 (41.2%) | 1173 (82.4%) | 93 (6.5%) | 974 (68.4%) | NA |
| | **SQ-LNS (N=3145)** | 40.6 (11.1) | 611 (19.4%) | 2395 (76.2%) | 208 (6.6%) | 1294 (41.1%) | 2590 (82.4%) | 208 (6.6%) | 2133 (67.8%) | NA |
| **RDNS** | **Control (N=846)** | 33.7 (11.3) | 248 (29.3%) | 661 (78.1%) | 27 (3.2%) | 417 (49.3%) | 726 (85.8%) | 28 (3.3%) | 492 (58.2%) | 12 (1.4%) |
| | **SQ-LNS (N=1721)** | 33.7 (11.4) | 464 (27.0%) | 1290 (75.0%) | 68 (4.0%) | 952 (55.3%) | 1475 (85.7%) | 72 (4.2%) | 1101 (64.0%) | 36 (2.1%) |
| **WASH-B Bangladesh** | **Control (N=3597)** | 32.8 (10.0) | 1111 (30.9%) | 3108 (86.4%) | NA | 1122 (31.2%) | 3199 (88.9%) | 255 (7.1%) | 1451 (40.3%) | 298 (8.3%) |
| | **SQ-LNS (N=1195)** | 31.0 (9.71) | 357 (29.9%) | 1019 (85.3%) | NA | 401 (33.6%) | 1055 (88.3%) | 69 (5.8%) | 476 (39.8%) | 90 (7.5%) |
| **PROMIS-BF** | **Control (N=918)** | 31.0 (10.3) | 134 (14.6%) | 854 (93.0%) | 6 (0.7%) | 850 (92.6%) | 910 (99.1%) | 11 (1.2%) | 7 (0.8%) | NA |
| | **SQ-LNS (N=864)** | 28.0 (10.4) | 118 (13.7%) | 812 (94.0%) | 1 (0.1%) | 773 (89.5%) | 859 (99.4%) | 4 (0.5%) | 7 (0.8%) | NA |
| **PROMIS-BF CS** | **Control (N=581)** | 30.1 (8.18) | 88 (15.1%) | 537 (92.4%) | 4 (0.7%) | 490 (84.3%) | 571 (98.3%) | 5 (0.9%) | NA | NA |
| | **SQ-LNS (N=576)** | 30.1 (8.56) | 89 (15.5%) | 515 (89.4%) | 2 (0.3%) | 510 (88.5%) | 564 (97.9%) | 4 (0.7%) | NA | NA |
| **iLiNS-Zinc** | **Control (N=672)** | 28.3 (8.72) | 73 (10.9%) | 479 (71.3%) | 0 (0%) | 639 (95.1%) | 650 (96.7%) | 83 (12.4%) | 631 (93.9%) | 1 (0.1%) |
| | **SQ-LNS (N=1975)** | 29.9 (9.20) | 243 (12.3%) | 1555 (78.7%) | 7 (0.4%) | 1908 (96.6%) | 1946 (98.5%) | 223 (11.3%) | 1873 (94.8%) | 6 (0.3%) |
| **Ghana** | **Control (N=96)** | 28.1 (13.8) | 51 (53.1%) | NA | 2 (2.1%) | NA | 79 (82.3%) | 3 (3.1%) | 0 (0%) | NA |
| | **SQ-LNS (N=98)** | 32.8 (13.0) | 68 (69.4%) | NA | 6 (6.1%) | NA | 92 (93.9%) | 7 (7.1%) | 0 (0%) | NA |
| **iLiNS-DYADG** | **Control (N=750)** | 66.6 (13.9) | 633 (84.4%) | 721 (96.1%) | 381 (50.8%) | 81 (10.8%) | 738 (98.4%) | 384 (51.2%) | 744 (99.2%) | 502 (66.9%) |
| | **SQ-LNS (N=363)** | 65.7 (14.3) | 283 (78.0%) | 345 (95.0%) | 185 (51.0%) | 39 (10.7%) | 353 (97.2%) | 185 (51.0%) | 362 (99.7%) | 233 (64.2%) |

| | | | | | | | | | | |
|---|---|---|---|---|---|---|---|---|---|---|
| **Haiti** | **Control (N=156)** | 39.8 (11.7) | 49 (31.4%) | 135 (86.5%) | 0 (0%) | 10 (6.4%) | 140 (89.7%) | 4 (2.6%) | 95 (60.9%) | 8 (5.1%) |
| | **SQ-LNS (N=166)** | 39.8 (13.0) | 69 (41.6%) | 146 (88.0%) | 4 (2.4%) | 17 (10.2%) | 156 (94.0%) | 8 (4.8%) | 76 (45.8%) | 13 (7.8%) |
| **WASH-B Kenya** | **Control (N=5317)** | 30.6 (10.6) | 630 (11.8%) | 4247 (79.9%) | 40 (0.8%) | 2775 (52.2%) | 4980 (93.7%) | 510 (9.6%) | 4937 (92.9%) | NA |
| | **SQ-LNS (N=1498)** | 30.6 (10.7) | 190 (12.7%) | 1218 (81.3%) | 10 (0.7%) | 801 (53.5%) | 1411 (94.2%) | 136 (9.1%) | 1382 (92.3%) | NA |
| **MAHAY** | **Control (N=1709)** | 13.1 (9.79) | 60 (3.5%) | 338 (19.8%) | 9 (0.5%) | 175 (10.2%) | 877 (51.3%) | 21 (1.2%) | 1132 (66.2%) | 7 (0.4%) |
| | **SQ-LNS (N=1729)** | 13.1 (8.95) | 35 (2.0%) | 237 (13.7%) | 5 (0.3%) | 135 (7.8%) | 718 (41.5%) | 8 (0.5%) | 1043 (60.3%) | 7 (0.4%) |
| **iLiNS-DYADM** | **Control (N=453)** | 33.5 (10.8) | 41 (9.1%) | 191 (42.2%) | 8 (1.8%) | 200 (44.2%) | 310 (68.4%) | 13 (2.9%) | 422 (93.2%) | 13 (2.9%) |
| | **SQ-LNS (N=222)** | 33.3 (9.82) | 17 (7.7%) | 90 (40.5%) | 3 (1.4%) | 89 (40.1%) | 155 (69.8%) | 4 (1.8%) | 211 (95.0%) | 6 (2.7%) |
| **iLiNS-DOSE** | **Control (N=257)** | 34.4 (16.1) | NA | 128 (49.8%) | 3 (1.2%) | 118 (45.9%) | 187 (72.8%) | 4 (1.6%) | 223 (86.8%) | 7 (2.7%) |
| | **SQ-LNS (N=761)** | 33.6 (15.4) | NA | 321 (42.2%) | 17 (2.2%) | 305 (40.1%) | 515 (67.7%) | 20 (2.6%) | 695 (91.3%) | 25 (3.3%) |
| **PROMIS-M** | **Control (N=506)** | 28.0 (10.7) | 121 (23.9%) | 455 (89.9%) | 5 (1.0%) | 432 (85.4%) | 499 (98.6%) | 5 (1.0%) | 19 (3.8%) | NA |
| | **SQ-LNS (N=507)** | 28.0 (10.1) | 94 (18.5%) | 452 (89.2%) | 3 (0.6%) | 437 (86.2%) | 498 (98.2%) | 4 (0.8%) | 13 (2.6%) | NA |
| **PROMIS-M CS** | **Control (N=972)** | 35.8 (9.45) | 252 (25.9%) | 854 (87.9%) | 9 (0.9%) | 799 (82.2%) | 950 (97.7%) | 14 (1.4%) | NA | NA |
| | **SQ-LNS (N=955)** | 35.8 (9.50) | 244 (25.5%) | 838 (87.7%) | 4 (0.4%) | 812 (85.0%) | 941 (98.5%) | 13 (1.4%) | NA | NA |
| **SHINE HIV-** | **Control (N=1798)** | 40.6 (17.4) | 557 (31.0%) | 1496 (83.2%) | 49 (2.7%) | 643 (35.8%) | 1588 (88.3%) | 1108 (61.6%) | 1535 (85.4%) | NA |
| | **SQ-LNS (N=1881)** | 41.1 (18.0) | 603 (32.1%) | 1579 (83.9%) | 102 (5.4%) | 749 (39.8%) | 1666 (88.6%) | 1199 (63.7%) | 1599 (85.0%) | NA |

**Table S2B. Asset indicators included in the construction of the International Wealth Index.** Excluded missing values for the intervention variable.

| Study | Arms | Water source quality Low N (%) | Water source quality Medium N (%) | Water source quality High N (%) | Toilet facility quality Low N (%) | Toilet facility quality Medium N (%) | Toilet facility quality High N (%) | Number of rooms 0 or 1 N (%) | Number of rooms 2 N (%) | Number of rooms 3 or more N (%) | Floor quality Low N (%) | Floor quality Medium N (%) | Floor quality High N (%) |
|---|---|---|---|---|---|---|---|---|---|---|---|---|---|
| **JiVitA-4** | **Control (N=1423)** | 268 (18.8%) | 1153 (81.0%) | NA | 338 (23.8%) | 1082 (76.0%) | 1 (0.1%) | NA | NA | NA | 175 (12.3%) | 1246 (87.6%) | NA |
| | **SQ-LNS (N=3145)** | 578 (18.4%) | 2562 (81.5%) | NA | 713 (22.7%) | 2425 (77.1%) | 2 (0.1%) | NA | NA | NA | 360 (11.4%) | 2780 (88.4%) | NA |
| **RDNS** | **Control (N=846)** | NA | 844 (99.8%) | 2 (0.2%) | 236 (27.9%) | 499 (59.0%) | 111 (13.1%) | 163 (19.3%) | 248 (29.3%) | 434 (51.3%) | 749 (88.5%) | 95 (11.2%) | 2 (0.2%) |
| | **SQ-LNS (N=1721)** | NA | 1714 (99.6%) | 7 (0.4%) | 478 (27.8%) | 1032 (60.0%) | 209 (12.1%) | 303 (17.6%) | 509 (29.6%) | 909 (52.8%) | 559 (90.6%) | 162 (9.4%) | 0 (0%) |
| **WASH-B Bangladesh** | **Control (N=3597)** | 918 (25.5%) | 2679 (74.5%) | NA | 204 (5.7%) | 3249 (90.3%) | NA | NA | NA | NA | 3203 (89.0%) | 394 (11.0%) | NA |
| | **SQ-LNS (N=1195)** | 308 (25.8%) | 887 (74.2%) | NA | 66 (5.5%) | 1075 (90.0%) | NA | NA | NA | NA | 1071 (89.6%) | 124 (10.4%) | NA |
| **PROMIS-BF** | **Control (N=918)** | 356 (38.8%) | 369 (40.2%) | 3 (0.3%) | 361 (39.3%) | 552 (60.1%) | NA | NA | NA | NA | 440 (47.9%) | 476 (51.9%) | 2 (0.2%) |
| | **SQ-LNS (N=864)** | 362 (41.9%) | 343 (39.7%) | 4 (0.5%) | 466 (53.9%) | 398 (46.1%) | NA | NA | NA | NA | 446 (51.6%) | 414 (47.9%) | 4 (0.5%) |
| **PROMIS-BF CS** | **Control (N=581)** | 217 (37.3%) | 361 (62.1%) | 2 (0.3%) | 350 (60.2%) | 229 (39.4%) | NA | NA | NA | NA | NA | NA | NA |
| | **SQ-LNS (N=576)** | 202 (35.1%) | 366 (63.5%) | 7 (1.2%) | 329 (57.1%) | 245 (42.5%) | NA | NA | NA | NA | NA | NA | NA |
| **iLiNS-Zinc** | **Control (N=672)** | 525 (78.1%) | 143 (21.3%) | 0 (0%) | 652 (97.0%) | 11 (1.6%) | 5 (0.7%) | 47 (7.0%) | 134 (19.9%) | 487 (72.5%) | 462 (68.8%) | 206 (30.7%) | NA |
| | **SQ-LNS (N=1975)** | 1407 (71.2%) | 539 (27.3%) | 21 (1.1%) | 1923 (97.4%) | 37 (1.9%) | 7 (0.4%) | 237 (12.0%) | 482 (24.4%) | 1248 (63.2%) | 1140 (57.7%) | 827 (41.9%) | NA |
| **Ghana** | **Control (N=96)** | 88 (91.7%) | 2 (2.1%) | NA | 11 (11.5%) | 60 (62.5%) | 20 (20.8%) | 36 (37.5%) | 36 (37.5%) | 19 (19.8%) | NA | NA | NA |
| | **SQ-LNS (N=98)** | 91 (92.9%) | 4 (4.1%) | NA | 5 (5.1%) | 64 (65.3%) | 29 (29.6%) | 42 (42.9%) | 40 (40.8%) | 16 (16.3%) | NA | NA | NA |
| **iLiNS-DYADG** | **Control (N=750)** | 14 (1.9%) | 508 (67.7%) | 226 (30.1%) | 20 (2.7%) | 651 (86.8%) | 77 (10.3%) | 382 (50.9%) | 226 (30.1%) | 140 (18.7%) | 50 (6.7%) | 694 (92.5%) | 4 (0.5%) |
| | **SQ-LNS (N=363)** | 4 (1.1%) | 256 (70.5%) | 103 (28.4%) | 10 (2.8%) | 328 (90.4%) | 25 (6.9%) | 193 (53.2%) | 112 (30.9%) | 58 (16.0%) | 23 (6.3%) | 340 (93.7%) | 0 (0%) |
| **Haiti** | **Control (N=156)** | NA | NA | NA | NA | NA | NA | 65 (41.7%) | 59 (37.8%) | 27 (17.3%) | 20 (12.8%) | 129 (82.7%) | 6 (3.8%) |

| | | | | | | | | | | | | | |
|---|---|---|---|---|---|---|---|---|---|---|---|---|---|
| | **SQ-LNS (N=166)** | NA | NA | NA | NA | NA | NA | 89 (53.6%) | 42 (25.3%) | 34 (20.5%) | 17 (10.2%) | 137 (82.5%) | 11 (6.6%) |
| **WASH-B Kenya** | **Control (N=5317)** | NA | NA | NA | 4128 (77.6%) | 854 (16.1%) | NA | NA | NA | NA | 5007 (94.2%) | 305 (5.7%) | NA |
| | **SQ-LNS (N=1498)** | NA | NA | NA | 1188 (79.3%) | 225 (15.0%) | NA | NA | NA | NA | 1405 (93.8%) | 90 (6.0%) | NA |
| **MAHAY** | **Control (N=1709)** | 1096 (64.1%) | 482 (28.2%) | 8 (0.5%) | 1591 (93.1%) | NA | NA | NA | NA | NA | 1007 (58.9%) | 587 (34.3%) | NA |
| | **SQ-LNS (N=1729)** | 1085 (62.8%) | 480 (27.8%) | 4 (0.2%) | 1602 (92.7%) | NA | NA | NA | NA | NA | 1154 (66.7%) | 449 (26.0%) | NA |
| **iLiNS-DYADM** | **Control (N=453)** | 36 (7.9%) | 353 (77.9%) | 61 (13.5%) | 408 (90.1%) | 36 (7.9%) | 6 (1.3%) | 127 (28.0%) | 192 (42.4%) | 133 (29.4%) | NA | NA | NA |
| | **SQ-LNS (N=222)** | 20 (9.0%) | 171 (77.0%) | 31 (14.0%) | 202 (91.0%) | 18 (8.1%) | 2 (0.9%) | 63 (28.4%) | 98 (44.1%) | 61 (27.5%) | NA | NA | NA |
| **iLiNS-DOSE** | **Control (N=257)** | 19 (7.4%) | 143 (55.6%) | 60 (23.3%) | 213 (82.9%) | 2 (0.8%) | 7 (2.7%) | 47 (18.3%) | 91 (35.4%) | 111 (43.2%) | NA | NA | NA |
| | **SQ-LNS (N=761)** | 54 (7.1%) | 423 (55.6%) | 174 (22.9%) | 635 (83.4%) | 4 (0.5%) | 11 (1.4%) | 146 (19.2%) | 285 (37.5%) | 320 (42.1%) | NA | NA | NA |
| **PROMIS-M** | **Control (N=506)** | 245 (48.4%) | 241 (47.6%) | 15 (3.0%) | 135 (26.7%) | 366 (72.3%) | NA | NA | NA | NA | 370 (73.1%) | 134 (26.5%) | 2 (0.4%) |
| | **SQ-LNS (N=507)** | 249 (49.1%) | 235 (46.4%) | 13 (2.6%) | 112 (22.1%) | 388 (76.5%) | NA | NA | NA | NA | 363 (71.6%) | 143 (28.2%) | 0 (0%) |
| **PROMIS-M CS** | **Control (N=972)** | 402 (41.4%) | 553 (56.9%) | 16 (1.6%) | 262 (27.0%) | 697 (71.7%) | NA | NA | NA | NA | NA | NA | NA |
| | **SQ-LNS (N=955)** | 374 (39.2%) | 556 (58.2%) | 24 (2.5%) | 234 (24.5%) | 704 (73.7%) | NA | NA | NA | NA | NA | NA | NA |
| **SHINE HIV-** | **Control (N=1798)** | 593 (33.0%) | 1048 (58.3%) | 14 (0.8%) | 1068 (59.4%) | 574 (31.9%) | 4 (0.2%) | NA | NA | NA | 815 (45.3%) | 829 (46.1%) | NA |
| | **SQ-LNS (N=1881)** | 558 (29.7%) | 1137 (60.4%) | 27 (1.4%) | 1042 (55.4%) | 669 (35.6%) | 2 (0.1%) | NA | NA | NA | 821 (43.6%) | 895 (47.6%) | NA |

**Table S3A. Asset variables by International Wealth Index tertiles.** We excluded observations with missing values for the intervention variable and IWI.

| Study | IWI tertiles | TV N (%) | Phone N (%) | Car N (%) | Bike N (%) | Cheap item N (%) | Expensive item N (%) | Electricity N (%) | Refrigerator N (%) |
|---|---|---|---|---|---|---|---|---|---|
| JiVitA-4 | T1 (N=1521) | 24 (1.6%) | 589 (38.7%) | 0 (0%) | 228 (15.0%) | 723 (47.5%) | 0 (0%) | 1072 (70.5%) | NA |
| | T2 (N=1520) | 100 (6.6%) | 1381 (90.9%) | 4 (0.3%) | 565 (37.2%) | 1520 (100%) | 4 (0.3%) | 1153 (75.9%) | NA |
| | T3 (N=1520) | 768 (50.5%) | 1507 (99.1%) | 297 (19.5%) | 1087 (71.5%) | 1520 (100%) | 297 (19.5%) | 882 (58.0%) | NA |
| RDNS | T1 (N=856) | 41 (4.8%) | 327 (38.2%) | 1 (0.1%) | 234 (27.3%) | 493 (57.6%) | 1 (0.1%) | 605 (70.7%) | 1 (0.1%) |
| | T2 (N=856) | 211 (24.6%) | 791 (92.4%) | 4 (0.5%) | 424 (49.5%) | 853 (99.6%) | 5 (0.6%) | 576 (67.3%) | 1 (0.1%) |
| | T3 (N=855) | 460 (53.8%) | 833 (97.4%) | 90 (10.5%) | 711 (83.2%) | 855 (100%) | 94 (11.0%) | 412 (48.2%) | 46 (5.4%) |
| WASH-B Bangladesh | T1 (N=1598) | 35 (2.2%) | 980 (61.3%) | NA | 267 (16.7%) | 1062 (66.5%) | 1 (0.1%) | 445 (27.8%) | 0 (0%) |
| | T2 (N=1597) | 280 (17.5%) | 1557 (97.5%) | NA | 414 (25.9%) | 1595 (99.9%) | 40 (2.5%) | 1077 (67.4%) | 12 (0.8%) |
| | T3 (N=1597) | 1153 (72.2%) | 1590 (99.6%) | NA | 842 (52.7%) | 1597 (100%) | 283 (17.7%) | 405 (25.4%) | 376 (23.5%) |
| PROMIS-BF | T1 (N=594) | 11 (1.9%) | 503 (84.7%) | 0 (0%) | 508 (85.5%) | 582 (98.0%) | 1 (0.2%) | 5 (0.8%) | NA |
| | T2 (N=594) | 33 (5.6%) | 573 (96.5%) | 1 (0.2%) | 558 (93.9%) | 593 (99.8%) | 3 (0.5%) | 6 (1.0%) | NA |
| | T3 (N=594) | 208 (35.0%) | 590 (99.3%) | 6 (1.0%) | 557 (93.8%) | 594 (100%) | 11 (1.9%) | 3 (0.5%) | NA |
| PROMIS-BF CS | T1 (N=385) | 2 (0.5%) | 285 (74.0%) | 0 (0%) | 294 (76.4%) | 364 (94.5%) | 0 (0%) | NA | NA |
| | T2 (N=384) | 0 (0%) | 384 (100%) | 0 (0%) | 358 (93.2%) | 384 (100%) | 0 (0%) | NA | NA |
| | T3 (N=384) | 174 (45.3%) | 381 (99.2%) | 6 (1.6%) | 346 (90.1%) | 384 (100%) | 9 (2.3%) | NA | NA |

| | | | | | | | | | |
|---|---|---|---|---|---|---|---|---|---|
| **iLiNS-Zinc** | **T1 (N=879)** | 1 (0.1%) | 375 (42.7%) | 0 (0%) | 816 (92.8%) | 840 (95.6%) | 21 (2.4%) | 844 (96.0%) | 1 (0.1%) |
| | **T2 (N=878)** | 24 (2.7%) | 798 (90.9%) | 1 (0.1%) | 874 (99.5%) | 878 (100%) | 40 (4.6%) | 864 (98.4%) | 0 (0%) |
| | **T3 (N=878)** | 291 (33.1%) | 861 (98.1%) | 6 (0.7%) | 857 (97.6%) | 878 (100%) | 245 (27.9%) | 796 (90.7%) | 6 (0.7%) |
| **Ghana** | **T1 (N=53)** | 14 (26.4%) | NA | 0 (0%) | NA | 40 (75.5%) | 0 (0%) | 0 (0%) | NA |
| | **T2 (N=53)** | 50 (94.3%) | NA | 0 (0%) | NA | 53 (100%) | 0 (0%) | 0 (0%) | NA |
| | **T3 (N=53)** | 51 (96.2%) | NA | 8 (15.1%) | NA | 53 (100%) | 10 (18.9%) | 0 (0%) | NA |
| **iLiNS-DYADG** | **T1 (N=371)** | 182 (49.1%) | 328 (88.4%) | 10 (2.7%) | 46 (12.4%) | 351 (94.6%) | 11 (3.0%) | 369 (99.5%) | 54 (14.6%) |
| | **T2 (N=370)** | 365 (98.6%) | 368 (99.5%) | 226 (61.1%) | 46 (12.4%) | 370 (100%) | 226 (61.1%) | 368 (99.5%) | 319 (86.2%) |
| | **T3 (N=370)** | 369 (99.7%) | 370 (100%) | 330 (89.2%) | 28 (7.6%) | 370 (100%) | 332 (89.7%) | 369 (99.7%) | 362 (97.8%) |
| **Haiti** | **T1 (N=107)** | 5 (4.7%) | 74 (69.2%) | 0 (0%) | 2 (1.9%) | 81 (75.7%) | 0 (0%) | 56 (52.3%) | 1 (0.9%) |
| | **T2 (N=107)** | 28 (26.2%) | 103 (96.3%) | 0 (0%) | 8 (7.5%) | 107 (100%) | 0 (0%) | 75 (70.1%) | 2 (1.9%) |
| | **T3 (N=107)** | 85 (79.4%) | 103 (96.3%) | 4 (3.7%) | 17 (15.9%) | 107 (100%) | 12 (11.2%) | 40 (37.4%) | 18 (16.8%) |
| **WASH-B Kenya** | **T1 (N=2272)** | 0 (0%) | 1124 (49.5%) | 0 (0%) | 489 (21.5%) | 1852 (81.5%) | 28 (1.2%) | 2067 (91.0%) | NA |
| | **T2 (N=2272)** | 37 (1.6%) | 2202 (96.9%) | 0 (0%) | 1719 (75.7%) | 2272 (100%) | 14 (0.6%) | 2221 (97.8%) | NA |
| | **T3 (N=2271)** | 783 (34.5%) | 2139 (94.2%) | 50 (2.2%) | 1368 (60.2%) | 2267 (99.8%) | 604 (26.6%) | 2031 (89.4%) | NA |
| **MAHAY** | **T1 (N=1146)** | 0 (0%) | 1 (0.1%) | 0 (0%) | 28 (2.4%) | 182 (15.9%) | 0 (0%) | 405 (35.3%) | 0 (0%) |
| | **T2 (N=1146)** | 0 (0%) | 74 (6.5%) | 0 (0%) | 97 (8.5%) | 532 (46.4%) | 0 (0%) | 846 (73.8%) | 1 (0.1%) |

| | | | | | | | | | |
|---|---|---|---|---|---|---|---|---|---|
| | **T3 (N=1146)** | 95 (8.3%) | 500 (43.6%) | 14 (1.2%) | 185 (16.1%) | 881 (76.9%) | 29 (2.5%) | 924 (80.6%) | 13 (1.1%) |
| **iLiNS-DYADM** | **T1 (N=222)** | 0 (0%) | 0 (0%) | 0 (0%) | 33 (14.9%) | 45 (20.3%) | 0 (0%) | 216 (97.3%) | 0 (0%) |
| | **T2 (N=222)** | 6 (2.7%) | 72 (32.4%) | 1 (0.5%) | 108 (48.6%) | 198 (89.2%) | 2 (0.9%) | 215 (96.8%) | 0 (0%) |
| | **T3 (N=222)** | 52 (23.4%) | 209 (94.1%) | 10 (4.5%) | 148 (66.7%) | 222 (100%) | 15 (6.8%) | 196 (88.3%) | 19 (8.6%) |
| **iLiNS-DOSE** | **T1 (N=340)** | NA | 12 (3.5%) | 0 (0%) | 47 (13.8%) | 96 (28.2%) | 0 (0%) | 308 (90.6%) | 0 (0%) |
| | **T2 (N=339)** | NA | 177 (52.2%) | 3 (0.9%) | 174 (51.3%) | 296 (87.3%) | 7 (2.1%) | 325 (95.9%) | 1 (0.3%) |
| | **T3 (N=339)** | NA | 260 (76.7%) | 17 (5.0%) | 202 (59.6%) | 310 (91.4%) | 17 (5.0%) | 285 (84.1%) | 31 (9.1%) |
| **PROMIS-M** | **T1 (N=338)** | 0 (0%) | 254 (75.1%) | 0 (0%) | 272 (80.5%) | 322 (95.3%) | 0 (0%) | 4 (1.2%) | NA |
| | **T2 (N=338)** | 26 (7.7%) | 322 (95.3%) | 0 (0%) | 301 (89.1%) | 338 (100%) | 0 (0%) | 10 (3.0%) | NA |
| | **T3 (N=337)** | 189 (56.1%) | 331 (98.2%) | 8 (2.4%) | 296 (87.8%) | 337 (100%) | 9 (2.7%) | 18 (5.3%) | NA |
| **PROMIS-M CS** | **T1 (N=632)** | 7 (1.1%) | 415 (65.7%) | 0 (0%) | 503 (79.6%) | 598 (94.6%) | 1 (0.2%) | NA | NA |
| | **T2 (N=632)** | 35 (5.5%) | 627 (99.2%) | 0 (0%) | 537 (85.0%) | 632 (100%) | 1 (0.2%) | NA | NA |
| | **T3 (N=631)** | 444 (70.4%) | 623 (98.7%) | 13 (2.1%) | 543 (86.1%) | 631 (100%) | 24 (3.8%) | NA | NA |
| **SHINE HIV-** | **T1 (N=1227)** | 57 (4.6%) | 700 (57.0%) | 0 (0%) | 264 (21.5%) | 806 (65.7%) | 339 (27.6%) | 848 (69.1%) | NA |
| | **T2 (N=1226)** | 301 (24.6%) | 1169 (95.4%) | 21 (1.7%) | 433 (35.3%) | 1222 (99.7%) | 849 (69.2%) | 1117 (91.1%) | NA |
| | **T3 (N=1226)** | 802 (65.4%) | 1206 (98.4%) | 130 (10.6%) | 695 (56.7%) | 1226 (100%) | 1119 (91.3%) | 1169 (95.4%) | NA |

**Table S3B. Asset indicators by International Wealth Index tertiles.** We excluded observations with missing values for the intervention variable and IWI.

| Study | IWI tertiles | Water source quality Low N (%) | Water source quality Medium N (%) | Water source quality High N (%) | Toilet facility quality Low N (%) | Toilet facility quality Medium N (%) | Toilet facility quality High N (%) | Number of rooms 0 or 1 N (%) | Number of rooms 2 N (%) | Number of rooms 3 or more N (%) | Floor quality Low N (%) | Floor quality Medium N (%) | Floor quality High N (%) |
|---|---|---|---|---|---|---|---|---|---|---|---|---|---|
| JiVitA-4 | T1 (N=1521) | 582 (38.3%) | 939 (61.7%) | NA | 676 (44.4%) | 845 (55.6%) | 0 (0%) | NA | NA | NA | 485 (31.9%) | 1036 (68.1%) | 1036 (68.1%) |
| | T2 (N=1520) | 243 (16.0%) | 1277 (84.0%) | NA | 358 (23.6%) | 1162 (76.4%) | 0 (0%) | NA | NA | NA | 45 (3.0%) | 1475 (97.0%) | 1475 (97.0%) |
| | T3 (N=1520) | 21 (1.4%) | 1499 (98.6%) | NA | 17 (1.1%) | 1500 (98.7%) | 3 (0.2%) | NA | NA | NA | 5 (0.3%) | 5 (0.3%) | 1515 (99.7%) |
| RDNS | T1 (N=856) | NA | 855 (99.9%) | 1 (0.1%) | 513 (59.9%) | 341 (39.8%) | 1 (0.1%) | 309 (36.1%) | 365 (42.6%) | 181 (21.1%) | 851 (99.4%) | 5 (0.6%) | 0 (0%) |
| | T2 (N=856) | NA | 854 (99.8%) | 2 (0.2%) | 190 (22.2%) | 641 (74.9%) | 25 (2.9%) | 128 (15.0%) | 312 (36.4%) | 416 (48.6%) | 845 (98.7%) | 11 (1.3%) | 0 (0%) |
| | T3 (N=855) | NA | 849 (99.3%) | 6 (0.7%) | 11 (1.3%) | 549 (64.2%) | 294 (34.4%) | 29 (3.4%) | 80 (9.4%) | 746 (87.3%) | 612 (71.6%) | 241 (28.2%) | 2 (0.2%) |
| WASH-B Bangladesh | T1 (N=1598) | 519 (32.5%) | 1079 (67.5%) | NA | 171 (10.7%) | 1328 (83.1%) | NA | NA | NA | NA | 1598 (100%) | 0 (0%) | NA |
| | T2 (N=1597) | 498 (31.2%) | 1099 (68.8%) | NA | 96 (6.0%) | 1434 (89.8%) | NA | NA | NA | NA | 1576 (98.7%) | 21 (1.3%) | NA |
| | T3 (N=1597) | 209 (13.1%) | 1388 (86.9%) | NA | 3 (0.2%) | 1562 (97.8%) | NA | NA | NA | NA | 1100 (68.9%) | 497 (31.1%) | NA |
| PROMIS-BF | T1 (N=594) | 395 (66.5%) | 186 (31.3%) | 0 (0%) | 437 (73.6%) | 157 (26.4%) | NA | NA | NA | NA | 510 (85.9%) | 84 (14.1%) | 0 (0%) |
| | T2 (N=594) | 271 (45.6%) | 222 (37.4%) | 3 (0.5%) | 304 (51.2%) | 288 (48.5%) | NA | NA | NA | NA | 246 (41.4%) | 346 (58.2%) | 2 (0.3%) |
| | T3 (N=594) | 52 (8.8%) | 304 (51.2%) | 4 (0.7%) | 86 (14.5%) | 505 (85.0%) | NA | NA | NA | NA | 130 (21.9%) | 460 (77.4%) | 4 (0.7%) |
| PROMIS-BF CS | T1 (N=385) | 341 (54.0%) | 291 (46.0%) | 0 (0%) | 418 (66.1%) | 214 (33.9%) | NA | NA | NA | NA | NA | NA | NA |
| | T2 (N=384) | 290 (45.9%) | 342 (54.1%) | 0 (0%) | 31 (4.9%) | 601 (95.1%) | NA | NA | NA | NA | NA | NA | NA |
| | T3 (N=384) | 135 (21.4%) | 457 (72.4%) | 39 (6.2%) | 47 (7.4%) | 584 (92.6%) | NA | NA | NA | NA | NA | NA | NA |

| | | | | | | | | | | | | | |
|---|---|---|---|---|---|---|---|---|---|---|---|---|---|
| **iLiNS-Zinc** | **T1 (N=879)** | 745 (84.8%) | 134 (15.2%) | 0 (0%) | 877 (99.8%) | 2 (0.2%) | 0 (0%) | 216 (24.6%) | 334 (38.0%) | 329 (37.4%) | 834 (94.9%) | 45 (5.1%) | NA |
| | **T2 (N=878)** | 637 (72.6%) | 241 (27.4%) | 0 (0%) | 868 (98.9%) | 10 (1.1%) | 0 (0%) | 62 (7.1%) | 175 (19.9%) | 641 (73.0%) | 660 (75.2%) | 218 (24.8%) | NA |
| | **T3 (N=878)** | 550 (62.6%) | 307 (35.0%) | 21 (2.4%) | 830 (94.5%) | 36 (4.1%) | 12 (1.4%) | 6 (0.7%) | 107 (12.2%) | 765 (87.1%) | 108 (12.3%) | 770 (87.7%) | NA |
| **Ghana** | **T1 (N=53)** | 53 (100%) | 0 (0%) | NA | 4 (7.5%) | 49 (92.5%) | 0 (0%) | 35 (66.0%) | 17 (32.1%) | 1 (1.9%) | NA | NA | NA |
| | **T2 (N=53)** | 53 (100%) | 0 (0%) | NA | 1 (1.9%) | 49 (92.5%) | 3 (5.7%) | 19 (35.8%) | 33 (62.3%) | 1 (1.9%) | NA | NA | NA |
| | **T3 (N=53)** | 52 (98.1%) | 1 (1.9%) | NA | 0 (0%) | 8 (15.1%) | 45 (84.9%) | 5 (9.4%) | 21 (39.6%) | 27 (50.9%) | NA | NA | NA |
| **iLiNS-DYADG** | **T1 (N=371)** | 11 (3.0%) | 304 (81.9%) | 56 (15.1%) | 17 (4.6%) | 347 (93.5%) | 7 (1.9%) | 240 (64.7%) | 84 (22.6%) | 47 (12.7%) | 36 (9.7%) | 335 (90.3%) | 0 (0%) |
| | **T2 (N=370)** | 6 (1.6%) | 293 (79.2%) | 71 (19.2%) | 12 (3.2%) | 338 (91.4%) | 20 (5.4%) | 233 (63.0%) | 89 (24.1%) | 48 (13.0%) | 33 (8.9%) | 337 (91.1%) | 0 (0%) |
| | **T3 (N=370)** | 1 (0.3%) | 167 (45.1%) | 202 (54.6%) | 1 (0.3%) | 294 (79.5%) | 75 (20.3%) | 102 (27.6%) | 165 (44.6%) | 103 (27.8%) | 4 (1.1%) | 362 (97.8%) | 4 (1.1%) |
| **Haiti** | **T1 (N=107)** | NA | NA | NA | NA | NA | NA | 74 (69.2%) | 29 (27.1%) | 2 (1.9%) | 34 (31.8%) | 70 (65.4%) | 2 (1.9%) |
| | **T2 (N=107)** | NA | NA | NA | NA | NA | NA | 66 (61.7%) | 31 (29.0%) | 7 (6.5%) | 2 (1.9%) | 103 (96.3%) | 2 (1.9%) |
| | **T3 (N=107)** | NA | NA | NA | NA | NA | NA | 14 (13.1%) | 41 (38.3%) | 52 (48.6%) | 1 (0.9%) | 93 (86.9%) | 13 (12.1%) |
| **WASH-B Kenya** | **T1 (N=2272)** | NA | NA | NA | 2138 (94.1%) | 92 (4.0%) | NA | NA | NA | NA | 2267 (99.8%) | 2 (0.1%) | NA |
| | **T2 (N=2272)** | NA | NA | NA | 2207 (97.1%) | 65 (2.9%) | NA | NA | NA | NA | 2267 (99.8%) | 5 (0.2%) | NA |
| | **T3 (N=2271)** | NA | NA | NA | 971 (42.8%) | 922 (40.6%) | NA | NA | NA | NA | 1878 (82.7%) | 388 (17.1%) | NA |
| **MAHAY** | **T1 (N=1146)** | 730 (63.7%) | 175 (15.3%) | 0 (0%) | 905 (79.0%) | NA | NA | NA | NA | NA | 904 (78.9%) | 1 (0.1%) | NA |
| | **T2 (N=1146)** | 795 (69.4%) | 336 (29.3%) | 0 (0%) | 1146 (100%) | NA | NA | NA | NA | NA | 950 (82.9%) | 196 (17.1%) | NA |
| | **T3 (N=1146)** | 656 (57.2%) | 451 (39.4%) | 12 (1.0%) | 1142 (99.7%) | NA | NA | NA | NA | NA | 307 (26.8%) | 839 (73.2%) | NA |
| **iLiNS-DYADM** | **T1 (N=222)** | 28 (12.6%) | 167 (75.2%) | 26 (11.7%) | 220 (99.1%) | 1 (0.5%) | 0 (0%) | 115 (51.8%) | 77 (34.7%) | 30 (13.5%) | NA | NA | NA |

| | | | | | | | | | | | | | |
|---|---|---|---|---|---|---|---|---|---|---|---|---|---|
| | **T2 (N=222)** | 15 (6.8%) | 182 (82.0%) | 25 (11.3%) | 210 (94.6%) | 11 (5.0%) | 1 (0.5%) | 58 (26.1%) | 109 (49.1%) | 55 (24.8%) | NA | NA | NA |
| | **T3 (N=222)** | 13 (5.9%) | 169 (76.1%) | 39 (17.6%) | 172 (77.5%) | 42 (18.9%) | 7 (3.2%) | 15 (6.8%) | 99 (44.6%) | 108 (48.6%) | NA | NA | NA |
| **iLiNS-DOSE** | **T1 (N=340)** | 51 (15.0%) | 266 (78.2%) | 6 (1.8%) | 323 (95.0%) | 0 (0%) | 0 (0%) | 107 (31.5%) | 142 (41.8%) | 73 (21.5%) | NA | NA | NA |
| | **T2 (N=339)** | 21 (6.2%) | 254 (74.9%) | 56 (16.5%) | 330 (97.3%) | 1 (0.3%) | 0 (0%) | 57 (16.8%) | 138 (40.7%) | 144 (42.5%) | NA | NA | NA |
| | **T3 (N=339)** | 1 (0.3%) | 46 (13.6%) | 172 (50.7%) | 195 (57.5%) | 5 (1.5%) | 18 (5.3%) | 29 (8.6%) | 96 (28.3%) | 214 (63.1%) | NA | NA | NA |
| **PROMIS-M** | **T1 (N=338)** | 238 (70.4%) | 99 (29.3%) | 0 (0%) | 171 (50.6%) | 166 (49.1%) | NA | NA | NA | NA | 332 (98.2%) | 5 (1.5%) | 0 (0%) |
| | **T2 (N=338)** | 129 (38.2%) | 206 (60.9%) | 2 (0.6%) | 62 (18.3%) | 274 (81.1%) | NA | NA | NA | NA | 283 (83.7%) | 54 (16.0%) | 1 (0.3%) |
| | **T3 (N=337)** | 127 (37.7%) | 171 (50.7%) | 26 (7.7%) | 14 (4.2%) | 314 (93.2%) | NA | NA | NA | NA | 118 (35.0%) | 218 (64.7%) | 1 (0.3%) |
| **PROMIS-M CS** | **T1 (N=632)** | 341 (54.0%) | 291 (46.0%) | 0 (0%) | 418 (66.1%) | 214 (33.9%) | NA | NA | NA | NA | NA | NA | NA |
| | **T2 (N=632)** | 290 (45.9%) | 342 (54.1%) | 0 (0%) | 31 (4.9%) | 601 (95.1%) | NA | NA | NA | NA | NA | NA | NA |
| | **T3 (N=631)** | 135 (21.4%) | 457 (72.4%) | 39 (6.2%) | 47 (7.4%) | 584 (92.6%) | NA | NA | NA | NA | NA | NA | NA |
| **SHINE HIV-** | **T1 (N=1227)** | 579 (47.2%) | 389 (31.7%) | 2 (0.2%) | 880 (71.7%) | 85 (6.9%) | 0 (0%) | NA | NA | NA | 852 (69.4%) | 101 (8.2%) | NA |
| | **T2 (N=1226)** | 338 (27.6%) | 874 (71.3%) | 9 (0.7%) | 879 (71.7%) | 337 (27.5%) | 1 (0.1%) | NA | NA | NA | 647 (52.8%) | 556 (45.4%) | NA |
| | **T3 (N=1226)** | 234 (19.1%) | 922 (75.2%) | 30 (2.4%) | 351 (28.6%) | 821 (67.0%) | 5 (0.4%) | NA | NA | NA | 137 (11.2%) | 1067 (87.0%) | NA |

**Figure S27. Pearson correlation between the international wealth index and study-specific wealth indices.** The study-specific wealth indices were constructed by each study investigators.

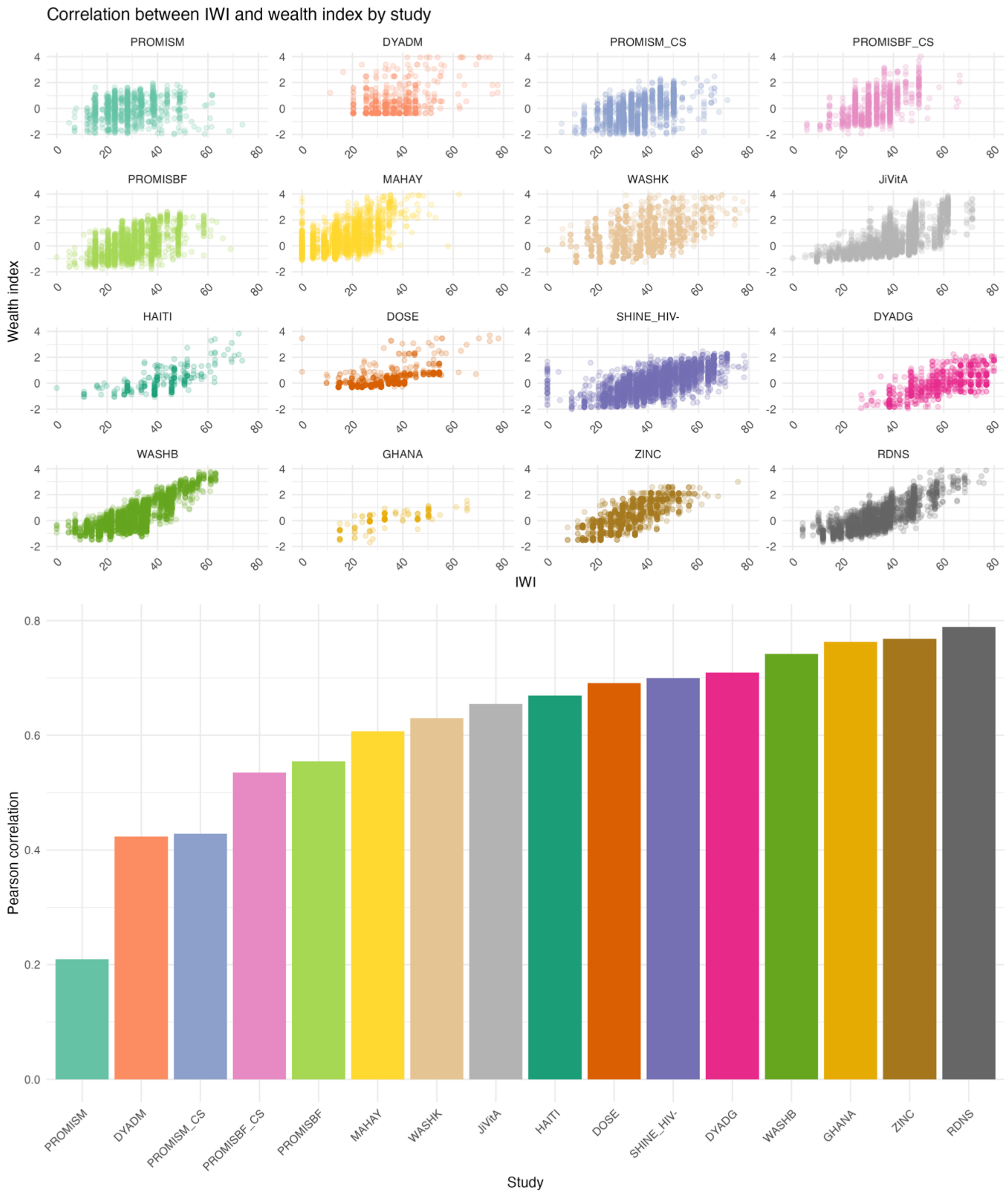

#### Text S3. Descriptive statistics of child outcomes.

The mean LAZ ranged from -2.24 (MAHAY) to -0.41 (Ghana) for the control group and from -2.21 (MAHAY) to -0.13 (Ghana) for the SQ-LNS group at endline (**Figure S28**). The mean WLZ ranged from -1.16 (JiVitA-4) to 0.09 (WASHB-Kenya) for the control group and from -1.08 (JiVitA-4) to 0.16 (WASHB-Kenya) for the SQ-LNS group. The prevalence of stunting ranged from 12.75% (DYADG) to 58.50% (MAHAY) in the control group and from 8.65% (DYADG) to 57.46% (MAHAY) in the SQ-LNS group (**Figure S28 and Table S3**). For wasting, prevalence ranged from 1.48% (WASHB-Kenya) to 16.38% (JiViTA-4) in the control group and from ~1% (PROMISM) to 14.86% (JiViTA-4) in the SQ-LNS group. Severe stunting prevalence varied from 1.01% (DYADG) to 23.37% (MAHAY) in the control group and from 1.73% (DYADG) to 23.68% (MAHAY) in the SQ-LNS group.

Language scores ranged from -0.24 (ZINC) to 0.07 (Haiti) for the control group and from -0.07 (Haiti) to 0.12 (ZINC) for the SQ-LNS group (**Figure S29**). Gross motor scores ranged from -0.05 (SHINE_HIV-) to 0.08 (JiVitA-4) for the control group and from -0.02 (DYADM) to 0.09 (WASHB-Bangladesh) for the SQ-LNS group. Fine motor scores ranged from -0.11 (JiVitA-4) to -0.0008 (DYADG) for the control group and from 0.02 (MAHAY) to 0.06 (DYADM) for the SQ-LNS group. Executive function scores ranged from -0.07 (RDNS) to 0.08 (DOSE) for the control group and from -0.06 (DYADM) to 0.03 (RDNS) for the SQ-LNS group. Socioemotional scores ranged from -0.23 (ZINC) to 0.04 (MAHAY) for the control group and from -0.04 (MAHAY) to 0.12 (ZINC) for the SQ-LNS group.

The average hemoglobin concentration ranged from 88.56 g/L (ZINC) to 118.12 g/L (WASHB-Bangladesh) for the control group and from 97.11 g/L (ZINC) to 121.14 g/L (WASHB-Bangladesh) for the SQ-LNS group (**Figure S30**).

Anemia prevalence ranged from 15.75% (JiViTA-4) to 91.06% (ZINC) in the control group and from 6.87% (WASHB-Bangladesh) to 79.12% (ZINC) in the SQ-LNS group (**Figure S30 and Table S3**).

It should be noted that PROMISM and PROMISBF were longitudinal studies with monthly measurements spanning from 6 to 23 months and from 6 to 18 months of age respectively. As such both studies had differential timing of anthropometric measurements[1]. Therefore, we took the last visit where most of the children were at least 22 months old (PROMISM) or 16 months old (PROMISBF) as "endline" to align with the other trials, similar with the previous individual participant data meta-analysis, which explains the difference in prevalence between this study and the original PROMISM trial. We also defined wasting as WLZ < -2, while in PROMISM and PROMISBF, wasting was defined as WLZ < −2 or MUAC < 125 mm the presence of bilateral pitting edema resulting in higher wasting prevalences. When we joined the longitudinal (PROMISM) and repeated cross-sectional (PROMISM_CS) data, the total wasting prevalence was around 7% which was consistent with the previous individual participant data meta-analysis[2]. For PROMISBF, the wasting prevalences were consistent for both studies[1].

**Figure S28. (Left to right) Mean length-for-age z score, percentage of stunting, percentage of severe wasting, mean weight-for-length z score, percentage of wasting, and percentage of severe wasting among children aged 6-24 months at endline between the control and intervention group (SQ-LNS) for each study.** Intervention included maternal supplementation.

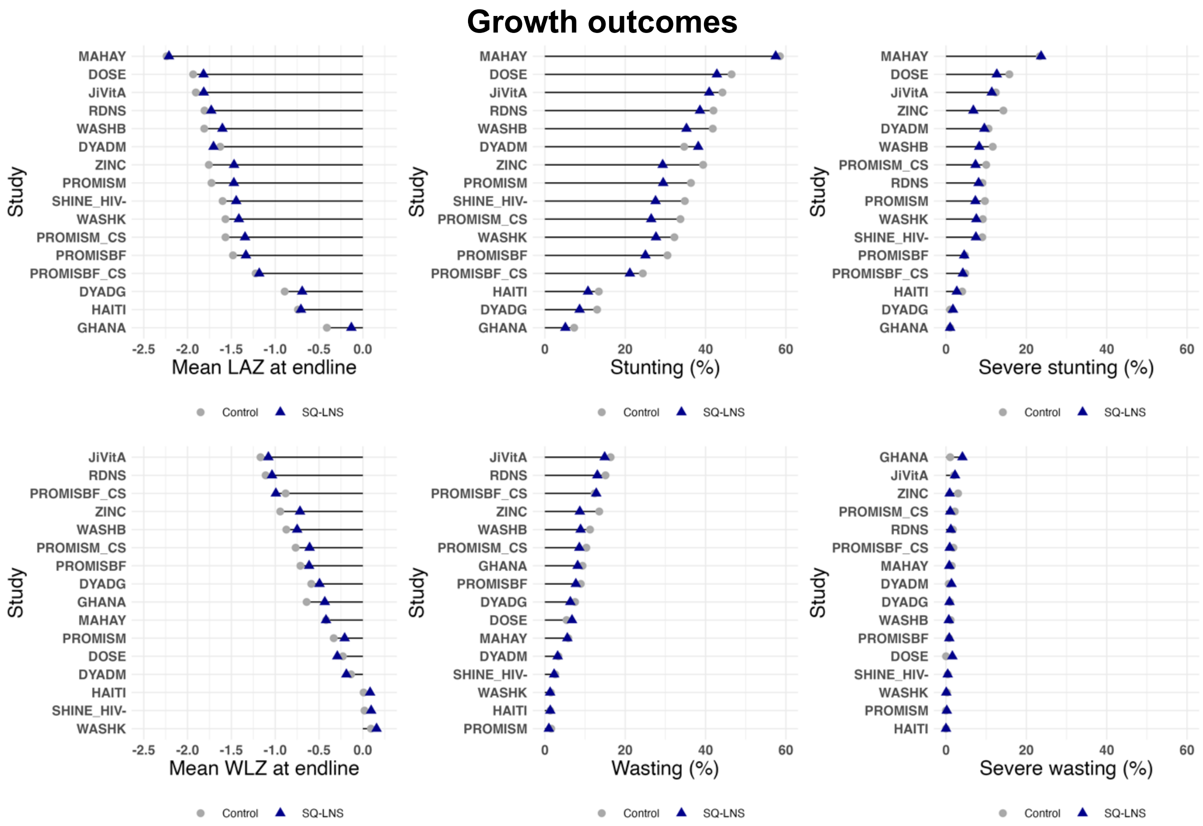

**Figure S29. Mean estimates of the different child developmental outcomes among children aged 6-24 months at endline between the control and intervention group (SQ-LNS) for each study.** The solid black circle denotes a developmental Z-score of zero, with development scores above zero further from the center and scores below zero closer to the center. Intervention included maternal supplementation. Blue colors indicate the intervention group, and grey colors indicate the control group. Development scores are internal to each study distribution of the study-specific indicator.

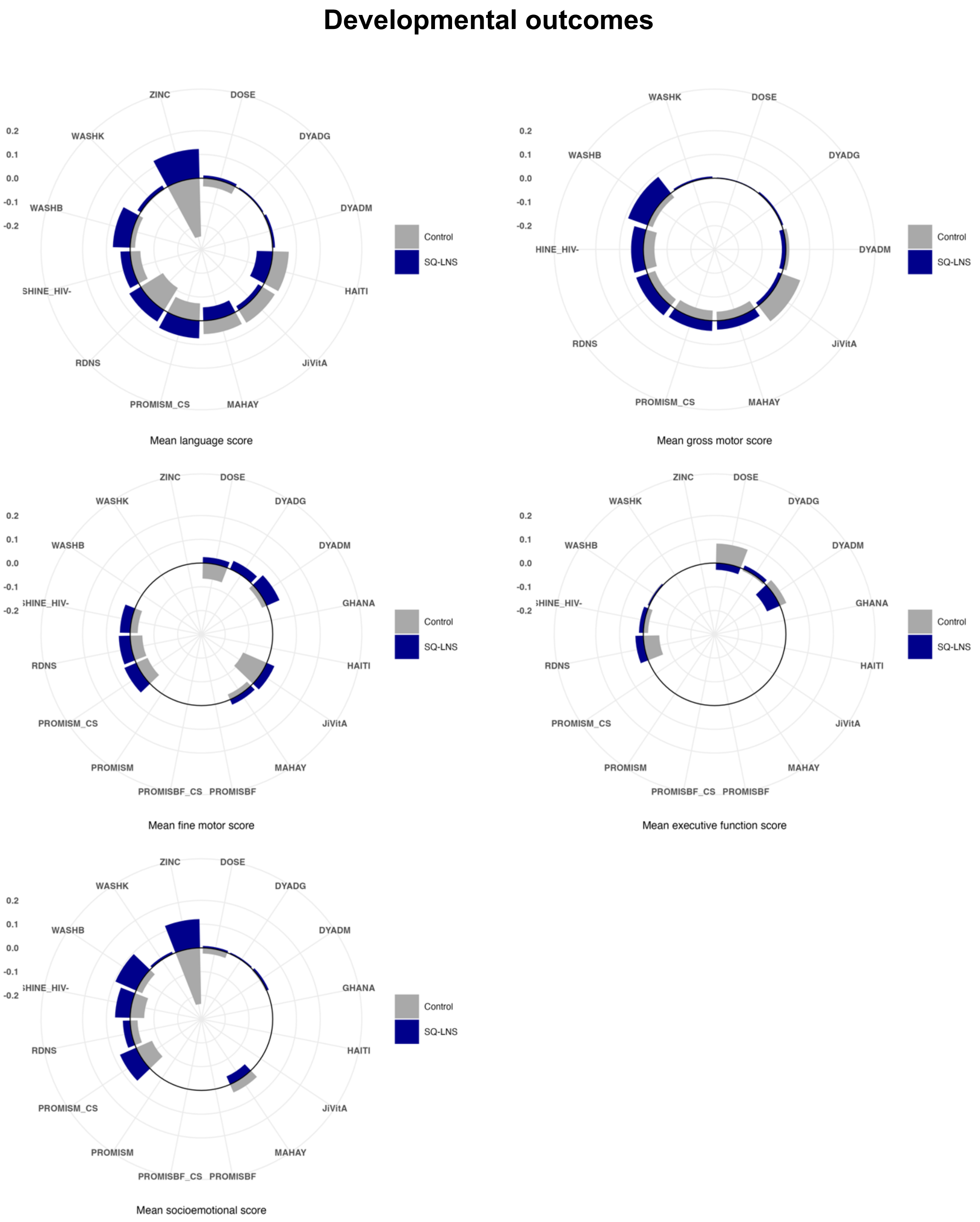

**Figure S30. Mean hemoglobin concentration and percentage of anemia at endline among children aged 6-24 months between the control and intervention group (SQ-LNS) for each study.** Intervention included maternal supplementation.

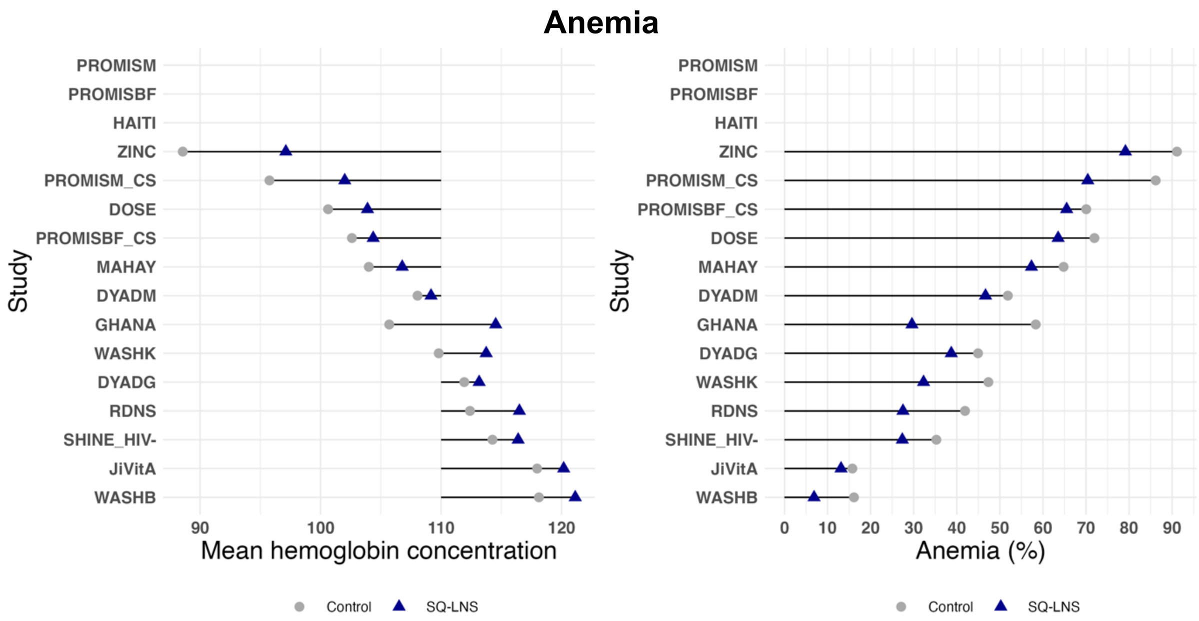

#### Text S4. Study-specific wealth inequalities in adverse child outcomes.

*Study-specific inequalities*

Study-specific analysis for stunting revealed that most studies showed relative and absolute inequalities favoring higher SEP households (stunting more prevalent among those with the lowest SEP) (**Table S3**). This trend was seen in JiViTA-4, RDNS, WASHB-Bangladesh and WASHB-Kenya across both arms. Notably, in JiViTA-4 and RDNS, both relative and absolute inequalities were more pronounced in the SQ-LNS group, while in WASHB-Kenya, absolute inequality were lower in the SQ-LNS group. In some instances, inequalities were only observed in the control group (MAHAY), or the SQ-LNS group (PROMISM and PROMISM_CS).

For wasting, similar inequalities were observed in all arms in WASHB-Bangladesh. In JiViTA-4, RDNS, PROMISBF_CS, DYADG (relative inequality) and DYADM (relative inequality), inequalities were observed in the SQ-LNS group. In MAHAY, both relative and absolute inequalities were found only in the control group.

For severe stunting, relative and absolute inequalities were observed in both arms for RDNS, WASHB-Bangladesh, and WASHB-Kenya. In MAHAY, absolute inequality was seen only in the SQ-LNS group. Relative inequality was observed in both arms for PROMISM_CS, while absolute inequality was found only in the SQ-LNS group. In contrast, for DOSE, both relative and absolute inequalities appeared only in the control group.

For anemia, most trials showed no significant inequalities. However, in a few cases, relative and absolute inequalities were observed. In JiViTA-4, inequalities in the control group favored those with the lowest SEP (anemia was more prevalent among higher SEP households). In MAHAY, relative inequality favored lower SEP households in the SQ-LNS group. Conversely, in DOSE, both relative and absolute inequalities favored higher SEP households in the SQ-LNS group.

**Table S4. Prevalence, and relative and absolute wealth inequalities in adverse child outcomes.** Red font signifies inequality that is favorable to the wealthiest households (i.e., the adverse outcome is more prelavent among the poorest households). Blue font signifies that the inequality is favorable to the poorest households (i.e., the adverse outcome is more prevalent among the wealthiest households). Grey font signifies no significant inequalities were observed.

| Study | Adverse outcome | Prevalence at endline | | RII (95% CI) | | SII (95% CI) | |
|---|---|---|---|---|---|---|---|
| | | Control (%) | SQ-LNS (%) | Control | SQ-LNS | Control | SQ-LNS |
| JiVitA-4 | Stunting | 44.15 | 40.93 | 1.33 (1.05; 1.68) | 1.70 (1.43; 2.02) | 13.30 (1.30; 25.30) | 24.60 (15.60; 33.50) |
| | Wasting | 16.38 | 14.86 | 1.12 (0.67; 1.88) | 1.81 (1.31; 2.50) | 1.90 (-7.00; 10.90) | 10.10 (3.80; 16.50) |
| | Severe stunting | 12.32 | 11.40 | 1.85 (1.00; 3.42) | 3.11 (2.05; 4.71) | 8.70 (-1.40; 18.80) | 16.90 (8.70; 25.10) |
| | Anemia | 15.75 | 13.16 | 0.15 (0.15; 0.87) | 2.10 (0.88; 5.04) | -22.40 (-40.10; -4.80) | 11.30 (-4.10; 26.70) |
| RDNS | Stunting | 41.96 | 38.60 | 1.77 (1.26; 2.48) | 2.15 (1.79; 2.60) | 25.10 (10.00; 40.20) | 31.60 (23.30; 39.90) |
| | Wasting | 15.09 | 13.06 | 1.22 (0.60; 2.48) | 2.13 (1.41; 3.20) | 3.10 (-7.90; 14.00) | 10.50 (4.60; 16.40) |
| | Severe stunting | 9.08 | 8.12 | 4.31 (2.02; 9.19) | 2.77 (1.37; 5.60) | 15.10 (6.40; 23.70) | 9.00 (2.40; 15.70) |
| | Anemia | 41.91 | 27.50 | 1.22 (0.67; 2.22) | 1.21 (0.69; 2.11) | 8.30 (-17.20; 33.80) | 5.20 (-10.30; 20.70) |
| WASH-B Bangladesh | Stunting | 41.79 | 35.25 | 2.21 (1.85; 2.65) | 2.58 (1.86; 3.57) | 33.60 (25.70; 41.40) | 33.60 (21.90; 45.20) |
| | Wasting | 11.24 | 8.89 | 2.52 (1.73; 3.67) | 3.24 (1.61; 6.52) | 10.50 (6.10; 15.00) | 10.60 (3.70; 17.60) |
| | Severe stunting | 11.64 | 8.27 | 2.68 (1.77; 4.06) | 3.88 (1.53; 9.88) | 11.70 (6.50; 16.90) | 11.40 (2.50; 20.40) |
| | Anemia | 16.13 | 6.87 | 1.55 (0.35; 6.95) | 1.56 (0.26; 9.21) | 7.10 (-17.00; 31.30) | 3.00 (-9.00; 15.00) |
| PROMIS-BF | Stunting | 30.53 | 25.03 | 0.99 (0.67; 1.48) | 0.96 (0.58; 1.59) | -0.20 (-12.40; 12.00) | -1.00 (-13.60; 11.70) |
| | Wasting | 8.93 | 7.71 | 1.28 (0.52; 3.15) | 1.35 (0.55; 3.32) | 2.20 (-6.10; 10.50) | 2.30 (-4.40; 9.00) |
| | Severe stunting | 4.70 | 4.52 | 1.78 (0.49; 6.47) | 1.16 (0.42; 3.20) | 2.80 (-3.90; 9.50) | 0.70 (-4.00; 5.30) |
| | Anemia | NA | NA | NA | NA | NA | NA |

| Study | Adverse outcome | Prevalence at endline | | RII (95% CI) | | SII (95% CI) | |
|---|---|---|---|---|---|---|---|
| | | Control (%) | SQ-LNS (%) | Control (%) | SQ-LNS (%) | Control (%) | SQ-LNS (%) |
| PROMIS-BF CS | **Stunting** | 30.53 | 21.26 | 1.13 (0.50; 2.56) | 1.19 (0.53; 2.67) | 2.90 (-17.10; 23.00) | 3.80 (-13.60; 21.20) |
| | **Wasting** | 12.39 | 12.85 | 0.31 (0.10; 1.04) | 4.31 (1.33; 13.93) | -14.40 (-30.20; 1.30) | 20.60 (1.50; 39.70) |
| | **Severe stunting** | 4.78 | 4.21 | 2.26 (0.21; 24.31) | 0.55 (0.07; 4.28) | 4.00 (-8.30; 16.30) | -2.50 (-11.10; 6.00) |
| | **Anemia** | 69.95 | 65.62 | 0.84 (0.66; 1.07) | 1.02 (0.78; 1.33) | -12.10 (-29.30; 5.00) | 1.40 (-15.90; 18.80) |
| iLiNS-Zinc | **Stunting** | 39.18 | 29.24 | 1.39 (0.87; 2.22) | 1.03 (0.73; 1.45) | 12.90 (-5.90; 31.60) | 0.80 (-9.40; 10.90) |
| | **Wasting** | 13.46 | 8.58 | 2.33 (0.97; 5.59) | 2.00 (0.96; 4.16) | 11.30 (0.10; 22.40) | 6.00 (-0.40; 12.50) |
| | **Severe stunting** | 14.07 | 6.73 | 1.08 (0.46; 2.55) | 1.22 (0.50; 2.94) | 1.10 (-10.80; 13.00) | 1.30 (-4.60; 7.30) |
| | **Anemia** | 91.06 | 79.12 | 1.07 (0.95; 1.20) | 0.91 (0.82; 1.00) | 5.90 (-4.80; 16.60) | -7.70 (-15.90; 0.40) |
| Ghana | **Stunting** | 2.63 | 2.41 | 4.16 (0.08; 208.95) | 0.06 (0.001; 6.88) | 3.80 (-8.10; 15.80) | -5.10 (-14.60; 4.30) |
| | **Wasting** | 10.53 | 8.43 | 2.24 (0.16; 30.97) | 1.07 (0.10; 12.09) | 8.60 (-20.10; 37.30) | 0.60 (-20.10; 21.30) |
| | **Severe stunting** | 1.32 | 0.00 | 1106.50 (118.84; 10302.20) | 1.00 (0.47; 2.11) | 11.60 (-12.30; 35.60) | 0.00 (-0.00; 0.00) |
| | **Anemia** | 53.95 | 26.51 | 1.45 (0.66; 3.20) | 1.764 (0.43; 7.32) | 19.60 (-21.50; 60.80) | 15.30 (-23.60; 54.10) |
| iLiNS-DYADG | **Stunting** | 12.75 | 8.65 | 1.04 (0.20; 5.38) | 1.10 (0.07; 16.26) | 0.50 (-21.20; 22.30) | 0.80 (-24.50; 26.10) |
| | **Wasting** | 7.54 | 6.34 | 1.35 (0.18; 10.25) | 0.15 (0.01; 2.81) | 2.60 (-17.50; 22.70) | -5.90 (-10.10; -1.60) |
| | **Severe stunting** | 1.01 | 1.73 | 9.14 (0.16; 513.57) | 68.51 (4.10; 1143.94) | 7.20 (-23.30; 37.60) | 66.40 (-80.90; 213.80) |

| | | | | | | | |
|---|---|---|---|---|---|---|---|
| | **Anemia** | 44.92 | 38.72 | 0.48 (0.21; 1.11) | 0.48 (0.15; 1.48) | -24.20 (-44.10; - 4.30) | -21.30 (-44.90; 2.40) |
| **Study** | **Adverse outcome** | **Prevalence at endline** | | **RII (95% CI)** | | **SII (95% CI)** | |
| | | **Control (%)** | **SQ-LNS (%)** | **Control (%)** | **SQ-LNS (%)** | **Control (%)** | **SQ-LNS (%)** |
| **Haiti** | **Stunting** | 13.42 | 10.81 | 0.41 (0.04; 3.94) | 2.87 (0.59; 13.90) | -9.70 (-29.20; 9.90) | 15.00 (-14.10; 44.10) |
| | **Wasting** | 1.34 | 1.35 | 0.00 (0.00; 11.73) | 0.03 (0.00; 29.39) | -4.00 (-10.00; 2.00) | -2.40 (-6.20; 1.50) |
| | **Severe stunting** | 4.03 | 2.70 | 0.93 (0.01; 113.65) | 8.78 (0.27); 286.96) | -0.30 (-18.90; 18.40) | 10.30 (-16.30; 36.80) |
| | **Anemia** | NA | NA | NA | NA | NA | NA |
| **WASH-B Kenya** | **Stunting** | 32.24 | 27.66 | 1.69 (1.42; 2.01) | 1.7 (1.23; 2.37) | 17.10 (11.40; 22.80) | 14.90 (5.40; 24.50) |
| | **Wasting** | 1.48 | 1.31 | 2.03 (0.76; 5.46) | 4.95 (0.61; 40.07) | 1.10 (-0.40; 2.60) | 2.20 (-1.20; 5.70) |
| | **Severe stunting** | 9.17 | 7.55 | 2.41 (1.61; 3.61) | 5.47 (2.10; 14.25) | 8.30 (4.40; 12.20) | 13.70 (4.60; 22.90) |
| | **Anemia** | 47.33 | 32.29 | 1.16 (0.68; 1.98) | 1.04 (0.53; 2.05) | 6.90 (-18.40; 32.30) | 1.40 (-20.20; 23.00) |
| **MAHAY** | **Stunting** | 58.50 | 57.46 | 1.28 (1.02; 1.59) | 1.31 (0.90; 1.91) | 13.30 (2.10; 24.50) | 14.30 (-3.50; 32.20) |
| | **Wasting** | 5.83 | 5.59 | 14.78 (2.58; 84.45) | 4.11 (0.43; 39.35) | 8.60 (4.20; 13.10) | 5.30 (-0.80; 11.30) |
| | **Severe stunting** | 23.37 | 23.68 | 1.65 (0.91; 3.01) | 2.20 (0.91; 5.31) | 10.20 (-0.20; 20.50) | 14.70 (1.70; 27.60) |
| | **Anemia** | 64.80 | 57.33 | 1.08 (0.66; 1.79) | 2.06 (0.93; 4.59) | 5.10 (-25.70; 35.90) | 32.90 (4.50; 61.30) |
| **iLiNS-DYADM** | **Stunting** | 34.02 | 38.07 | 1.31 (0.78; 2.18) | 1.36 (0.66; 2.77) | 9.40 (-9.00; 27.80) | 11.90 (-17.00; 40.80) |
| | **Wasting** | 3.20 | 3.21 | 2.78 (0.32; 24.40) | 36.81 (1.93; 702.26) | 3.70 (-5.20; 12.60) | 18.50 (-8.20; 45.10) |
| | **Severe stunting** | 10.50 | 9.63 | 2.59 (0.93; 7.21) | 0.56 (0.09; 3.56) | 11.10 (-2.50; 24.60) | -5.30 (-21.60; 11.00) |

| | Anemia | 51.99 | 46.41 | 0.87 (0.59; 1.27) | 1.16 (0.61; 2.23) | -7.20 (-26.80; 12.40) | 7.10 (-23.50; 37.70) |
|---|---|---|---|---|---|---|---|
| **Study** | **Adverse outcome** | **Prevalence at endline** | | **RII (95% CI)** | | **SII (95% CI)** | |
| | | **Control (%)** | **SQ-LNS (%)** | **Control (%)** | **SQ-LNS (%)** | **Control (%)** | **SQ-LNS (%)** |
| **iLiNS-DOSE** | **Stunting** | 46.47 | 42.82 | 1.37 (0.89; 2.11) | 1.23 (0.93; 1.62) | 15.00 (-6.00; 36.00) | 8.80 (-3.40; 21.00) |
| | **Wasting** | 5.39 | 6.75 | 1.22 (0.24; 6.14) | 2.22 (0.86; 5.75) | 1.10 (-7.90; 10.10) | 5.70 (-1.60; 13.00) |
| | **Severe stunting** | 15.77 | 12.64 | 2.97 (1.16; 7.56) | 1.27 (0.68; 2.34) | 18.60 (0.90; 36.20) | 3.00 (-5.00; 11.10) |
| | **Anemia** | 71.95 | 63.52 | 0.74 (0.50; 1.10) | 1.42 (1.03; 1.95) | -21.70 (-49.40; 6.00) | 22.40 (2.20; 42.70) |
| **PROMIS-M** | **Stunting** | 36.36 | 29.45 | 1.35 (0.84; 2.18) | 2.20 (1.21; 4.00) | 10.90 (-6.00; 27.70) | 22.50 (5.70; 39.20) |
| | **Wasting** | 1.58 | 0.99 | 7.00 (0.30; 161.53) | 1.15 (0.13; 9.98) | 2.90 (-2.20; 8.10) | 0.10 (-2.00; 2.30) |
| | **Severe stunting** | 9.68 | 7.31 | 1.71 (0.56; 5.17) | 1.49 (0.44; 5.05) | 5.00 (-5.30; 15.40) | 2.90 (-5.80; 11.50) |
| | **Anemia** | NA | NA | NA | NA | NA | NA |
| **PROMIS-M CS** | **Stunting** | 33.61 | 26.12 | 1.29 (0.89; 1.86) | 2.48 (1.56; 3.94) | 8.80 (-4.70; 22.20) | 27.10 (12.00; 42.30) |
| | **Wasting** | 10.26 | 8.32 | 1.81 (0.90; 3.66) | 1.13 (0.51; 2.55) | 6.60 (-1.80; 15.00) | 1.10 (-5.90; 8.00) |
| | **Severe stunting** | 10.05 | 7.17 | 2.21 (1.10; 4.46) | 2.47 (1.18; 5.18) | 8.90 (-0.50; 18.20) | 7.40 (0.50; 14.30) |
| | **Anemia** | 86.40 | 70.37 | 1.04 (0.93; 1.16) | 0.98 (0.81; 1.19) | 3.40 (-6.00; 12.90) | -1.50 (-14.90; 11.90) |
| **SHINE HIV-** | **Stunting** | 34.84 | 27.55 | 1.09 (0.88; 1.35) | 1.18 (0.93; 1.50) | 3.10 (-4.70; 10.80) | 4.80 (-2.20; 11.70) |
| | **Wasting** | 2.58 | 2.30 | 1.46 (0.60; 3.50) | 1.26 (0.47; 3.37) | 1.00 (-1.60; 3.70) | 0.60 (-1.90; 3.00) |
| | **Severe stunting** | 9.03 | 7.45 | 1.49 (0.95; 2.34) | 1.58 (0.95; 2.63) | 3.90 (-0.80; 8.50) | 3.70 (-0.90; 8.40) |
| | **Anemia** | 35.26 | 27.35 | 1.14 (0.92; 1.43) | 0.95 (0.73; 1.25) | 4.80 (-3.30; 13.00) | -1.30 (-8.50; 6.00) |

| Study | Adverse outcome | Prevalence at endline | | RII (95% CI) | | SII (95% CI) | |
|---|---|---|---|---|---|---|---|
| | | Control (%) | SQ-LNS (%) | Control (%) | SQ-LNS (%) | Control (%) | SQ-LNS (%) |
| **Common effect** | **Stunting** | 21.95 | 18.49 | 1.49 (1.38; 1.61) | 1.62 (1.49; 1.76) | 13.43 (10.56; 16.31) | 13.27 (10.36; 16.18) |
| | **Wasting** | 4.26 | 3.64 | 1.73 (1.40; 2.14) | 1.95 (1.61; 2.38) | 2.23 (1.14; 3.32) | 1.46 (0.33; 2.59) |
| | **Severe stunting** | 4.98 | 5.82 | 2.21 (1.85; 2.65) | 2.06 (1.69; 2.51) | 7.66 (5.67; 9.65) | 5.02 (3.05; 6.99) |
| | **Anemia** | 56.34 | 36.14 | 1.02 (0.96; 1.10) | 0.98 (0.91; 1.06) | -0.12 (-4.41; 4.16) | 0.35 (-3.49; 4.20) |
| **Random effect** | **Stunting** | 21.95 | 18.49 | 1.41 (1.22; 1.62) | 1.56 (1.30; 1.87) | 11.03 (5.46; 16.60) | 13.28 (6.66; 19.90) |
| | **Wasting** | 4.26 | 3.64 | 1.65 (1.21; 2.24) | 1.95 (1.61; 2.38) | 3.24 (0.69; 5.78) | 3.47 (0.71; 6.23) |
| | **Severe stunting** | 4.98 | 5.82 | 2.28 (1.77; 2.93) | 2.00 (1.49; 2.68) | 7.70 (5.32; 10.08) | 5.84 (2.65; 9.03) |
| | **Anemia** | 56.34 | 36.14 | 1.02 (0.96; 1.10) | 1.05 (0.92; 1.19) | -2.39 (-9.22; 4.44) | 2.11 (-3.21; 7.54) |
| **$I^2$** | **Stunting** | 0% | 0% | 62% | 68% | 71% | 80% |
| | **Wasting** | 0% | 0% | 40% | 11% | 59% | 70% |
| | **Severe stunting** | 0% | 0% | 64% | 51% | 0% | 51% |
| | **Anemia** | 0% | 0% | 28% | 30% | 45% | 40% |

#### Text S5. Sex-stratified analyses.

*Descriptive statistics*

In general, both female and male children benefit from SQ-LNS (**Figures S31-S33**), with females generally showing slightly better growth and development outcomes than males. For gross motor skills, males demonstrated better improvements (**Figure S32**). Both sexes tend to benefit from SQ-LNS in reducing anemia in majority of the studies (**Figures S33**).

*Wealth-related inequalities in growth, development and anemia outcomes*

Overall, for stunting, we observed relative (**Figure S34**) and absolute inequalities (**Figure S35**) favoring children born into the wealthiest households (stunting was more prevalent among the poorest households) in both intervention arms.

For wasting, pooled relative inequalities (**Figure S34**) were observed in the control group, while both relative and absolute inequalities were evident among boys in both arms. Relative inequality was higher among boys compared to girls. No significant absolute inequality (**Figure S35**) was observed among females in either arm, whereas absolute inequality favoring those with higher wealth level was observed among boys. For severe stunting, we found overall relative (**Figure S34**) and absolute (**Figure S35**) inequalities favoring wealthier households, with relative inequality being more pronounced among girls compared to boys. No significant inequalities were observed for anemia in either intervention arm or across sexes **(Figures S34 and S35**).

*Effect of SQ-LNS on different child outcomes by wealth stratified by sex*

**Figures S36-S38** show the effect of SQ-LNS on growth, development and anemia among girls. **Figures S39-S41** are the findings among boys. In general, the SQ-LNS intervention significantly improved growth (**Figures S36 and S39**), developmental (**Figures S37 and S40**), and anemia (**Figures S38 and S41**) outcomes for both sexes. However, among girls, the pooled intervention effect between SQ-LNS and the control group did not vary across the IWI range for any child outcomes (pooled interaction p-value not significant) (**Figures S36-S38**). In contrast, among boys, the intervention effect varied significantly across the IWI range for WLZ (**Figure S39B**, pooled interaction p-value = 0.048) and socioemotional scores (**Figure S40E**, pooled interaction p-value = 0.020).

**Figure S31. Mean length-for-age z-score, percentage of stunting, and percentage of severe stunting among children aged 6–24 months in the control group and intervention group (SQ-LNS), presented separately for females (left panel) and males (right panel) in each study.** Intervention included maternal supplementation.

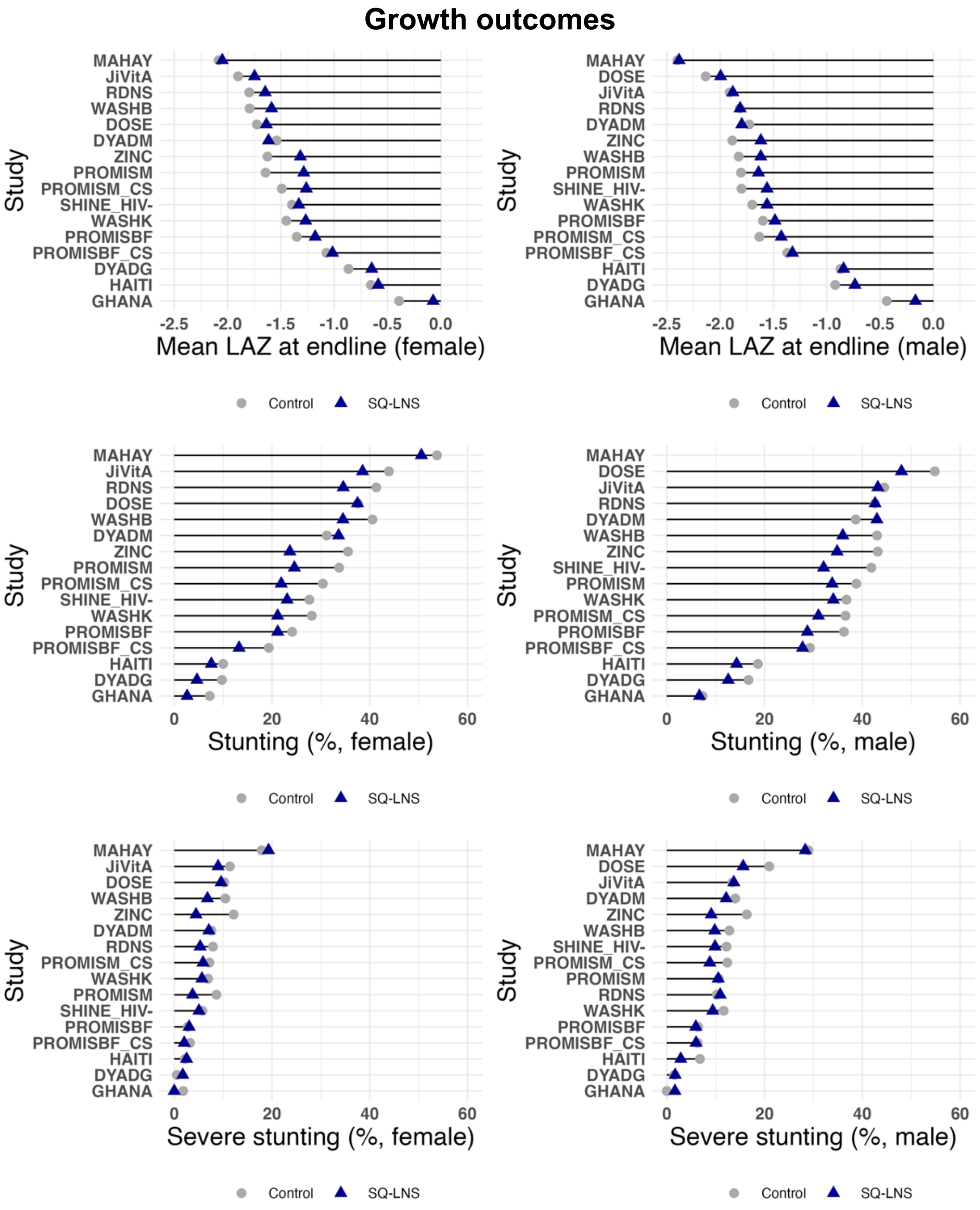

**Figure S32. Developmental outcomes among children aged 6–24 months in the control group and intervention group (SQ-LNS), presented separately for females (left panel) and males (right panel) in each study.** Intervention included maternal supplementation. Development scores are internal to each study distribution of the study-specific indicator.

#### Development outcomes

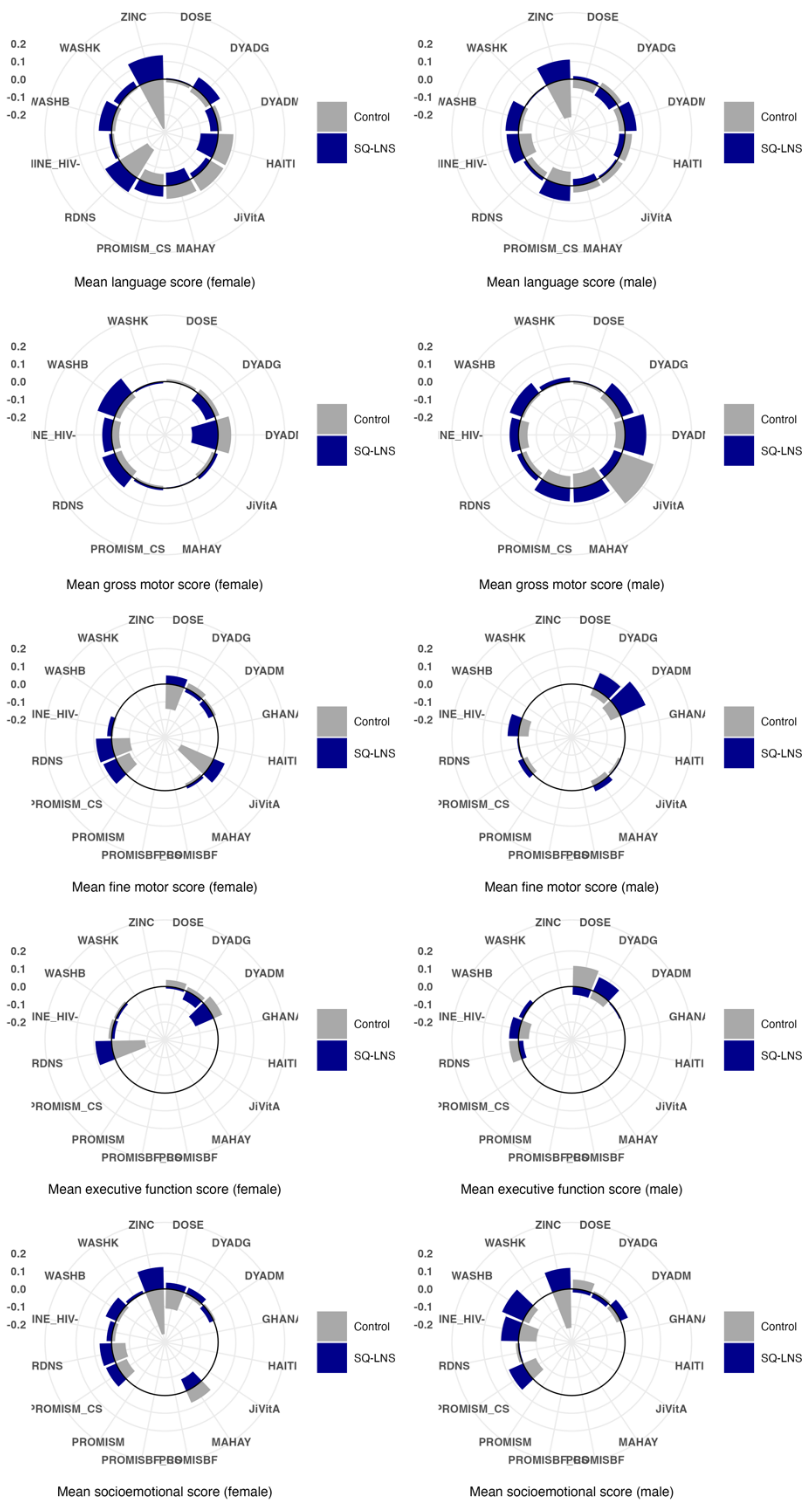

**Figure S33. Mean hemoglobin concentration and percentage of anemia among children aged 6–24 months in the control group and intervention group (SQ-LNS), presented separately for females (left panel) and males (right panel) in each study.** Intervention included maternal supplementation.

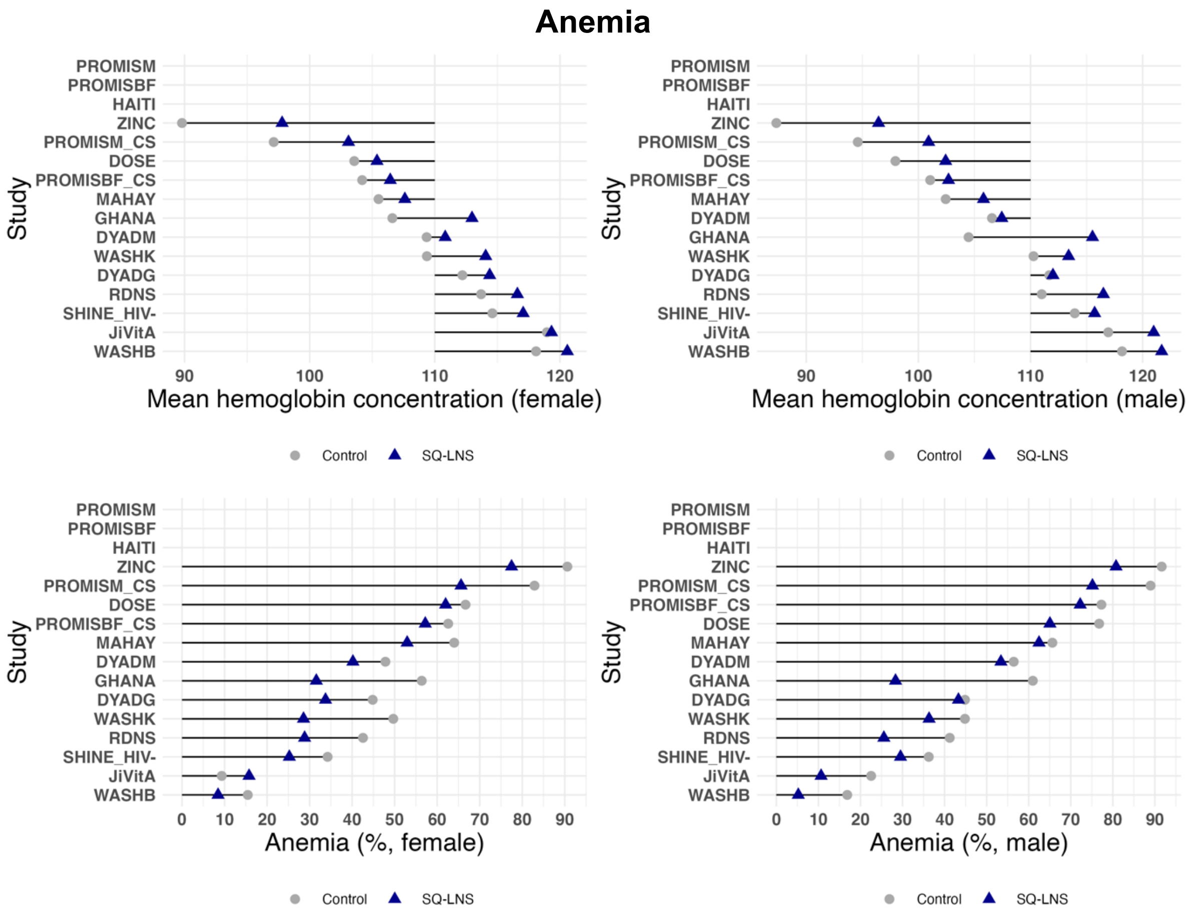

**Figure S34. Pooled Relative Index of Inequality and their 95% confidence intervals across studies for stunting, wasting, severe stunting and anemia at endline between intervention arms.** A. Female. B. Male. Pooled estimates were estimated using random-effect meta-analysis.

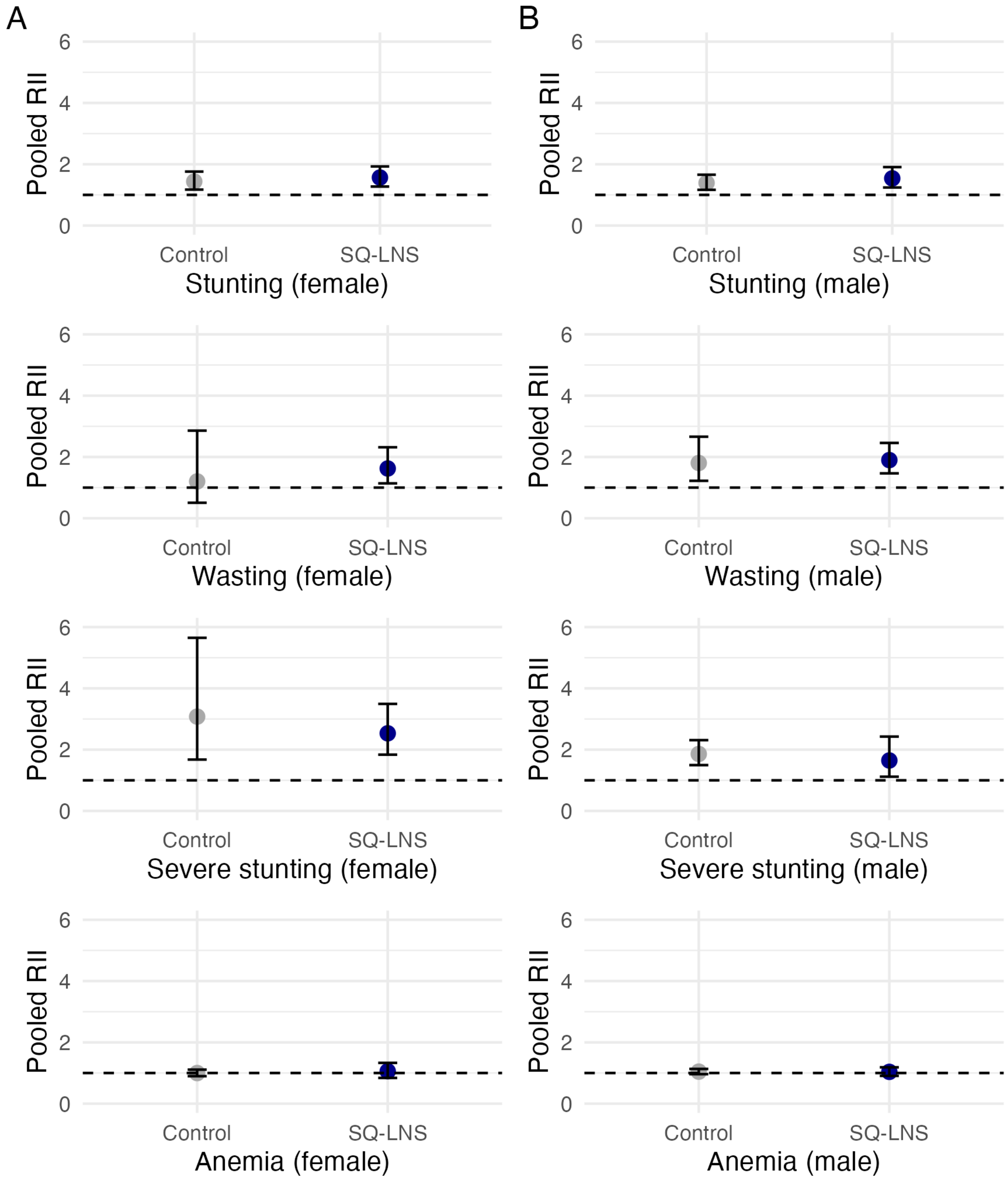

**Figure S35. Pooled Slope Index of Inequality and their 95% confidence intervals across studies for stunting, wasting, severe stunting and anemia at endline between intervention arms.** A. Female. B. Male. Pooled estimates were estimated using random-effect meta-analysis. The SII point estimates and their 95% confidence intervals were multiplied by 100 to express them as percentage points.

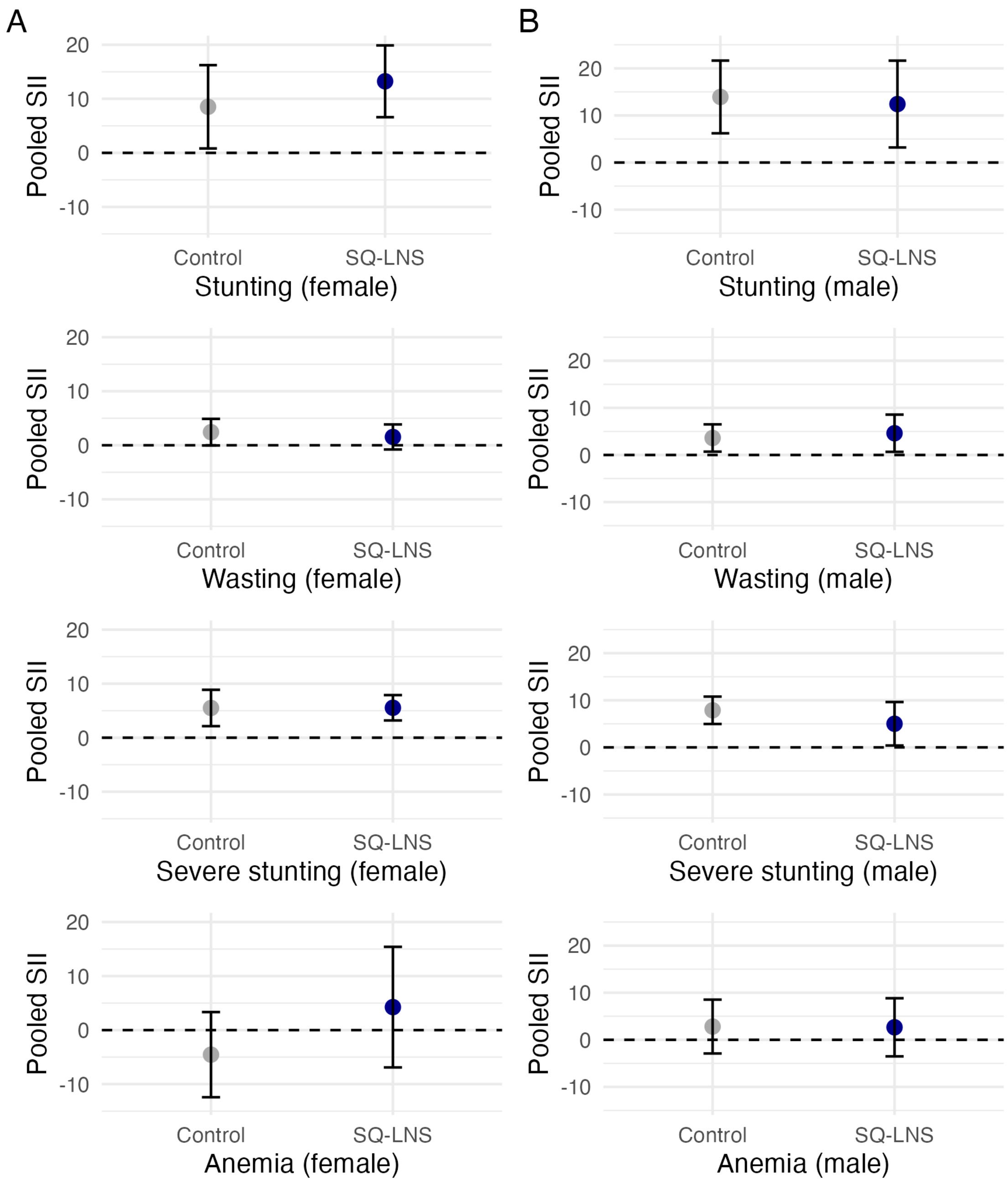

### Female

**Figure S36. Pooled female growth outcomes by intervention group (first row) and their absolute difference (second row). A.** Pooled length-for-age Z-score. **B.** Pooled weight-for-length Z-score. **C.** Pooled probability of stunting. **D.** Pooled probability of wasting. **E.** Probability of severe stunting. We pooled the child growth outcomes for each intervention arm using pointwise random-effect meta-analysis with Resticted Maximum Likelihood (REML). Intervention effects conditional on IWI were estimated by subtracting the spline fits for the intervention group from the control group within each study. These effects were then pooled using pointwise random-effects meta-analysis with REML. For the linear regression models (dashed lines), intervention effects were estimated by pooling the coefficients for the interaction term between IWI and intervention through random-effects meta-analysis and getting the difference between intervention groups. The pooled-p-for-interaction were from the random-effect meta-analysis of the pooled coefficients of interaction and standard errors across studies. Intervention included maternal supplementation. Shaded areas represent the 95% confidence intervals. F: Female.

#### Growth outcomes

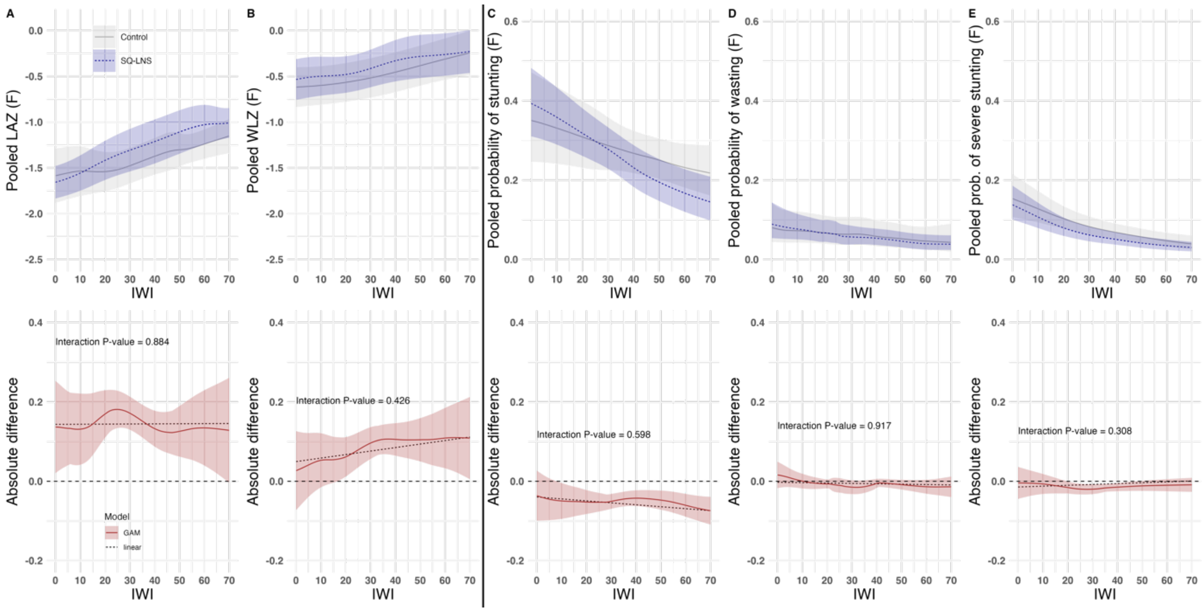

**Figure S37. Pooled female developmental outcomes by intervention group (first row) and their absolute difference (second row). A.** Pooled language score. **B.** Pooled gross motor score. **C.** Pooled fine motor score. **D.** Pooled executive function score. **E.** Pooled socioemotional score. We pooled the child development outcomes for each intervention arm using pointwise random-effect meta-analysis with Resticted Maximum Likelihood (REML). Intervention effects conditional on IWI were estimated by subtracting the spline fits for the intervention group from the control group within each study. These effects were then pooled using pointwise random-effects meta-analysis with REML. For the linear regression models (dashed lines), intervention effects were estimated by pooling the coefficients for the interaction term between IWI and intervention through random-effects meta-analysis and getting the difference between intervention groups. The pooled-p-for-interaction were from the random-effect meta-analysis of the pooled coefficients of interaction and standard errors across studies. Intervention included maternal supplementation. Shaded areas represent the 95% confidence intervals. Development scores are internal to each study distribution of the study-specific indicator. F: Female.

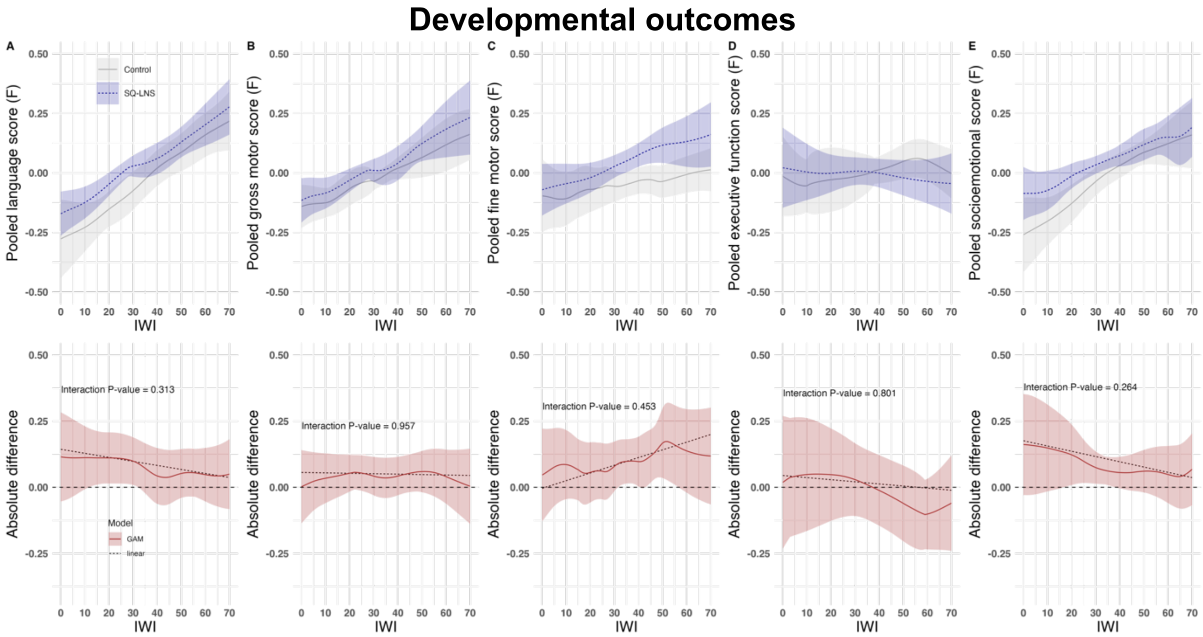

**Figure S38. Pooled female anemia outcomes by intervention group (first row) and their absolute difference (second row). A.** Pooled hemoglobin concentration. **B.** Pooled probability of anemia. We pooled the child hematological outcomes for each intervention arm using pointwise random-effect meta-analysis with Resticted Maximum Likelihood (REML). Intervention effects conditional on IWI were estimated by subtracting the spline fits for the intervention group from the control group within each study. These effects were then pooled using pointwise random-effects meta-analysis with REML. For the linear regression models (dashed lines), intervention effects were estimated by pooling the coefficients for the interaction term between IWI and intervention through random-effects meta-analysis and getting the difference between intervention groups. The pooled-p-for-interaction were from the random-effect meta-analysis of the pooled coefficients of interaction and standard errors across studies. Intervention included maternal supplementation. Shaded areas represent the 95% confidence intervals. F: Female.

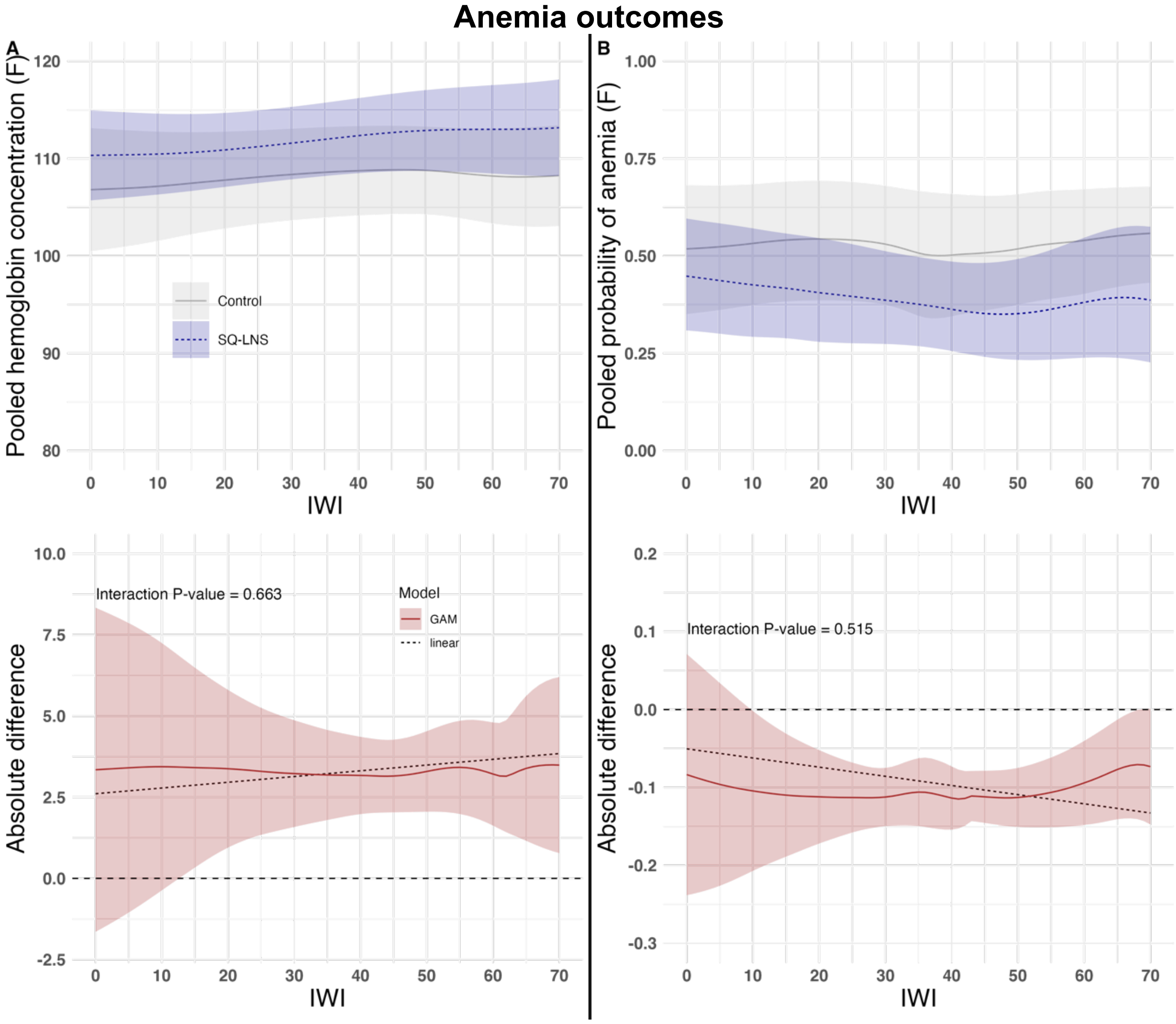

### Male

**Figure S39. Pooled male growth outcomes by intervention group (first row) and their absolute difference (second row). A.** Pooled length-for-age Z-score. **B.** Pooled weight-for-length Z-score. **C.** Pooled probability of stunting. **D.** Pooled probability of wasting. **E.** Probability of severe stunting. We pooled the child growth outcomes for each intervention arm using pointwise random-effect meta-analysis with Resticted Maximum Likelihood (REML). Intervention effects conditional on IWI were estimated by subtracting the spline fits for the intervention group from the control group within each study. These effects were then pooled using pointwise random-effects meta-analysis with REML. For the linear regression models (dashed lines), intervention effects were estimated by pooling the coefficients for the interaction term between IWI and intervention through random-effects meta-analysis and getting the difference between intervention groups. The pooled-p-for-interaction were from the random-effect meta-analysis of the pooled coefficients of interaction and standard errors across studies. Intervention included maternal supplementation. Shaded areas represent the 95% confidence intervals. M: Male.

#### Growth outcomes

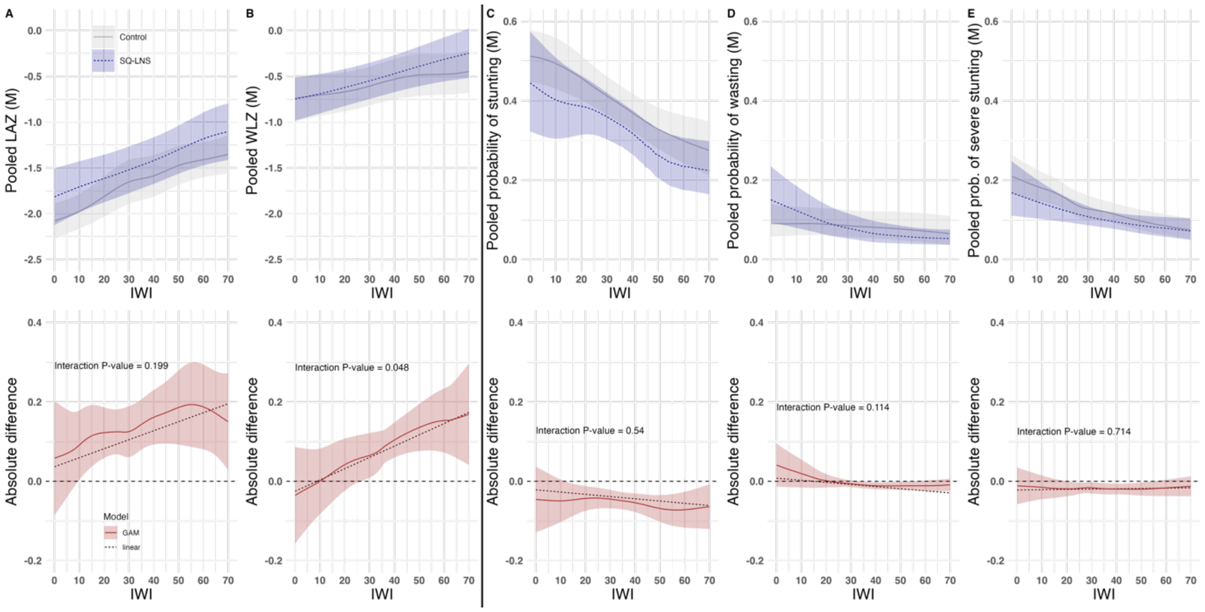

**Figure S40. Pooled male developmental outcomes by intervention group (first row) and their absolute difference (second row). A.** Pooled language score. **B.** Pooled gross motor score. **C.** Pooled fine motor score. **D.** Pooled executive function score. **E.** Pooled socioemotional score. We pooled the child development outcomes for each intervention arm using pointwise random-effect meta-analysis with Resticted Maximum Likelihood (REML). Intervention effects conditional on IWI were estimated by subtracting the spline fits for the intervention group from the control group within each study. These effects were then pooled using pointwise random-effects meta-analysis with REML. For the linear regression models (dashed lines), intervention effects were estimated by pooling the coefficients for the interaction term between IWI and intervention through random-effects meta-analysis and getting the difference between intervention groups. The pooled-p-for-interaction were from the random-effect meta-analysis of the pooled coefficients of interaction and standard errors across studies. Intervention included maternal supplementation. Shaded areas represent the 95% confidence intervals. Development scores are internal to each study distribution of the study-specific indicator. M: Male.

#### Developmental outcomes

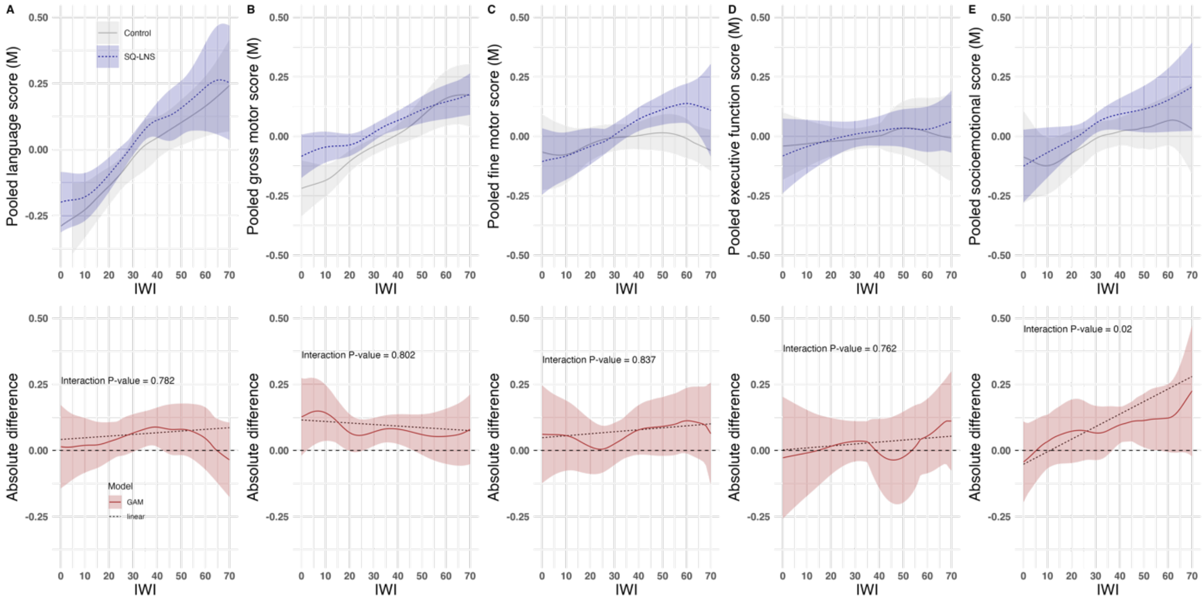

**Figure S41. Pooled male anemia outcomes by intervention group (first row) and their absolute difference (second row). A.** Pooled hemoglobin concentration. **B.** Pooled probability of anemia. We pooled the child hematological outcomes for each intervention arm using pointwise random-effect meta-analysis with Resticted Maximum Likelihood (REML). Intervention effects conditional on IWI were estimated by subtracting the spline fits for the intervention group from the control group within each study. These effects were then pooled using pointwise random-effects meta-analysis with REML. For the linear regression models (dashed lines), intervention effects were estimated by pooling the coefficients for the interaction term between IWI and intervention through random-effects meta-analysis and getting the difference between intervention groups. The pooled-p-for-interaction were from the random-effect meta-analysis of the pooled coefficients of interaction and standard errors across studies. Intervention included maternal supplementation. Shaded areas represent the 95% confidence intervals. M: Male.

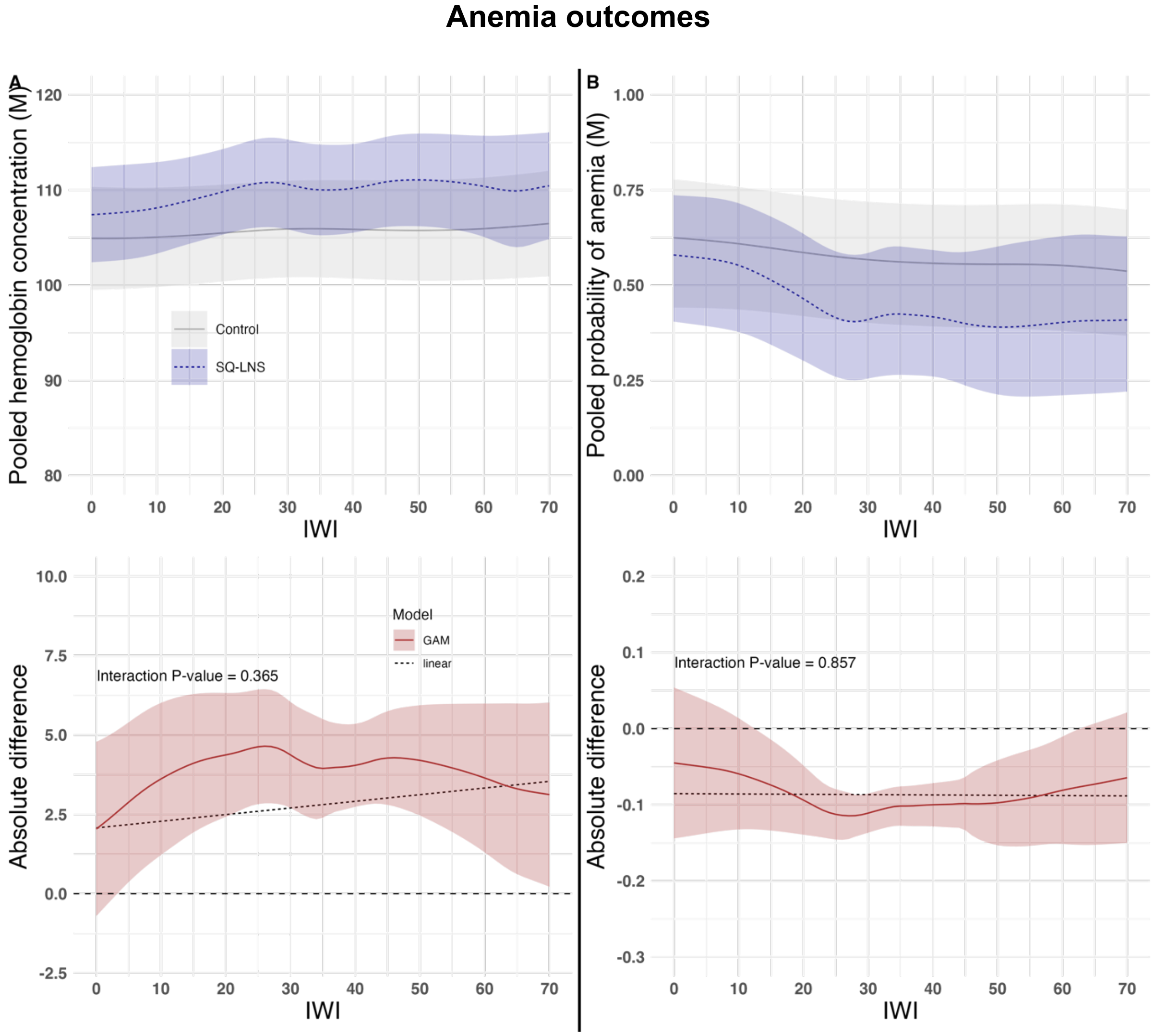

#### Text S6. Maternal education as socioeconomic indicator.

We observed consistent results when using IWI except for the outcome hemoglobin concentration where we found a significant interaction (maternal education x intervention) p-value using a one-stage meta-analysis (**Table S5**). However, results were consistent when using a two-stage meta-analysis for each factor level.

**Table S5. Using maternal education as effect modifier of intervention on child growth, development and anemia outcomes.**

| Child outcome | Global Wald-type F test p-value for interaction (based on one-stage analysis) | Pooled coefficient for interaction and 95% CI (Intermediate×SQ-LNS)* | Pooled coefficient for interaction and 95% CI (Secondary/higher education×SQ-LNS)* |
|---|---|---|---|
| **Growth** | | | |
| **LAZ** | 0.5332 | -0.015 (-0.080; 0.049) | -0.059 (-0.250; 0.132) |
| **WLZ** | 0.4017 | -0.025 (-0.083; 0.032) | 0.036 (-0.078; 0.150) |
| **Stunting** | 0.2047 | -0.012 (-0.041; 0.017) | 0.018 (-0.140; 0.176) |
| **Wasting** | 0.1938 | 0.008 (-0.004; 0.020) | 0.006 (-0.010; 0.023) |
| **Severe stunting** | 0.7051 | 0.012 (-0.014; 0.037) | 0.020 (-0.001; 0.041) |
| **Development** | | | |
| **Language score** | 0.1047 | -0.031 (-0.121; 0.059) | 0.155 (-0.336; 0.647) |
| **Gross motor score** | 0.3488 | -0.057 (-0.129; 0.016) | -0.041 (-0.157; 0.074) |
| **Fine motor score** | 0.258 | -0.160 (-0.286; -0.035) | -0.088 (-0.285; 0.109) |
| **Executive function score** | 0.9344 | -0.024 (-0.151; 0.102) | -0.155 (-0.372; 0.061) |
| **Socioemotional score** | 0.3727 | -0.010 (-0.140; 0.120) | 0.262 (-0.336; 0.860) |
| **Anemia** | | | |
| **Hemoglobin concentration** | p-value < 0.0001 | 1.082 (-0.911; 3.075) | 0.308 (-2.708; 3.323) |
| **Anemia** | 0.1188 | -0.029 (-0.098; 0.041) | -0.004 (-0.090; 0.083) |

*Pooled the coefficients across studies using two-stage random-effect meta-analysis.
LAZ: Length-for-age z-score; WLZ: Weight-for-length z-score; CI: Confidence interval, SQ-LNS: Small quantity lipid-based nutrient supplement. Intermediate education: Complete primary and incomplete secondary, Secondary/higher: Secondary and higher education.

#### Text S7. Adherence level by International Wealth Index.

We estimated adherence levels for a subset of trials with readily available individual-level adherence data across IWI categories (0–20, 21–40, 41–60, 61–80, and 81–100). These trials included iLiNS-DOSE, iLiNS-DYAD-G, iLiNS-DYAD-M, iLiNS-ZINC, RDNS, WASH-B, WASH-K, PROMIS-BF and PROMIS-M. Adherence measures varied across studies. In the iLiNS trials, adherence was defined as the percentage of days per week that caregivers reported the child consumed SQ-LNS. In RDNS, it was based on the number of supplement packets or tablets consumed in the past week. For WASH-B, adherence was calculated as the percentage of expected SQ-LNS consumed, while in WASH-K, it was defined as the number of sachets consumed in the past week divided by 14. For the PROMIS studies, they were defined as the percentage of monthly LNS doses received.

**Figure S42. Adherence levels in the study arm that received small quantity lipid-based nutrient supplements by International Wealth Index (IWI) across sub-studies with available data.** Adherence was measured differently across studies. **A.** ILiNS studies: Adherence reflects the percentage of days per week that caregivers reported the child consumed SQ-LNS. **B.** RDNS, WASH Benefits Bangladesh, WASH Benefits Kenya, PROMIS Burkina Faso and PROMIS Malawi: Adherence definitions varied by study.

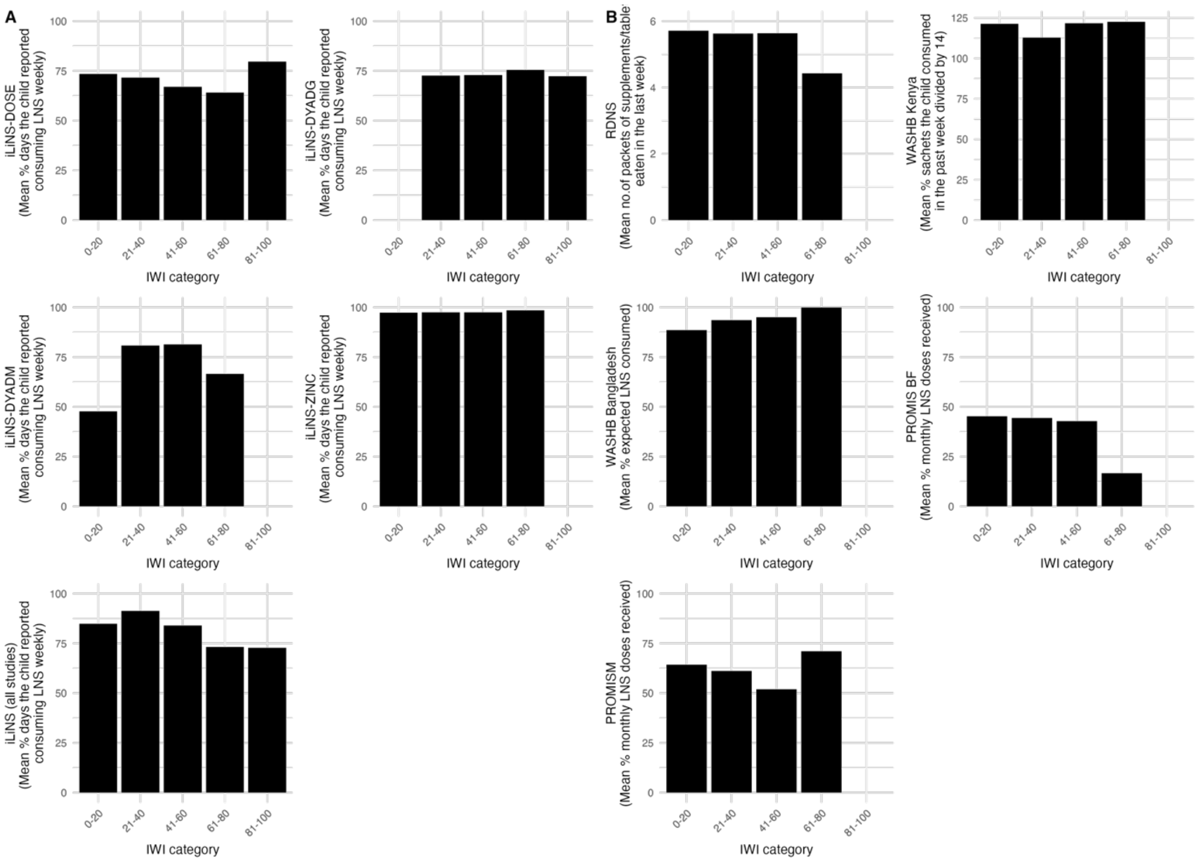


For PROMIS-BF and PROMIS-M, there were only one and two observations, respectively, for the mean adherence in the IWI 61–80 category, making them unrepresentative. Additionally, the PROMIS studies were more programmatic in nature — implemented through the health system — which typically resulted in lower adherence.
